# County Prevalence and Control of High Blood Pressure from Health Kiosks, 2017-2024

**DOI:** 10.1101/2025.08.13.25333546

**Authors:** Thomas W. Hsiao, Lauren Fede, Cameron Gocke, Lance A. Waller, Mohammed K. Ali, Jithin Sam Varghese

**Affiliations:** Department of Biostatistics and Bioinformatics, Rollins School of Public Health, Emory University, Atlanta, Georgia, USA; Pursuant Health LLC, Atlanta, Georgia, USA; Emory Global Diabetes Research Center of Woodruff Health Sciences Center and Emory University, Atlanta, Georgia, USA; Hubert Department of Global Health, Rollins School of Public Health, Emory University, Atlanta, Georgia, USA; Department of Family and Preventive Medicine, School of Medicine, Emory University, Atlanta, Georgia, USA

**Keywords:** Care continuum, surveillance, hypertension

## Abstract

**Background:** As the leading modifiable risk factor for death in the United States, hypertension requires timely and locally detailed surveillance. Current estimates of the high blood pressure (BP) care continuum are lagged national averages from sample surveys with low participation and unknown subnational variation. This study explores the use of self-service health kiosks in retail stores as an alternative source for subnational estimates of prevalence, awareness, and control.

**Methods:** We analyzed data from adult kiosk users (n = 1,270,485) across 1,892 counties in 49 States (except Massachusetts) and District of Columbia in a serial cross-sectional analysis from November 2017 to September 2024. High BP was defined as self-reported diagnosed or elevated BP (systolic ≥140 mmHg or diastolic ≥90 mmHg). Among those diagnosed, control was defined as BP <140/90 mmHg. Small area estimates for counties were calculated using multilevel regression and poststratification based on individual and areal socio-demographic covariates. We compared the prevalence of diagnosed hypertension with the Behavioral Risk Factor Surveillance System 2021.

**Results:** The analytic sample had a mean age of 42.0 years (SD=15.6). Prevalence of high BP was 51.9% in 2017-2018 and 50.4% in 2023-2024. In 2023-2024, awareness and control were 73.7% and 61.8% and county-level prevalence ranged from 39.5% to 63.1%. Similar hotspots in the Southeast were identified in both kiosk and BRFSS (Spearman’s ρ = 0.52).

**Conclusions:** Health kiosk data reveal substantial spatial and socio-demographic variation in high BP. Near real-time sub-national surveillance of health kiosk users can provide insights to guide interventions and track progress.

**NOVELTY AND RELEVANCE:** *What is New?:*

- Self-service health kiosk data complement national surveys by offering direct BP measurements with wider geographic coverage and higher representation of vulnerable populations.
- This study provides recent state- and county- level estimates of prevalence, awareness, and control of high BP for 1.3 million kiosk users.

*What is Relevant?:*

- In 2023-2024, prevalence of high BP among kiosk users exceeded 50.0% with county-level variation ranging from 39.5% in the Mountain West to 63.1% in the Southeast.
- National awareness and control were suboptimal at 73.7% and 61.8%.
- These results highlight persistent gaps in high BP detection and management.

*Clinical/Pathophysiological Implications?:*

- Near real-time kiosk-based BP surveillance can help identify high-risk populations and administer timely interventions to reduce cardiovascular risk.

## Introduction

Hypertension is the primary modifiable risk factor for stroke and cardiovascular disease and affects an estimated 120 million adults in the United States.^1^ Although awareness of high blood pressure (BP) has increased over the last two decades, nearly 11 million adults, i.e., 15% of those with hypertension are unaware of their high BP and only 43% among those taking antihypertensive medication have their BP under control.^1–4^ Recent analyses show both a national decline in control and persistent disparities by race & ethnicity, age, and income.^5–8^ Current estimates and trend data are all derived from annual or biennial sample surveys that lack temporal or spatial granularity as estimates are lagged by 3-4 years and have inadequate statistical power to detect appreciable differences at the local county level. As such, state and county health departments do not have disaggregated data for local level planning, implementation, and monitoring of blood pressure screening and management initiatives.

To address this gap, the Centers for Disease Control & Prevention (CDC) releases small areal estimates through data initiatives such as PLACES: Local Data for Better Health and the Selected Metropolitan/Micropolitan Area Risk Trends (or SMART).^9,10^ These initiatives rely on self-reported estimates of diagnosed and treated high BP from state and metropolitan area level surveys conducted by the National Center for Health Statistics and the Behavioral Risk Factor Surveillance System (BRFSS).^11^ BRFSS does not track prevalence of undiagnosed high BP and BP control among those with hypertension.^3^

There is a critical need for alternative strategies to monitor county-level population health, especially among vulnerable populations that tend to be non-responders in national surveys.^12,13^ Furthermore, participation in surveys is low and declining. Response rates for telephone-based BRFSS surveys were 47% in 2004 and 45% in 2023 [range for states: 22-63%]).^14,15^ Bias from non-response depends on whether participation was random, and previous studies have shown higher response rates among older, white participants and lower response rates among low-income, younger, and racial & ethnic minority participants.^12,16,17^

Efforts are already underway for leveraging administrative data sources such as electronic health records (EHR) and insurance claims for surveillance of hypertension and other non-communicable diseases.^18–20^ While such data sources are an important alternative to national surveys for local estimation of awareness and control of high BP, these data sources also cannot capture undiagnosed individuals, and tend to underrepresent marginalized populations who cannot or do not access health care.^21,22^ Effective national and sub-national surveillance is needed, especially among marginalized communities.^23,24^

In previous work, we presented national-level summaries of high BP derived from Pursuant Health self-service health kiosks located in Walmart and select CVS stores from November 2017 to September 2024.^25^ Compared to National Health and Nutrition Examination Survey (NHANES) and BRFSS, the analytic sample had higher representation of younger individuals, racial ethnic minorities, and rural populations who are traditionally underrepresented in national surveys.^25^ Here, we extend the analysis to estimate prevalence, awareness and control of high BP by state, county, and socio-demographic group in the United States. We also assessed whether aggregated kiosk-based estimates correlate with national survey data. Findings from this study can aid population health monitoring of high-risk groups with the goal of administering timely interventions to reduce cardiovascular risk.

## Methods

### Data Source

Pursuant Health provides free health screenings in Walmart and select CVS stores through voluntary self-service kiosks. Kiosks are classified by the Food and Drug Administration (FDA) as Class II Medical Devices.^25^ Session data was provided under a data use agreement (DUA) approved by both parties by authors (LF and CG) who were not involved in data analysis and interpretation. The DUA outlines the provision of a Limited Data Set as described under the HIPAA Privacy Rule.

### Study Population and Data collection

The study population and analytic sample selection have been described in detail in a prior publication.^25^ We summarize the selection here. Of all kiosk sessions from November 2017 to September 2024, 91.9 million recorded BP. Self-reported age, gender, and race & ethnicity were available for each respondent, along with the geographic location and time of kiosk use.

Location was classified as urban or rural based on the kiosk county’s 2023 Rural-Urban Commuting Area Code. We only included sessions of users who self-reported their hypertension status and further excluded sessions of users reporting age outside the range of 18 to 99 years, missing gender information, recording unreasonable blood pressure readings (either SBP outside of 60 to 300 mmHg or DBP outside of 30 to 300 mmHg), or having duplicate sessions within a two-year interval.

We defined high blood pressure as either 1) diagnosed high BP based on self-report (responding “yes” to “Have you ever been diagnosed with high blood pressure by a health provider?”) or 2) undiagnosed high BP, defined as no self-reported history of hypertension diagnosis but a recorded elevated SBP (≥ 140 mmHg) or DBP (≥ 90 mmHg) from the kiosk. Among those with diagnosed plus undiagnosed high BP, we defined awareness as the proportion with diagnosed high BP. Among those who were aware, we defined BP control as the proportion with SBP < 140 mmHg and DBP < 90 mmHg.

### Statistical Analysis

We generated county, state, and national level estimates of the high BP care continuum (prevalence, awareness, control) and diagnosed high BP by age group (18-19, 20-44, 45-64, 65 and older), gender (men, women), race and ethnicity (Hispanic, Non-Hispanic (NH) White, NH Black, NH Asian, NH Other), and urbanicity using multilevel regression and poststratification (MRP).^26,27^ MRP has been widely used in various small area estimation applications with nonrepresentative data, including estimating voter turnout among subgroups and modeling population health outcomes in United States counties.^28–30^

We followed a two-stage modeling approach. First, we fit a mixed effects logistic regression model on the sample data, including nested random intercepts over county and state, and fixed effects for individual-level demographic poststratification variables (age group, racial ethnic group, and gender), county-level covariates from the 5-year American Community Survey 2018-2022 (ACS), and a state-level indicator for geographic region (Northeast, South, Midwest, West).^31^ Estimates were generated separately for 2-year reporting periods (2017-2018, 2019- 2020, 2021-2022, 2023-2024), corresponding to NHANES data collection cycles. Second, we computed aggregated estimates using poststratification weighting. Prevalence, awareness, and control were calculated as a weighted average of stratified estimates for subgroups defined by county, age category, race & ethnicity, and gender. Detailed discussion of statistical methods and computation of the care continuum can be found in the **Supplementary Methods Note** in **Supplementary File 1**. Estimates and standard errors for prevalence, awareness, and control of high BP based on the 2017 AHA/ACC cutoffs (130/80 mmHg) are also available as supplementary material (**Supplementary File 2, Supplementary File 3**).

Our choice of which auxiliary county-level covariates to extract from the ACS was based on a previous study of factors contributing to county-level variation in cardiovascular disease,^32^ including median household income (in current United States dollars), the percentage of residents who are unemployed, the percentage of residents who lack a high school education, and the percentage of residents without health insurance.

We assessed the external validity of our estimates by benchmarking the county estimates of diagnosed high BP in the total population among the analytic sample in 2021-2022 with the PLACES 2023 data based on BRFSS 2021.^9,33,34^ A summary of demographic characteristics and comparison of the analytic sample to BRFSS survey populations are available in a prior publication.^25^ We calculated Local Moran’s I to measure spatial cluster similarity between PLACES 2023 and the post stratified estimates from the analytic sample.

## Results

The analytic sample of 1,270,485 individuals covered 49 states (excluding Massachusetts) and Washington DC and included 1,892 counties, with most of the absent counties located in sparsely populated areas within the Great Plains and Mountain states. We observed a decline in participants from 528,247 in 2019 to just 92,366 participants in 2020, coinciding with the first year of the COVID-19 pandemic in the U.S.

A comparison of estimates for the national continuum of care for high BP for NHANES (2017-March 2020, 2021-2023), BRFSS (2019, 2023), and the analytic sample are presented in the **Table**. Compared to NHANES, the prevalence of high BP from kiosk data was higher, while awareness and control were lower. Model-based estimates of national high BP prevalence from the analytic sample were 51.9% (95% CI: 51.0-52.7) in 2017-2018 and 50.4% (95% CI: 49.6-51.1) in 2023-2024.

Awareness increased from 68.7% (95% CI: 67.9-69.5) in 2017-2018 to 73.7% (95% CI: 73.0-74.4) in 2023-2024. Proportions achieving control among those aware increased from 53.0% (95% CI: 52.1-53.9) to 61.8% (95% CI: 61.1-62.7) between 2017-2018 and 2023-2024.

Prevalence estimates showed variability between states (Range: 45.5-56.1%, IQR: 3.34 in 2023-2024). The highest prevalence estimates were concentrated in southern and midwestern states (**Figure 1A**): the largest cluster was identified in a spatially contiguous area encompassing Alabama, Mississippi, Louisiana, Arkansas, and Missouri. States in the western and northeastern United States exhibited relatively lower estimates.

**Figure 1.**
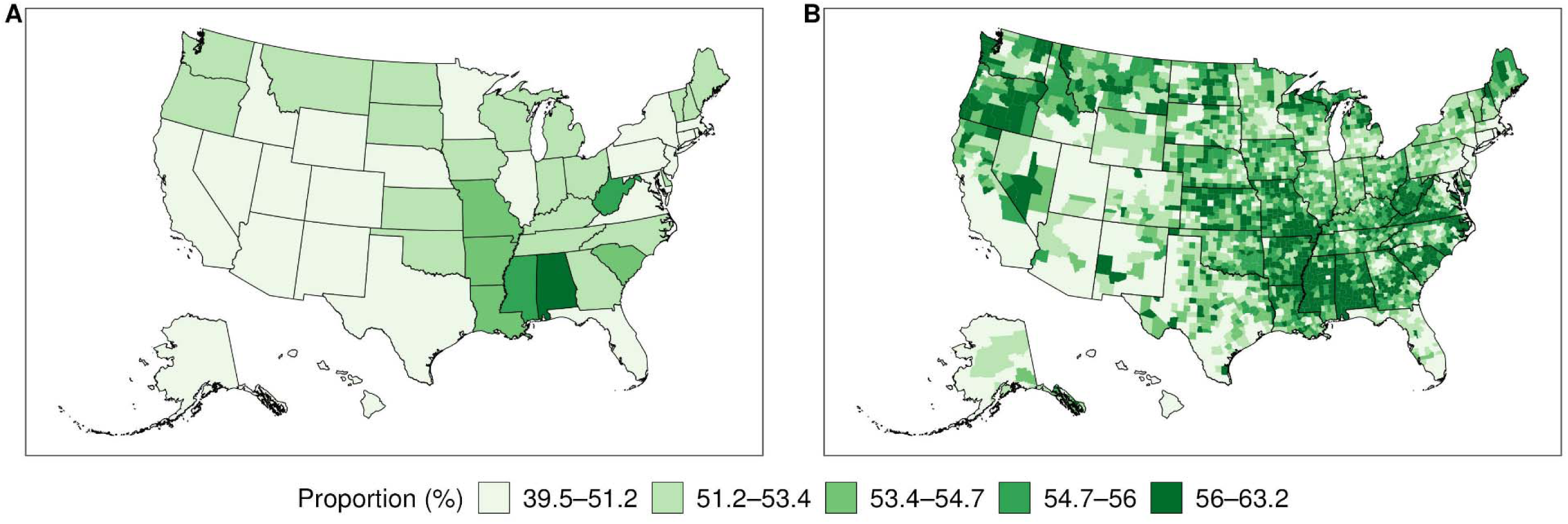
Estimates of high blood pressure prevalence, 2023-2024. Panel A and Panel B are estimates of prevalence of high BP (self-reported diagnosed high BP or undiagnosed high BP with elevated SBP ≥ 140 mmHg or DBP ≥ 90 mmHg) at state and county level. Visualizations for all years are presented in **Supplementary File 2**. Estimates of uncertainty are presented in **Supplementary File 3**.

County-level estimates showed even greater heterogeneity (Range: 39.5-63.1%, IQR: 3.87 in 2023-2024), with the lowest prevalence counties located in the West and Northeast and highest prevalence counties in the Southeast. Additional high-burden clusters emerged even within states located in regions with low prevalence. For example, while overall prevalence was lower in the western states, county-level estimates revealed clusters of high prevalence in Idaho, Oregon, and Washington (**Figure 1B, Figure S1**).

County-level awareness (Range: 67.4-83.0%, IQR: 2.56) mirrored prevalence patterns, with the highest awareness levels observed in the Southeast though these areas tended to have lower control rates (**Figure 2B**). In contrast, the highest levels of county-level control (Range: 55.0-70.7%, IQR: 2.53) were in Arizona, New Mexico, Texas, Florida, Virginia, and the Northeast (**Figure 3B**).

**Figure 2.**
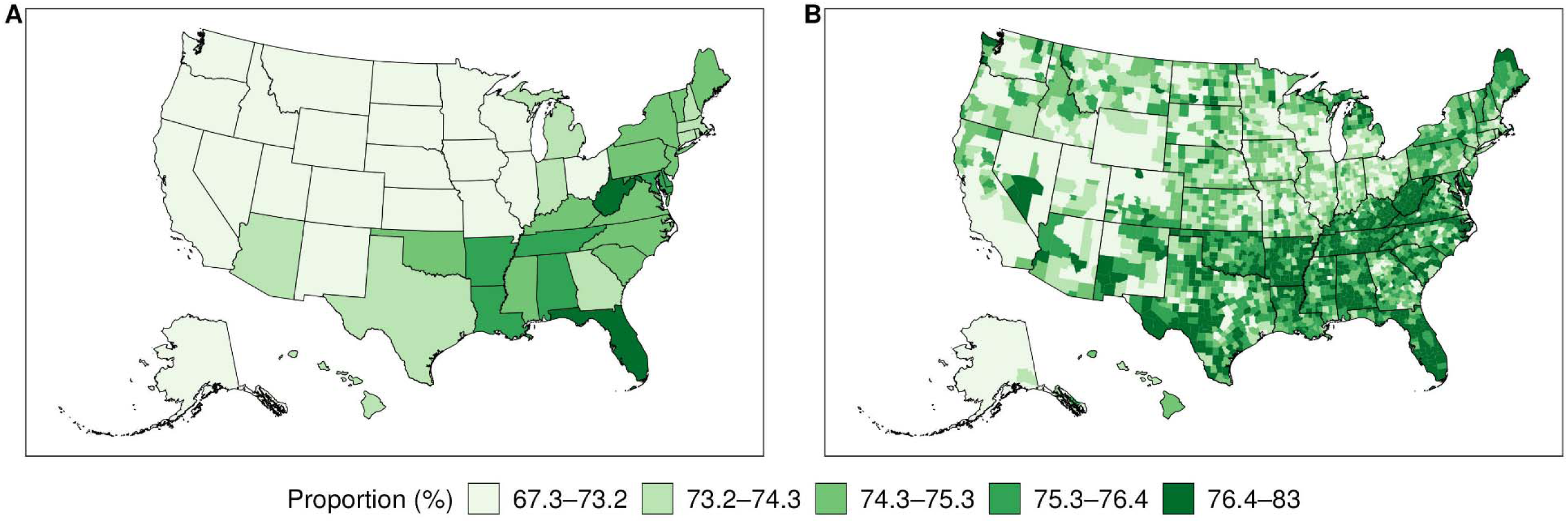
Estimates of awareness, 2023-2024. Panel A and Panel B are estimates of proportion aware among those with high BP at state and county level. Visualizations for all years are presented in **Supplementary File 2**. Estimates of uncertainty are presented in **Supplementary File 3**.

**Figure 3.**
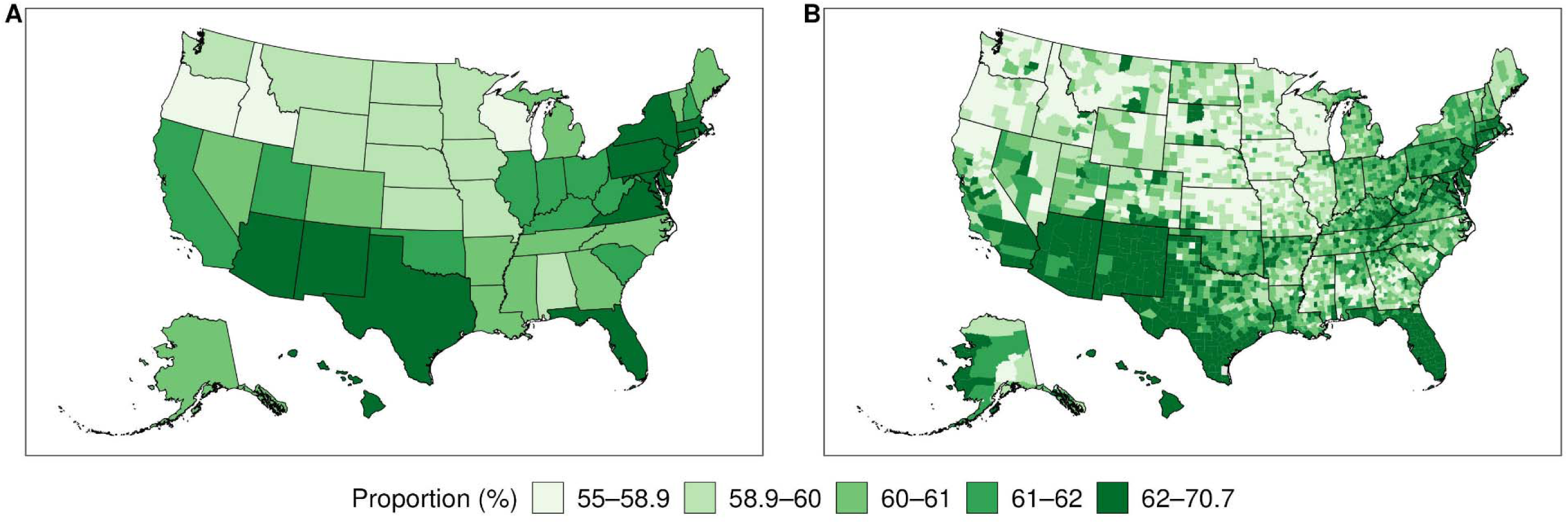
Estimates of control, 2023-2024. Panel A and Panel B are estimates of proportion controlled among those aware of their hypertension at state and county level. Visualizations for all years are presented in Supplementary File 2. Estimates of uncertainty are presented in **Supplementary File 3**.

**Figure 4.**
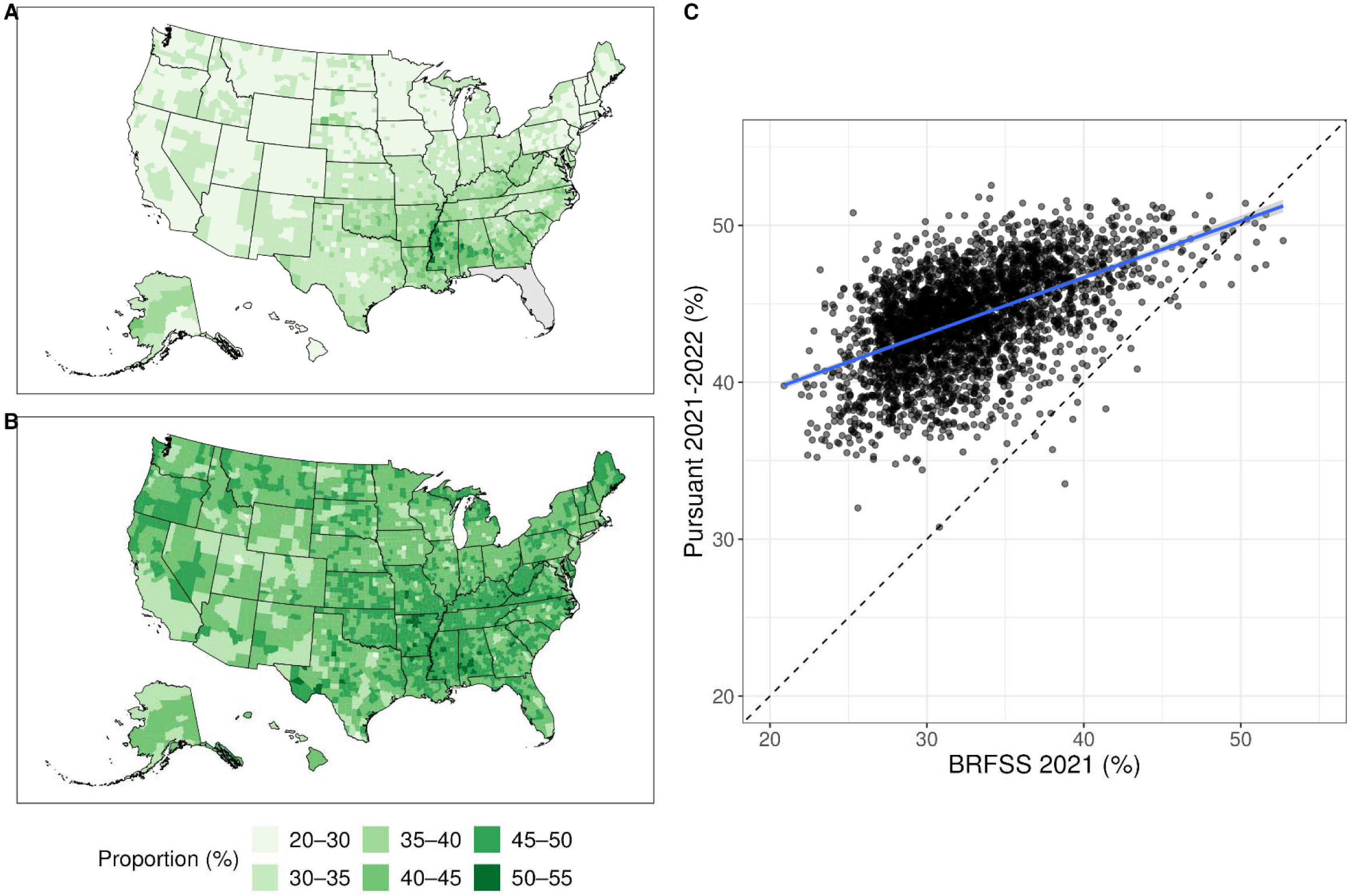
Prevalence of diagnosed high BP for PLACES and the analytic sample in United States counties. Panel A and Panel B are modeled estimates of diagnosed high BP in the population from Behavioral Risk Factor Surveillance System 2021 (PLACES 2023) and Pursuant 2021-2022. Panel C shows the correlation between modeled estimates of diagnosed high BP (ρ = 0.52) between both sources.

Precision in county-level estimates varied, reflecting differences in data availability and population sizes (**Supplementary File 3**). Further breakdowns of prevalence, awareness, control and diagnosed high BP by age, gender, and race & ethnicity can be found in **Supplementary File 4**. Black populations had the highest prevalence of high BP among all racial ethnic groups and high levels of awareness across time periods but the lowest levels of controlled hypertension by a substantial amount. We also found that post-stratification generally resulted in higher estimates compared to unadjusted state-level rates (**Supplementary File 5**).

### Comparison to BRFSS

Local estimates of prevalence of diagnosed high BP in United States counties were correlated (ρ = 0.52) between the analytic sample and PLACES 2023 (**Figure 3A-B**). County-level estimates of diagnosed high BP were consistently higher in the kiosk analytic sample than in PLACES 2023 (**Figure 3C**). At the same time, Local Moran’s I identified similar hypertension hotspots in the Southeastern U.S. in both the analytic sample and PLACES (**Supplementary File 1 - Figure S1)**. The kiosk estimates revealed more outlier counties compared to PLACES, particularly low outliers in metropolitan areas and high outliers in the Midwest and Pacific Northwest.

## Discussion

We developed local and recent county-level estimates of the high BP care continuum from local BP kiosks. National prevalence of high BP was high while awareness and control were suboptimal. The modeled estimates of high BP prevalence, awareness, control, and diagnosed high BP displayed substantial geographic variability. Estimates of diagnosed high BP among kiosk users were systematically higher across all counties compared to PLACES 2023 but were correlated and spatially similar. Our results illustrate how careful analysis of voluntary and non-representative data sources can complement existing efforts to monitor health, especially in socially vulnerable communities that are traditionally underrepresented in research and survey studies.

Findings from this study build on research using EHR and survey data wherein care continua using EHRs report higher estimates of diagnosis than national surveys, although observed geographic and sociodemographic trends remain similar.^18,19^ However, studies using EHR and surveys do not provide county-level estimates for all counties and did not include individuals with undiagnosed high BP.^35^ This limits their ability to comprehensively estimate the care continuum and generate valid subnational comparisons to surveys like NHANES.^25^

Our study has three key strengths relative to traditional surveys and methods. First, the analytic sample has greater access to historically vulnerable populations and difficult to survey groups compared to traditional national surveys. Over two-thirds of Americans shop at Walmart stores, with rural and minority shoppers being overrepresented among shoppers.^36,37^ On the other hand, responders to national surveys like BRFSS are predominantly older and white, and the most recent NHANES wave no longer samples sufficient numbers of underrepresented minorities for reliable estimates. Second, the kiosks permit rapid survey data collection and near real-time monitoring of local policy effects. In contrast, BRFSS and NHANES require substantial time for preparation, administration, and post processing before release. Kiosk data are automatically recorded and are not dependent on costly in-person visits. Lastly, the availability of both self-reported survey questions and blood pressure readings from the kiosk combined with statistical methodology like the MRP generate rapid estimates and uncertainties for the hypertension care continuum at the county level, making kiosks a potentially valuable tool for future public health responses during crises.

Our study is not without limitations. Prevalence of high BP in the analytic sample may not indicate prevalence of hypertension since clinical diagnosis of hypertension requires elevated BP (≥130/80 mmHg) from two measurements on at least two separate days. As a tool for estimating high BP prevalence at the population level, the self-service kiosk data is also subject to selection bias. The individuals in the analytic sample were self-selected according to an unknown mechanism, which can affect the generalizability of our estimates to the population.

However, our model-based estimates statistically adjust for county, age, gender, race and ethnicity, under an assumption of non-differential selection by these socio-demographic covariates, following an approach similar to national surveys. However, we found that upon adjustment for covariates and poststratification, kiosk participants had higher prevalence of diagnosed high BP, especially among those under 45 years of age, across all counties (**Supplementary File 4**). One possible explanation is that the kiosk users, who were disproportionately racial & ethnic minorities and rural adults, are more likely to experience hypertension at younger ages and/or are underrepresented in BRFSS.^3^ Another explanation could be that those with more severe and uncontrolled disease are more likely to check their BP at kiosks, either due to symptom onset or anxiety about control.

We highlight three opportunities provided by non-traditional data sources beyond hypertension monitoring. First, platforms such as self-service kiosks can assess self-reported health through questionnaires. Second, by having users log into an individualized account tracking their measurements, there are opportunities for routine, longitudinal monitoring of blood pressure for individuals who may lack regular access to a health provider or a home BP monitoring device.^38^ Longitudinal hypertension care continua can provide even more nuanced data on maintenance of control and which patients to target to close persistent care gaps.^39^ Third, by combining these data with those of other administrative and survey data sources using novel data fusion strategies, it may be possible to derive more precise and accurate indicators of hypertension care continua.^23^ Additionally, the performance of the MRP model can be improved by introducing dependence between the levels of the care continuum and flexible nonlinearities between the spatial predictors and response^40^.

### Perspectives

In the United States, response rates are declining for nationally representative surveys.^15,41,42^ Previous reports suggest one in two adults have hypertension, 15% of adults are undiagnosed and over 77% have uncontrolled hypertension. However, these estimates mask substantial geographic and socio-demographic differences in the care continuum that are not routinely captured using modeled estimates from self-reported data on diagnosed hypertension. Leveraging near real-time data from voluntary blood pressure measurements at self-service kiosks, we provide an alternative description of the United States’ hypertension care continuum nationally, at the state and county levels, and by socio-demographic group, and find that prevalence is high, and awareness and control remain at suboptimal levels. Non-representative data sources can augment current surveillance efforts for monitoring policy initiatives aimed at mitigating the hypertension burden.

## Ethics approval and consent to participate

We were exempt from ethical approval by the Institutional Review Board of Emory University for this secondary analysis of the proprietary Limited Data Set.

## Consent for publication

Not applicable

## Data availability statement

The proprietary data used in this manuscript is available upon signing a data use agreement and at the discretion of Pursuant Health LLC. The code and data used to generate figures and tables are available at https://github.com/jvargh7/spatial_kiosks.

## Author contributions

LF and CG extracted the data and were not involved in the analysis and interpretation of results. TH drafted the analytic plan with inputs from LF, JSV, MKA and LW. TH conducted all analysis and wrote the first draft with inputs from JSV, LW and MKA. All authors edited subsequent drafts and approved the final draft of the manuscript.

## Acknowledgements

We thank Mark Hutcheson (Emory Global Diabetes Research Center) and Rajsekhar Guddneppanavar (Emory Office of Technology Transfer) for their administrative support. We thank David Rosenblatt (Pursuant Health) and Leslie Gerdes-Sommers (Pursuant Health) for their inputs on the manuscript.

---

**Table.**
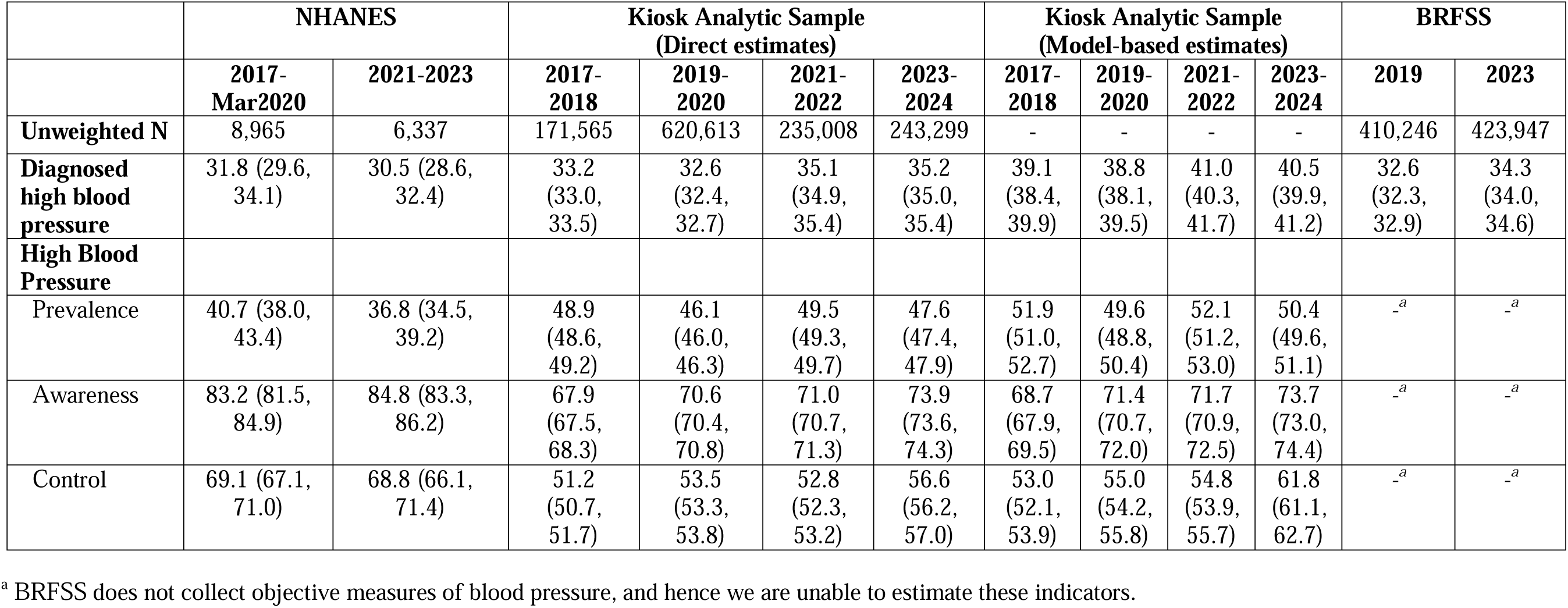
National estimates of hypertension prevalence, awareness and control.

|  | NHANES |  | Kiosk Analytic Sample<br>(Direct estimates) |  |  |  | Kiosk Analytic Sample<br>(Model-based estimates) |  |  |  | BRFSS |  |
| --- | --- | --- | --- | --- | --- | --- | --- | --- | --- | --- | --- | --- |
|  | 2017-Mar2020 | 2021-2023 | 2017-2018 | 2019-2020 | 2021-2022 | 2023-2024 | 2017-2018 | 2019-2020 | 2021-2022 | 2023-2024 | 2019 | 2023 |
| **Unweighted N** | 8,965 | 6,337 | 171,565 | 620,613 | 235,008 | 243,299 | - | - | - | - | 410,246 | 423,947 |
| **Diagnosed high blood pressure** | 31.8 (29.6, 34.1) | 30.5 (28.6, 32.4) | 33.2 (33.0, 33.5) | 32.6 (32.4, 32.7) | 35.1 (34.9, 35.4) | 35.2 (35.0, 35.4) | 39.1 (38.4, 39.9) | 38.8 (38.1, 39.5) | 41.0 (40.3, 41.7) | 40.5 (39.9, 41.2) | 32.6 (32.3, 32.9) | 34.3 (34.0, 34.6) |
| **High Blood Pressure** |  |  |  |  |  |  |  |  |  |  |  |  |
| Prevalence | 40.7 (38.0, 43.4) | 36.8 (34.5, 39.2) | 48.9 (48.6, 49.2) | 46.1 (46.0, 46.3) | 49.5 (49.3, 49.7) | 47.6 (47.4, 47.9) | 51.9 (51.0, 52.7) | 49.6 (48.8, 50.4) | 52.1 (51.2, 53.0) | 50.4 (49.6, 51.1) | -<sup>a</sup> | -<sup>a</sup> |
| Awareness | 83.2 (81.5, 84.9) | 84.8 (83.3, 86.2) | 67.9 (67.5, 68.3) | 70.6 (70.4, 70.8) | 71.0 (70.7, 71.3) | 73.9 (73.6, 74.3) | 68.7 (67.9, 69.5) | 71.4 (70.7, 72.0) | 71.7 (70.9, 72.5) | 73.7 (73.0, 74.4) | -<sup>a</sup> | -<sup>a</sup> |
| Control | 69.1 (67.1, 71.0) | 68.8 (66.1, 71.4) | 51.2 (50.7, 51.7) | 53.5 (53.3, 53.8) | 52.8 (52.3, 53.2) | 56.6 (56.2, 57.0) | 53.0 (52.1, 53.9) | 55.0 (54.2, 55.8) | 54.8 (53.9, 55.7) | 61.8 (61.1, 62.7) | -<sup>a</sup> | -<sup>a</sup> |

<sup>a</sup> BRFSS does not collect objective measures of blood pressure, and hence we are unable to estimate these indicators.

