## Supplementary File 1 for "County Prevalence and Control of High Blood Pressure from Health Kiosks, 2017-2024"

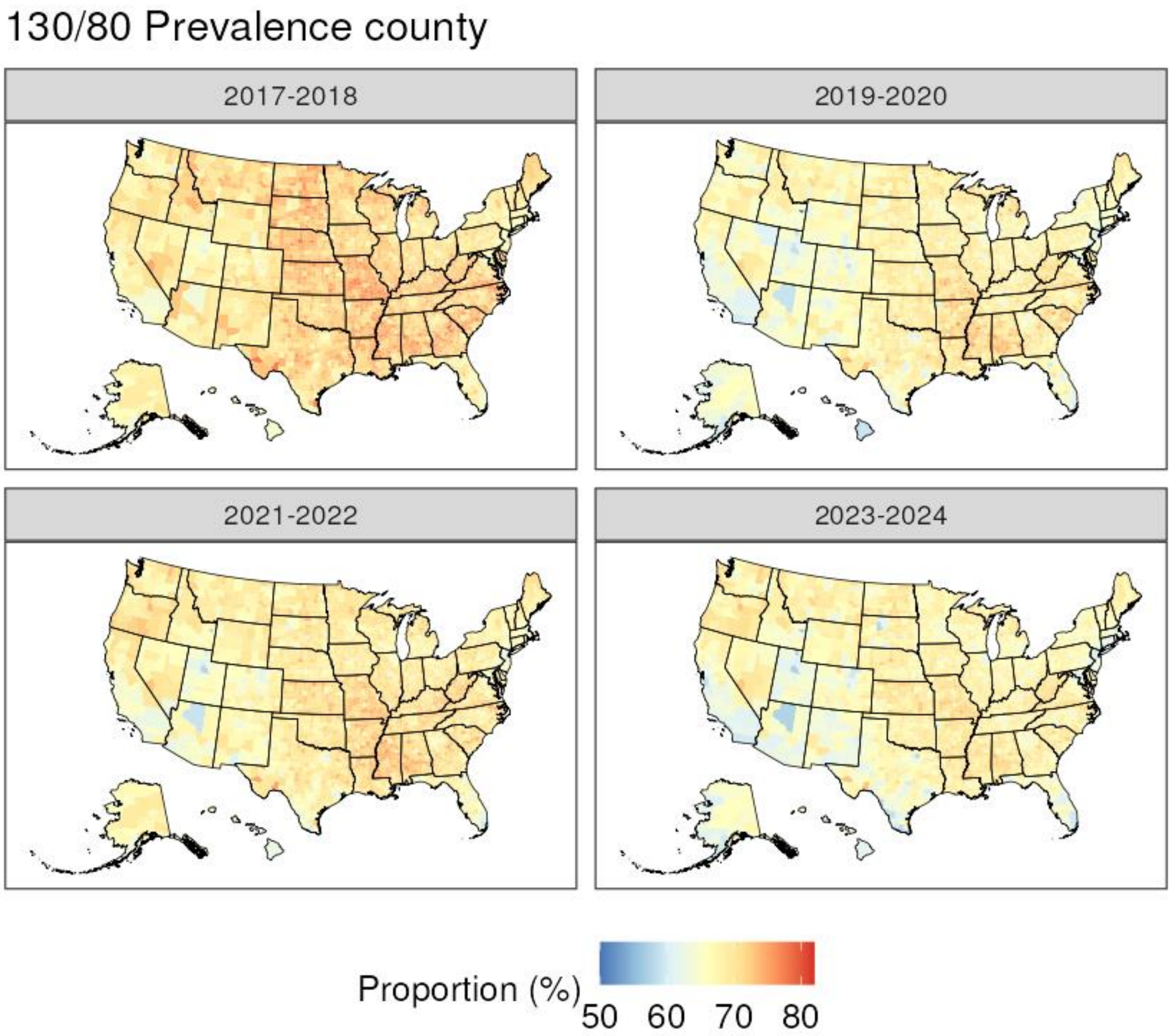
130/80 Prevalence county
2017-2018
2019-2020
2021-2022
2023-2024
Proportion (%)
50 60 70 80

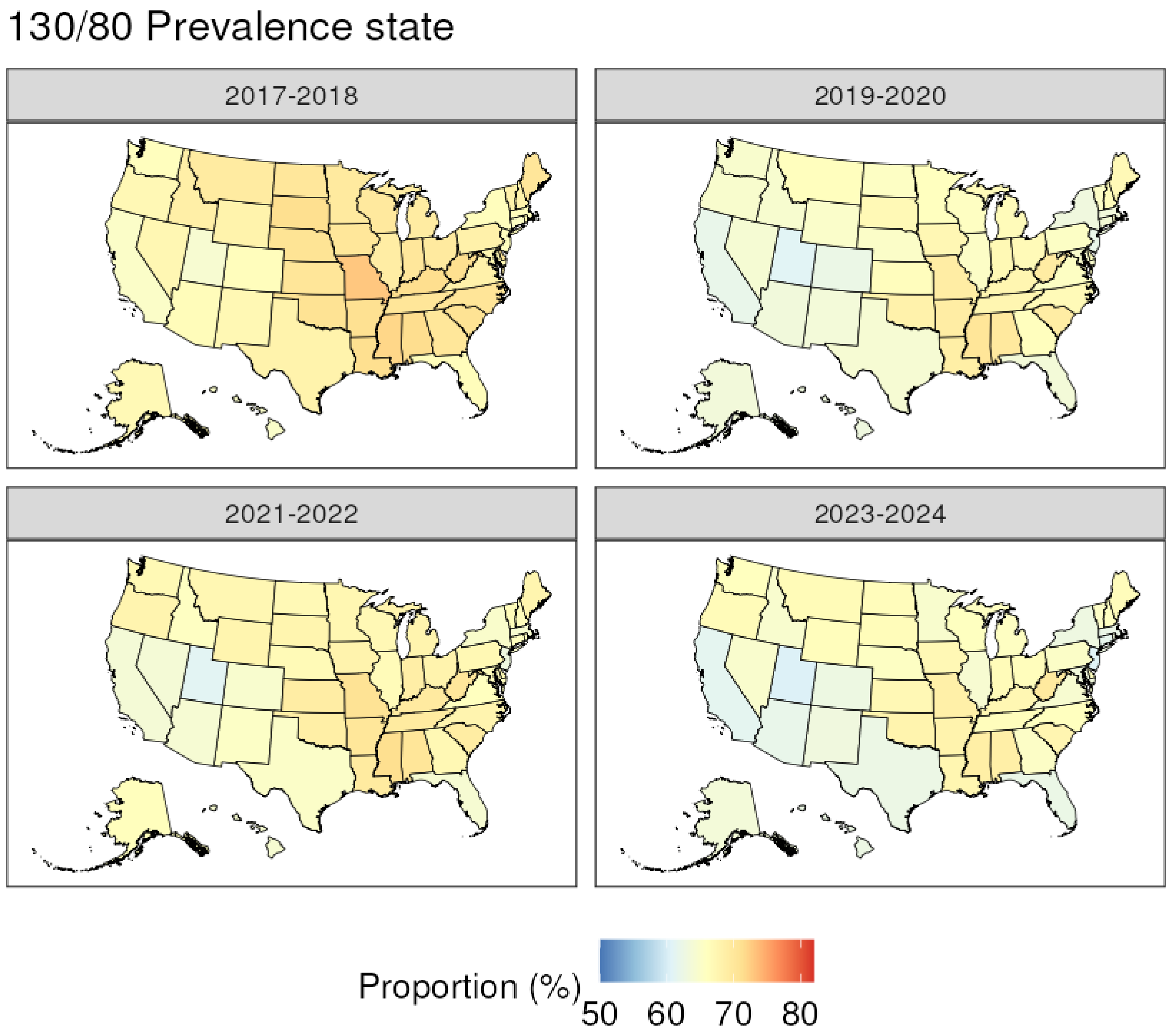
130/80 Prevalence state
2017-2018
2019-2020
2021-2022
2023-2024
Proportion (%)
50
60
70
80

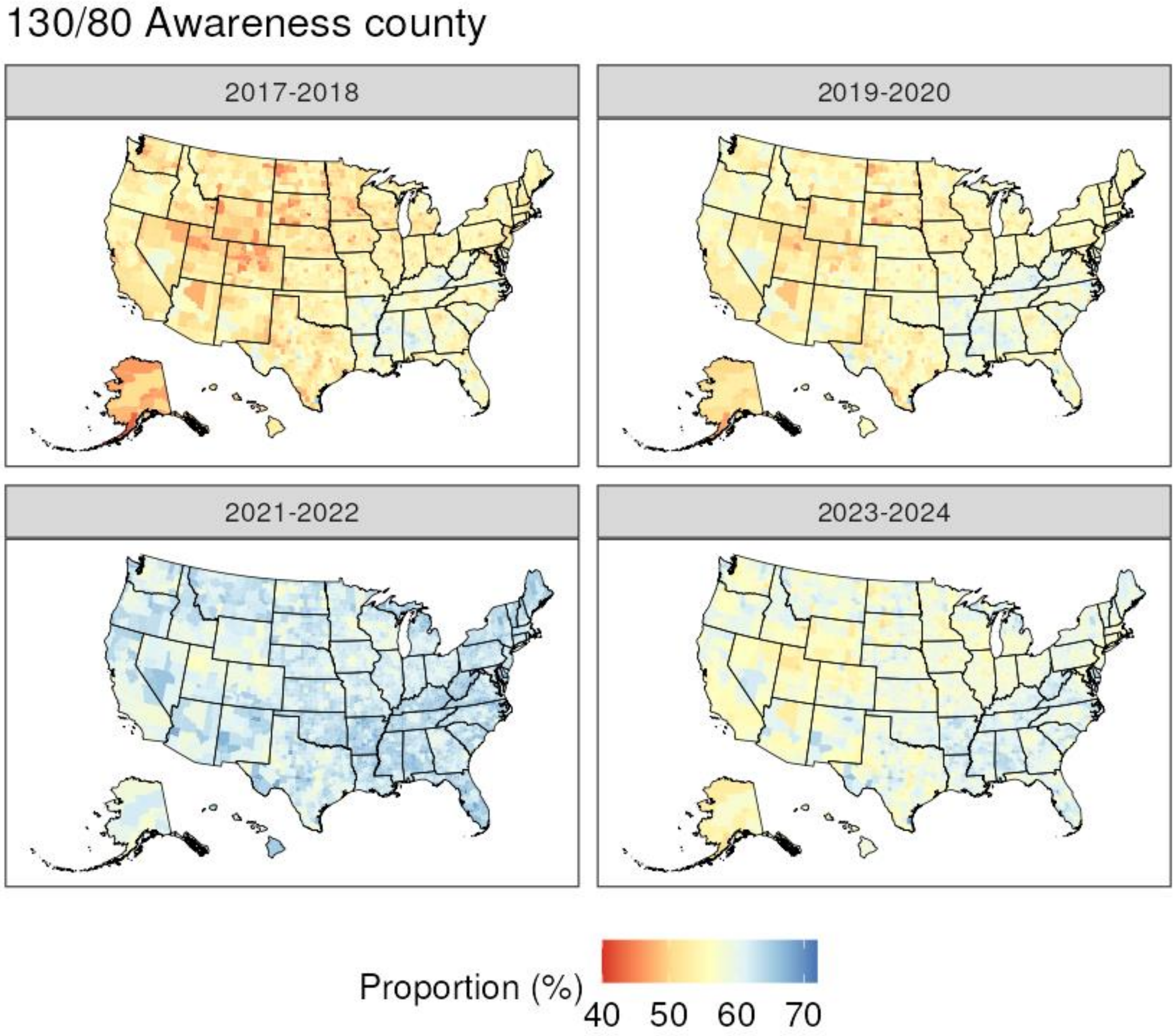

130/80 Awareness county
2017-2018
2019-2020
2021-2022
2023-2024
Proportion (%)
40 50 60 70

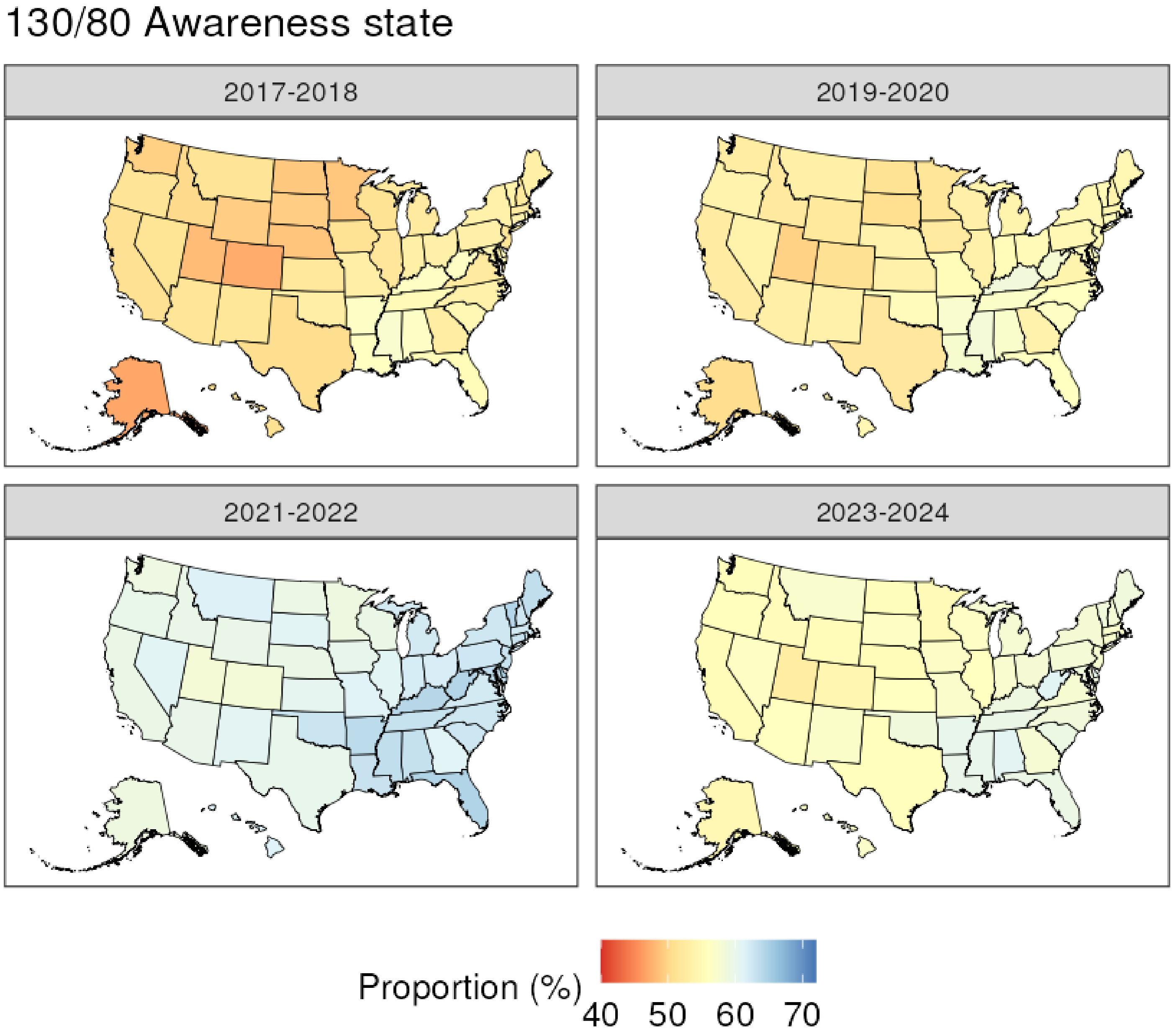
130/80 Awareness state
2017-2018
2019-2020
2021-2022
2023-2024
Proportion (%)
40
50
60
70

### 130/80 Controlled county

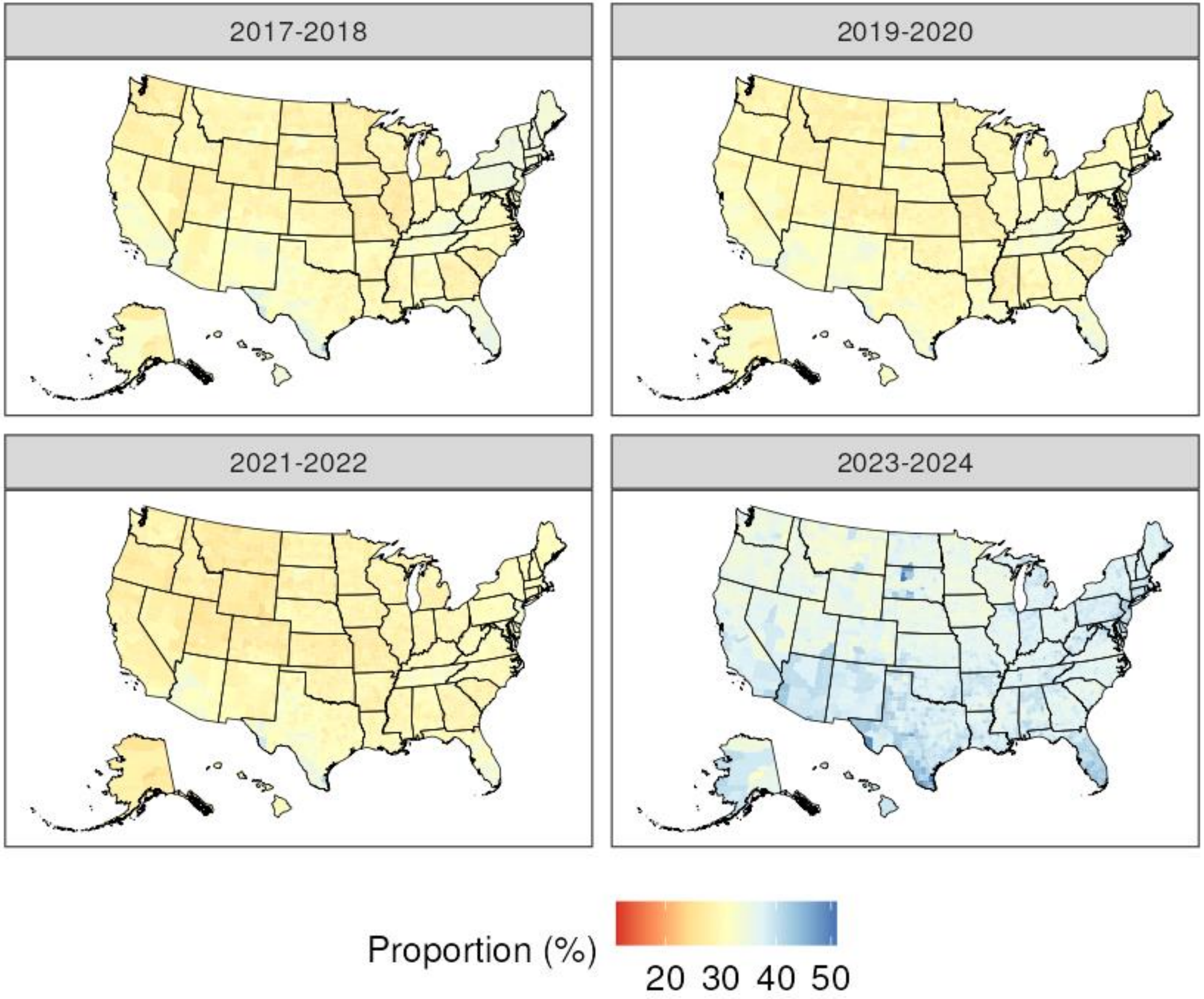

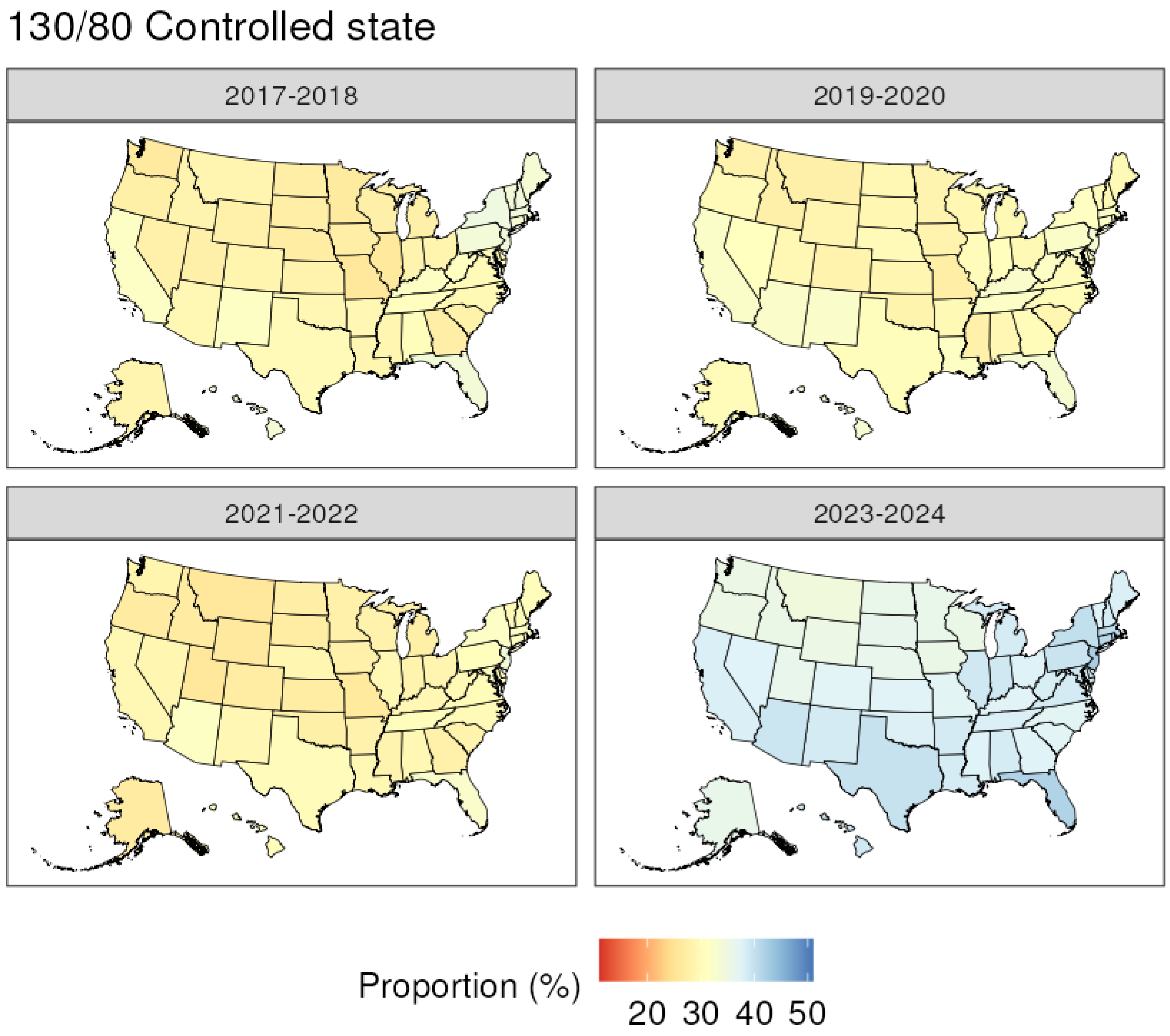

130/80 Controlled state
2017-2018
2019-2020
2021-2022
2023-2024
Proportion (%)
20 30 40 50

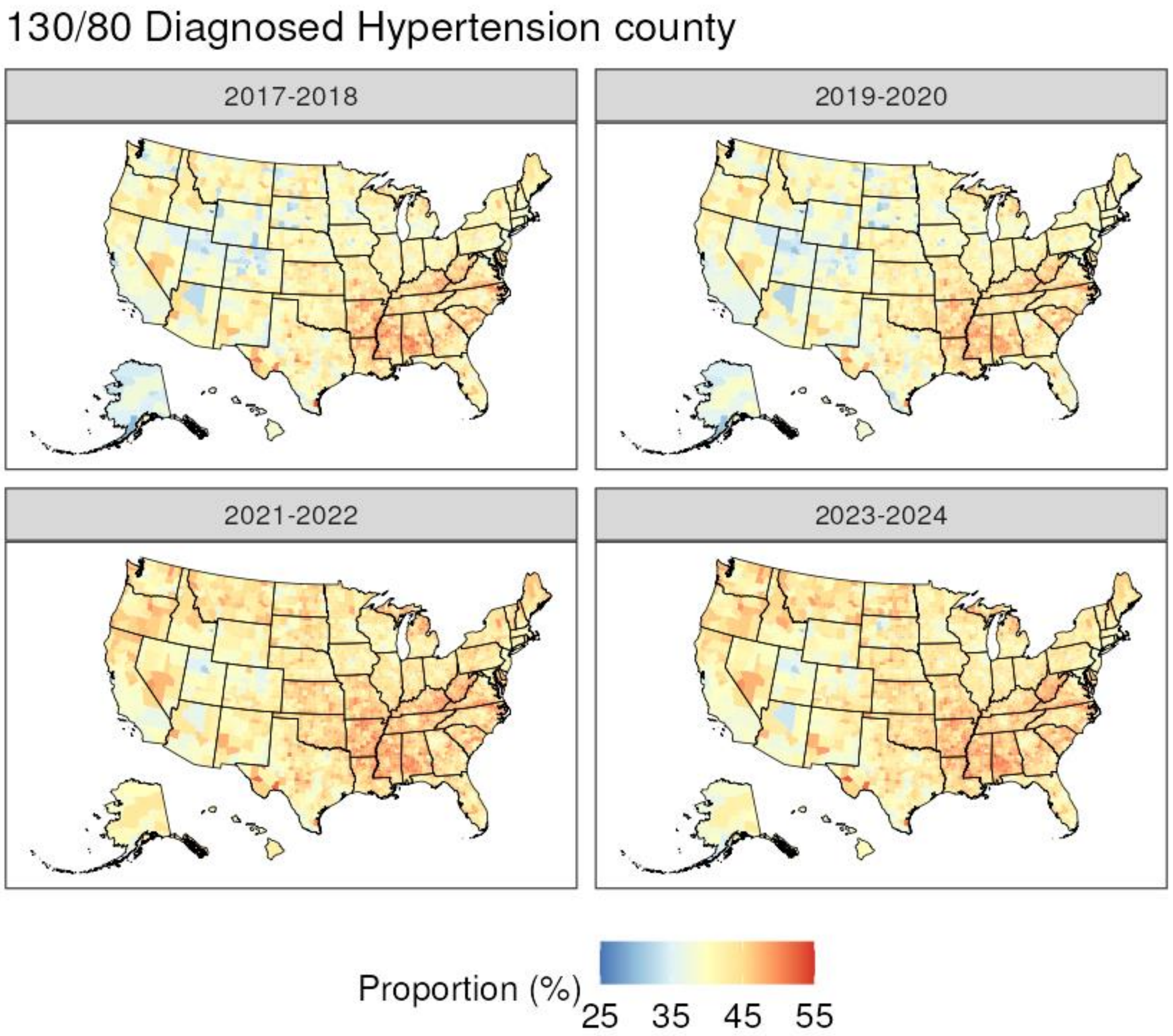
130/80 Diagnosed Hypertension county
2017-2018
2019-2020
2021-2022
2023-2024
Proportion (%)
25
35
45
55

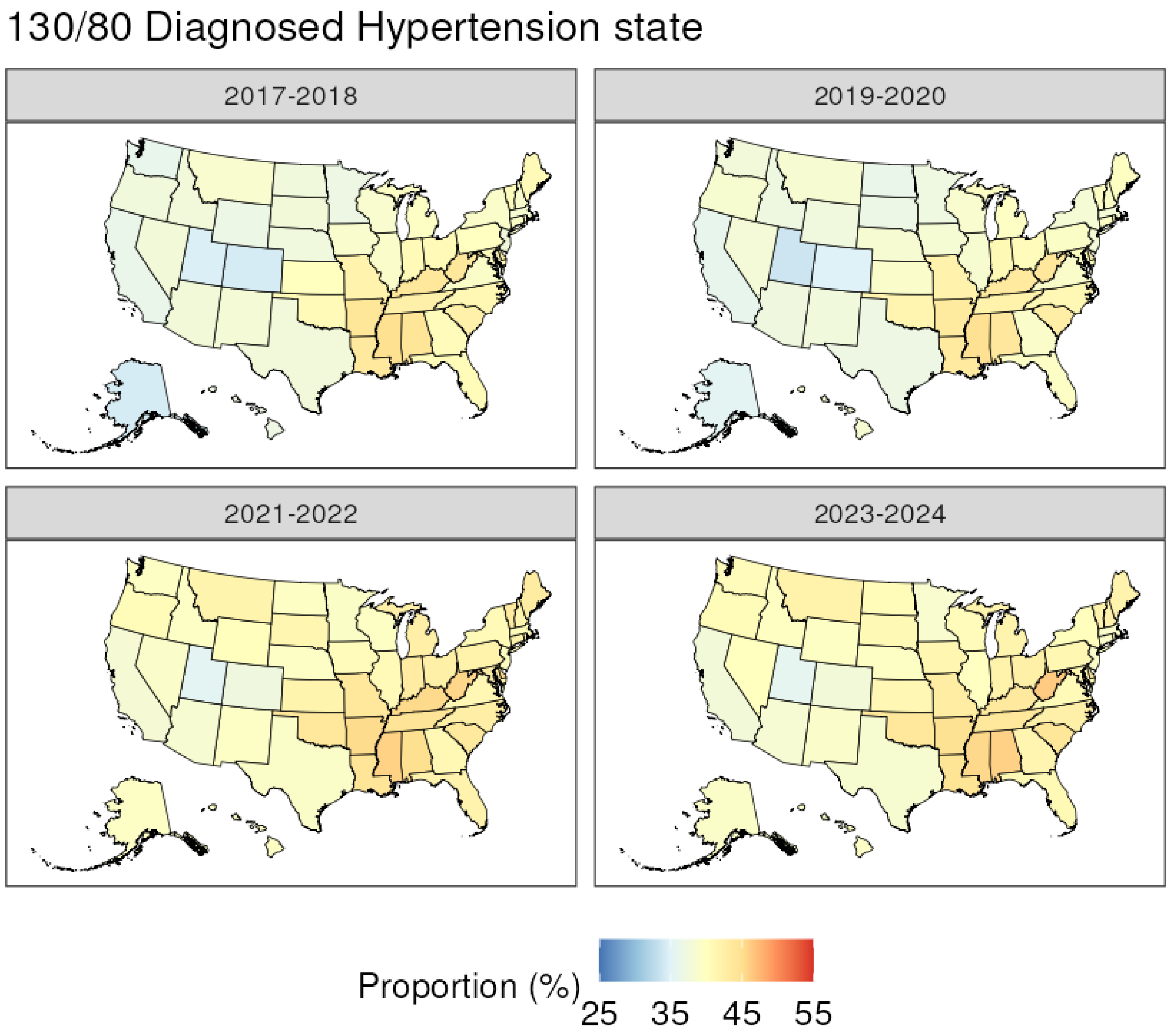
130/80 Diagnosed Hypertension state
2017-2018
2019-2020
2021-2022
2023-2024
Proportion (%)
25
35
45
55

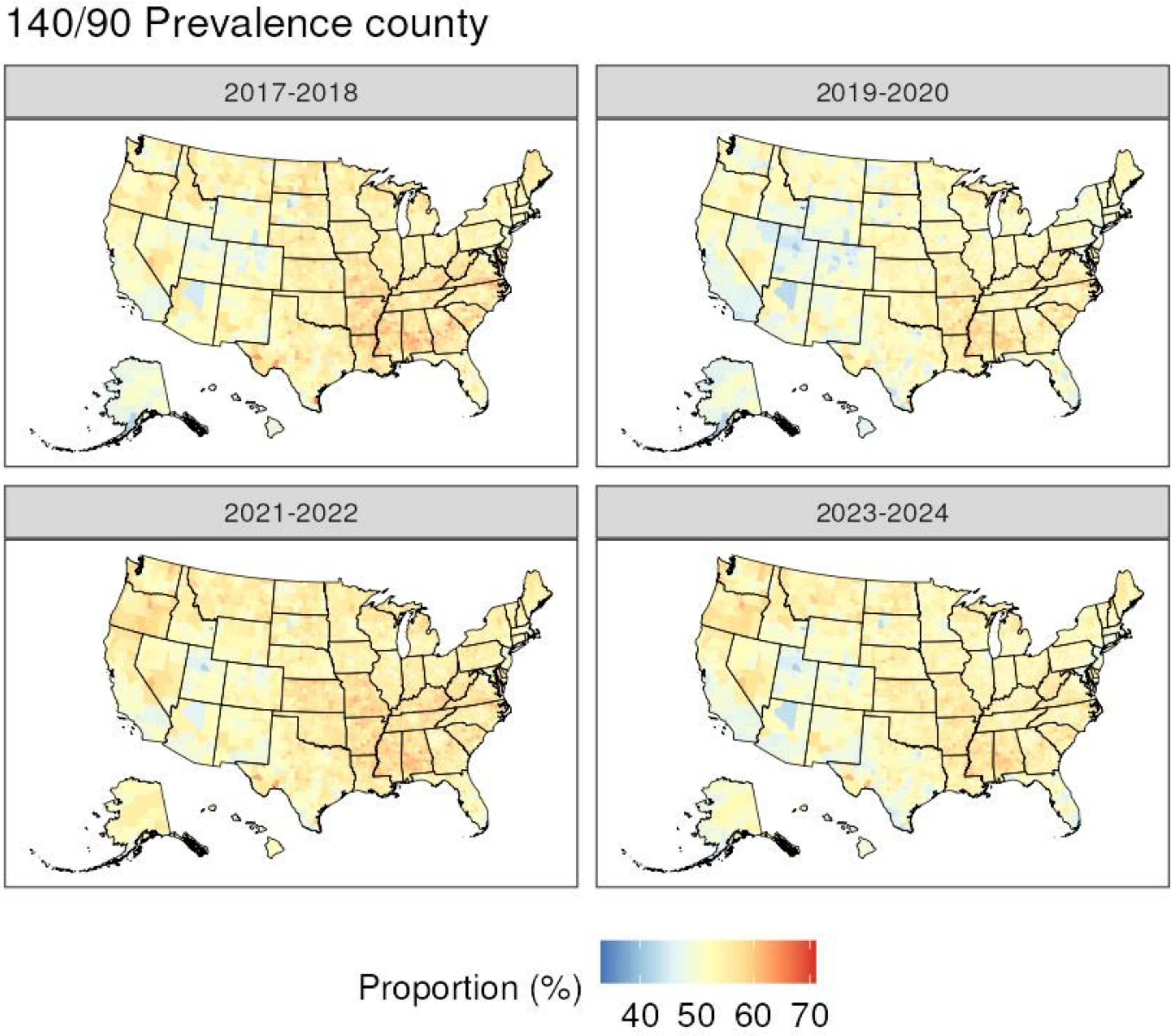

140/90 Prevalence county
2017-2018
2019-2020
2021-2022
2023-2024
Proportion (%)
40 50 60 70

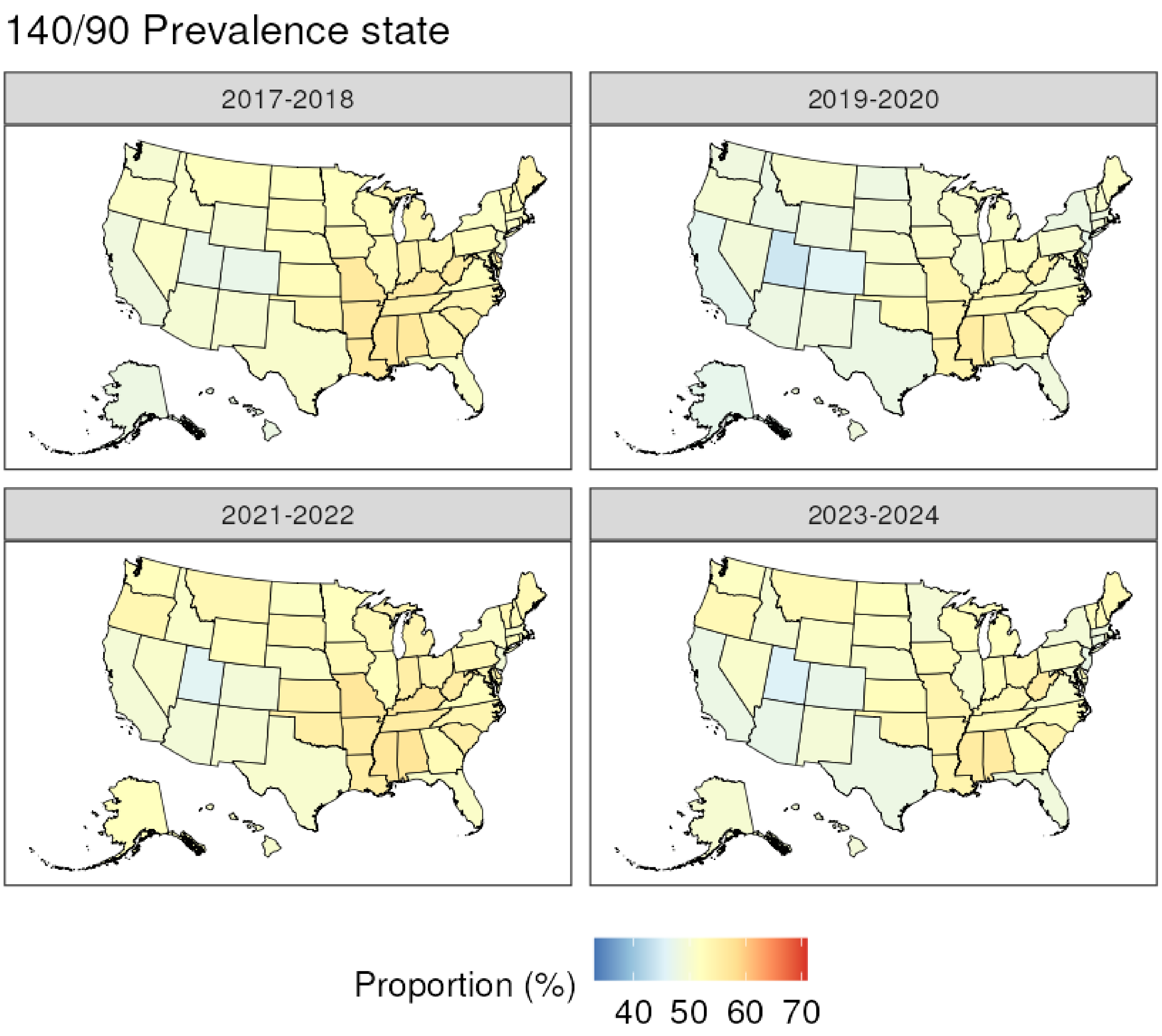
140/90 Prevalence state
2017-2018
2019-2020
2021-2022
2023-2024
Proportion (%)
40 50 60 70

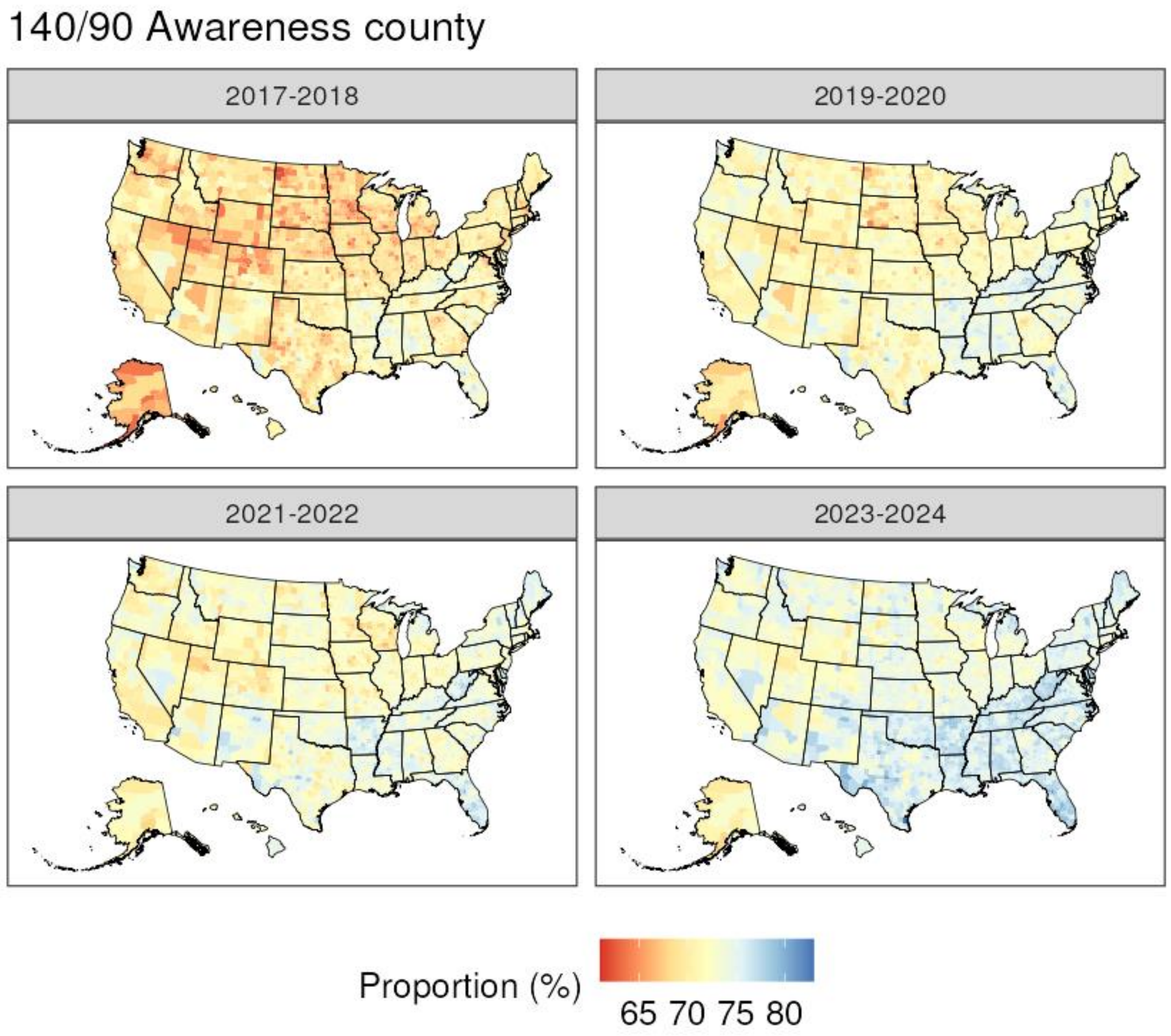

140/90 Awareness county
2017-2018
2019-2020
2021-2022
2023-2024
Proportion (%)
65 70 75 80

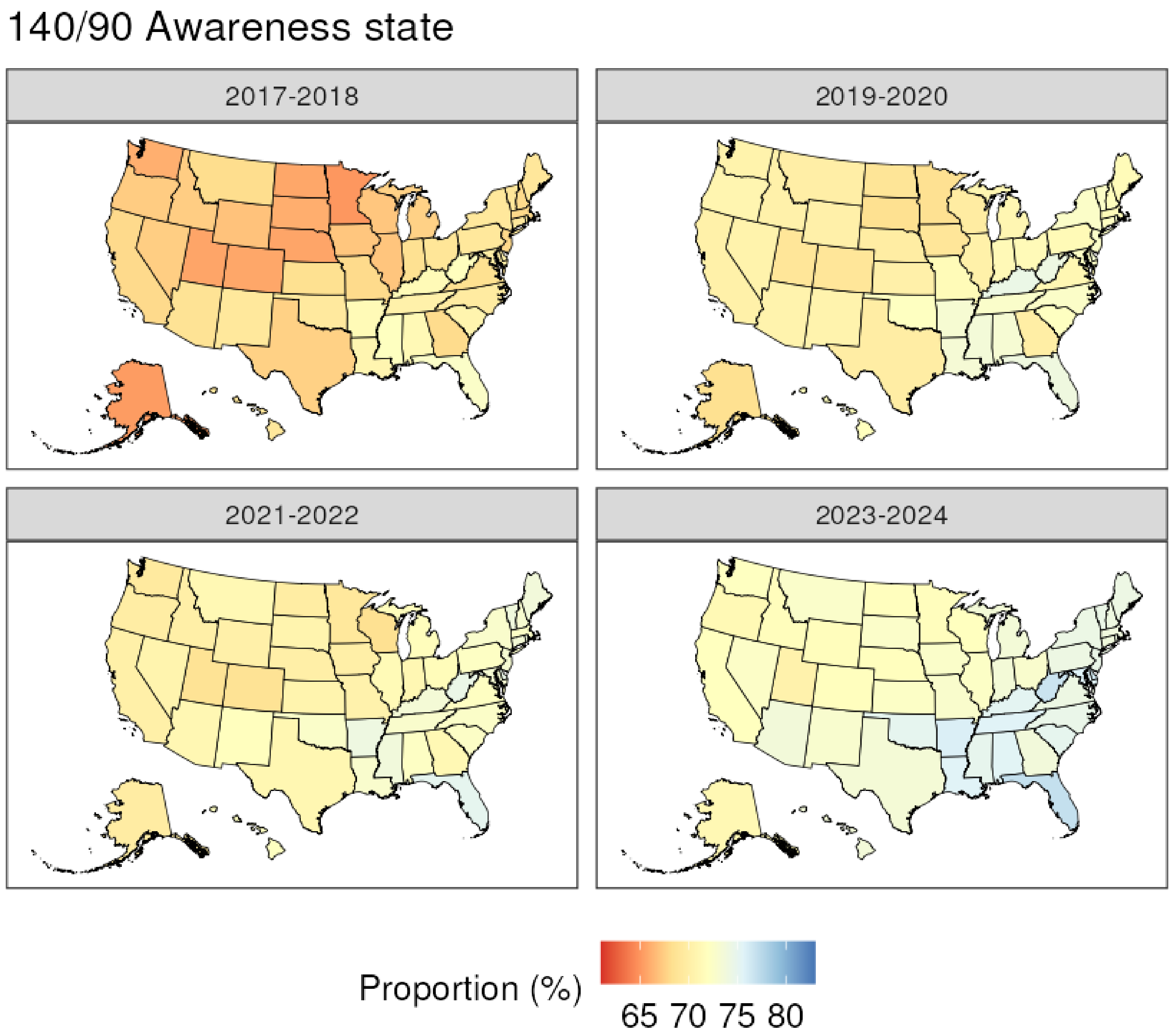
140/90 Awareness state
2017-2018
2019-2020
2021-2022
2023-2024
Proportion (%)
65 70 75 80

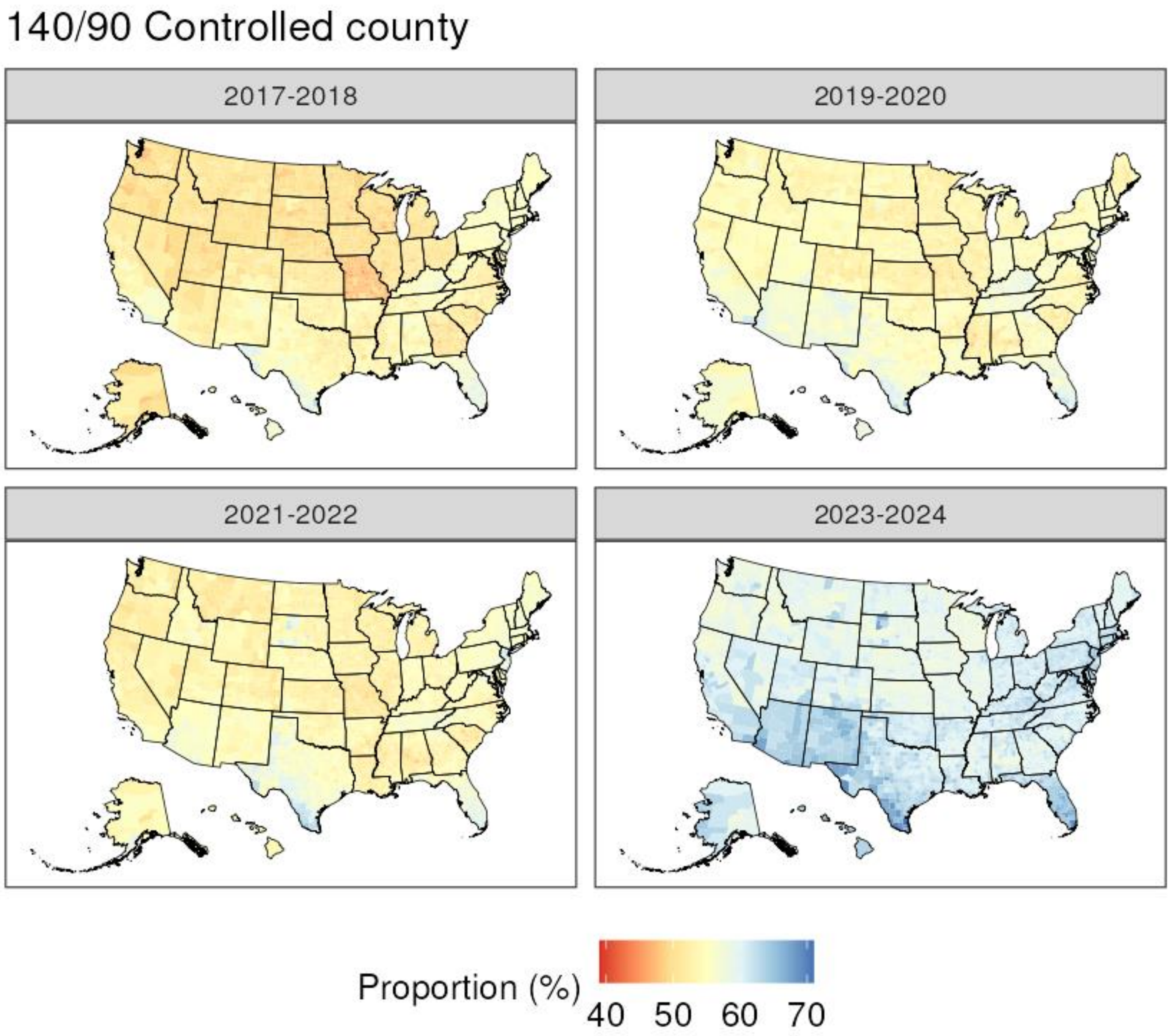
140/90 Controlled county
2017-2018
2019-2020
2021-2022
2023-2024
Proportion (%)
40 50 60 70

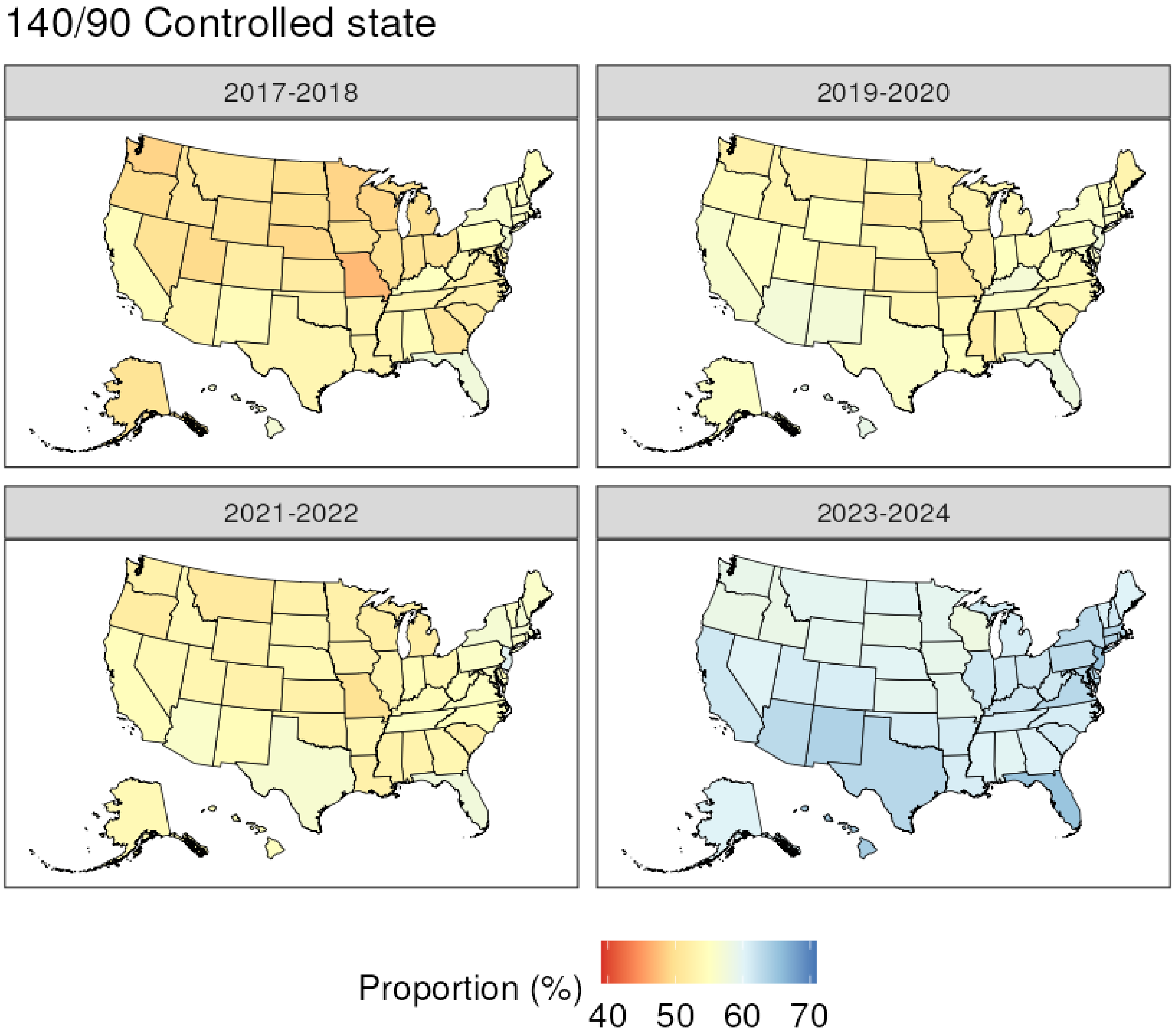
140/90 Controlled state
2017-2018
2019-2020
2021-2022
2023-2024
Proportion (%)
40
50
60
70

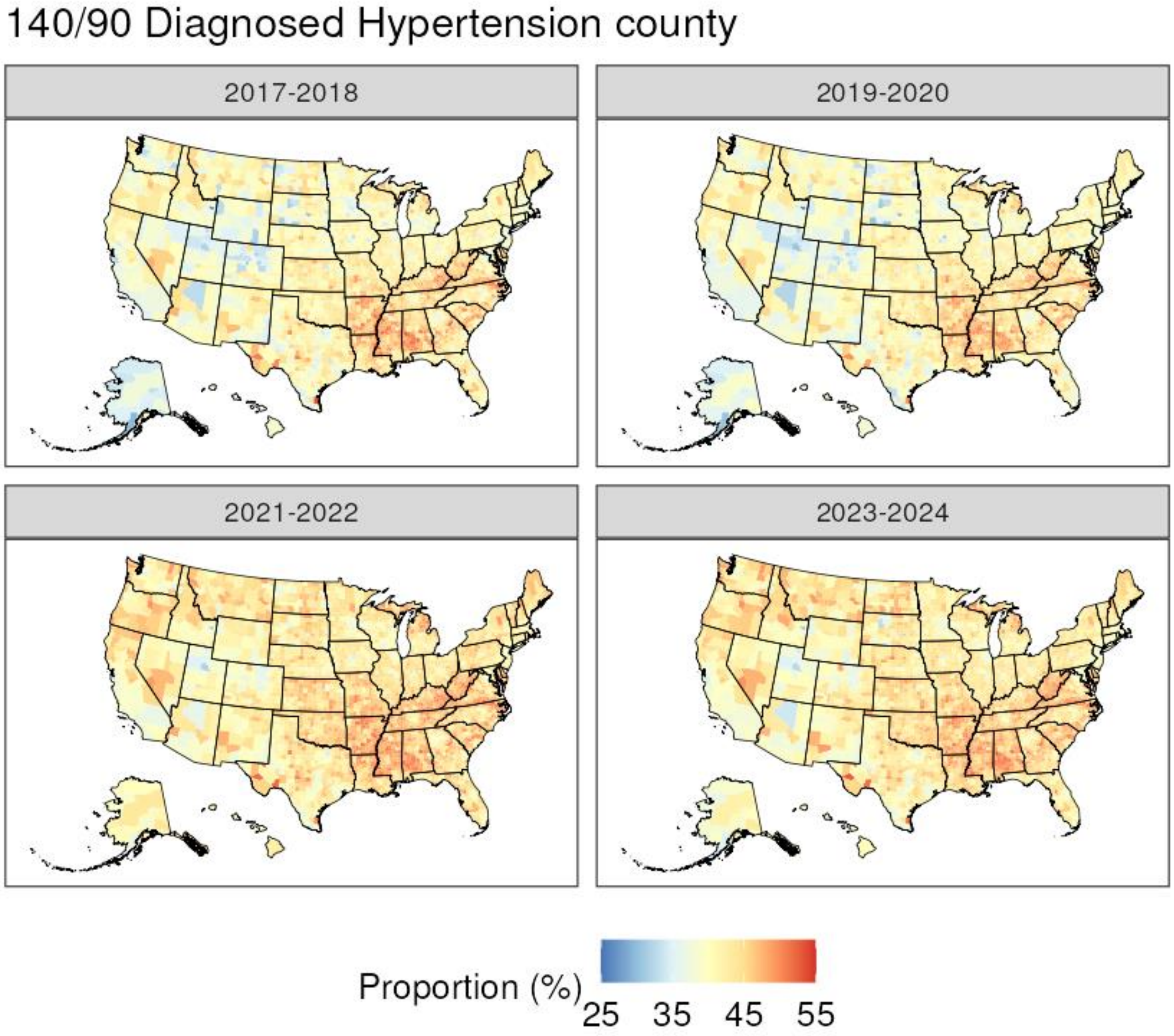

140/90 Diagnosed Hypertension county
2017-2018
2019-2020
2021-2022
2023-2024
Proportion (%)
25
35
45
55

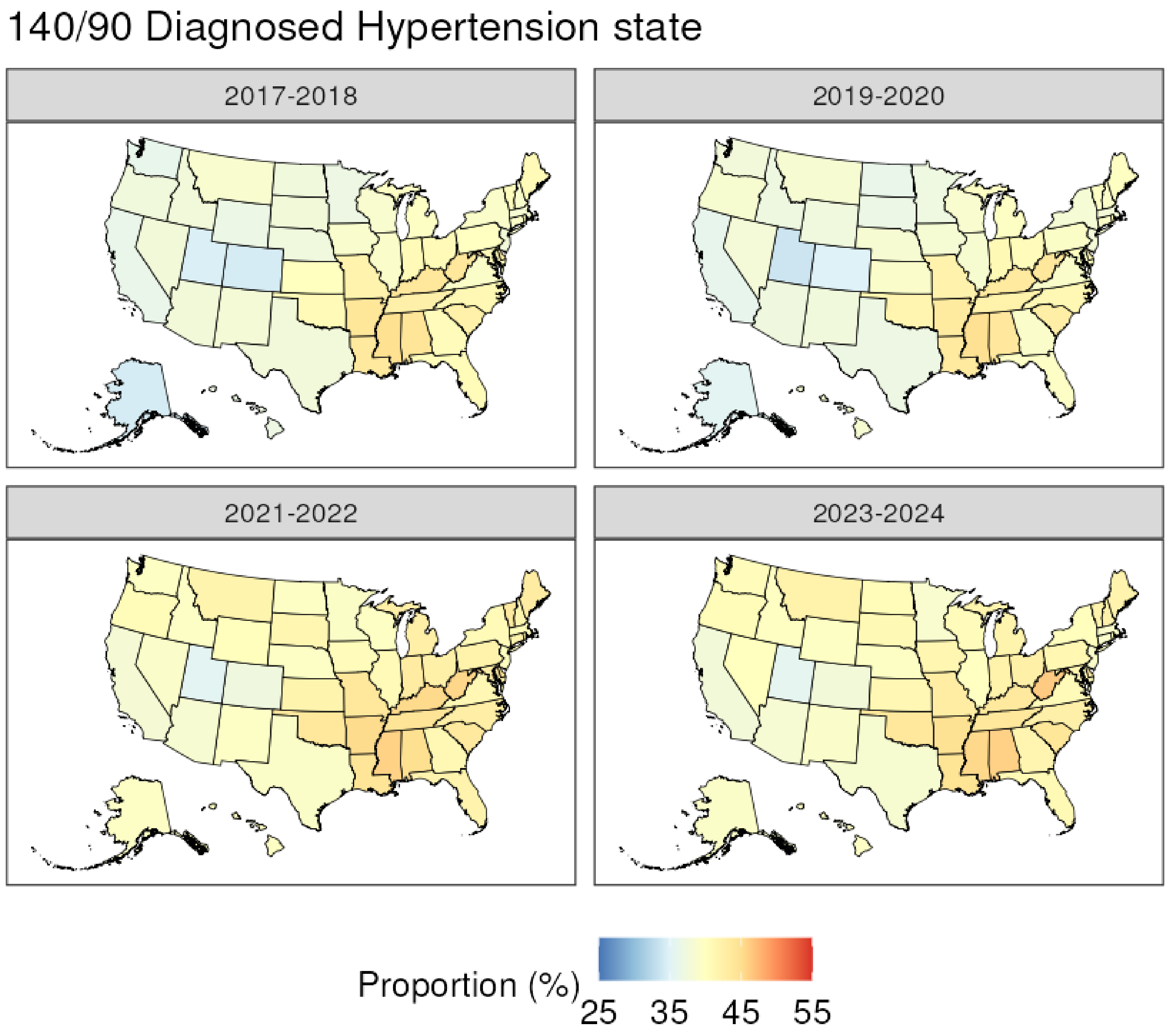
140/90 Diagnosed Hypertension state
2017-2018
2019-2020
2021-2022
2023-2024
Proportion (%)
25
35
45
55
