## Supplementary File 2 for "County Prevalence and Control of High Blood Pressure from Health Kiosks, 2017-2024"

130/80 Prevalence county

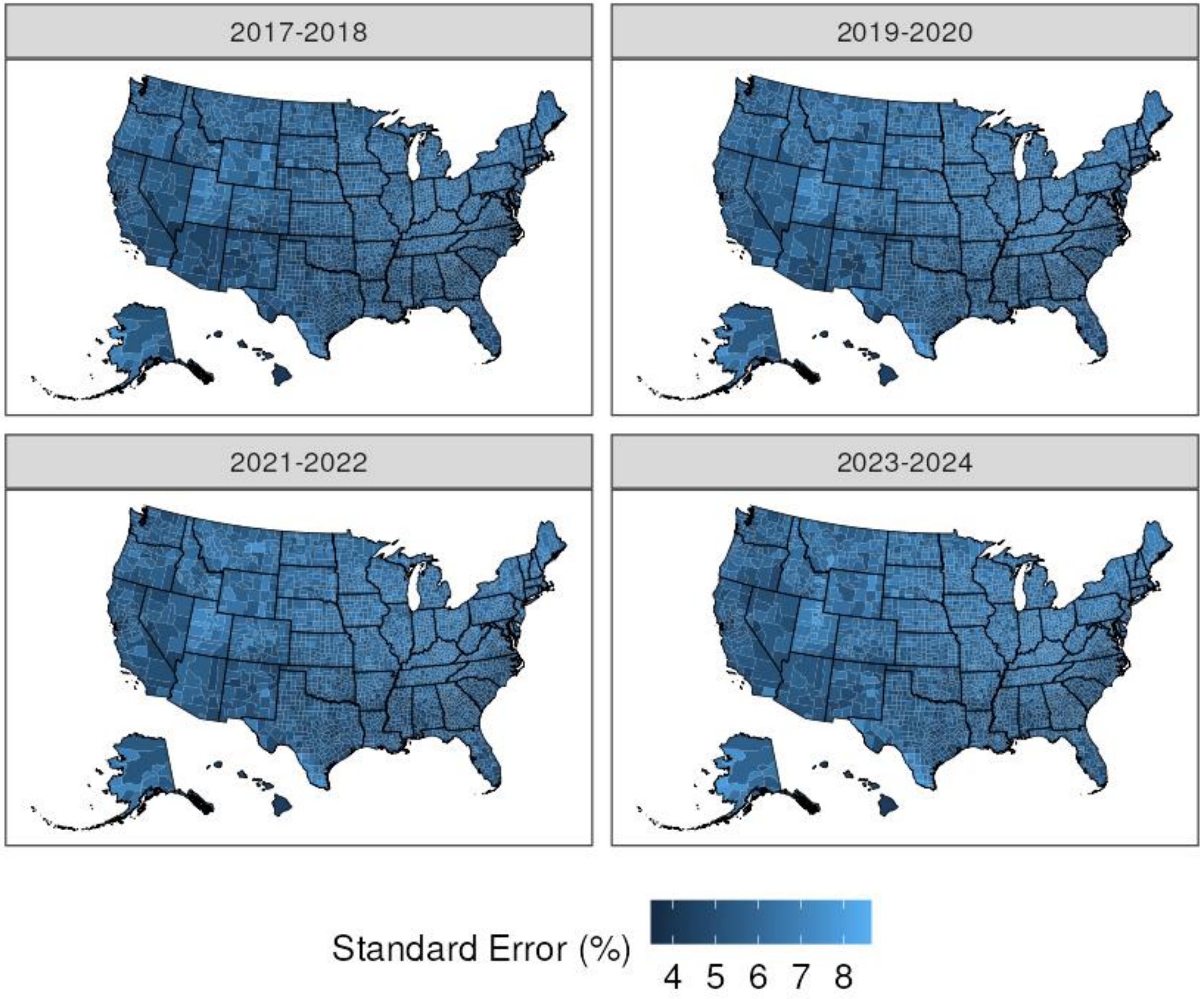

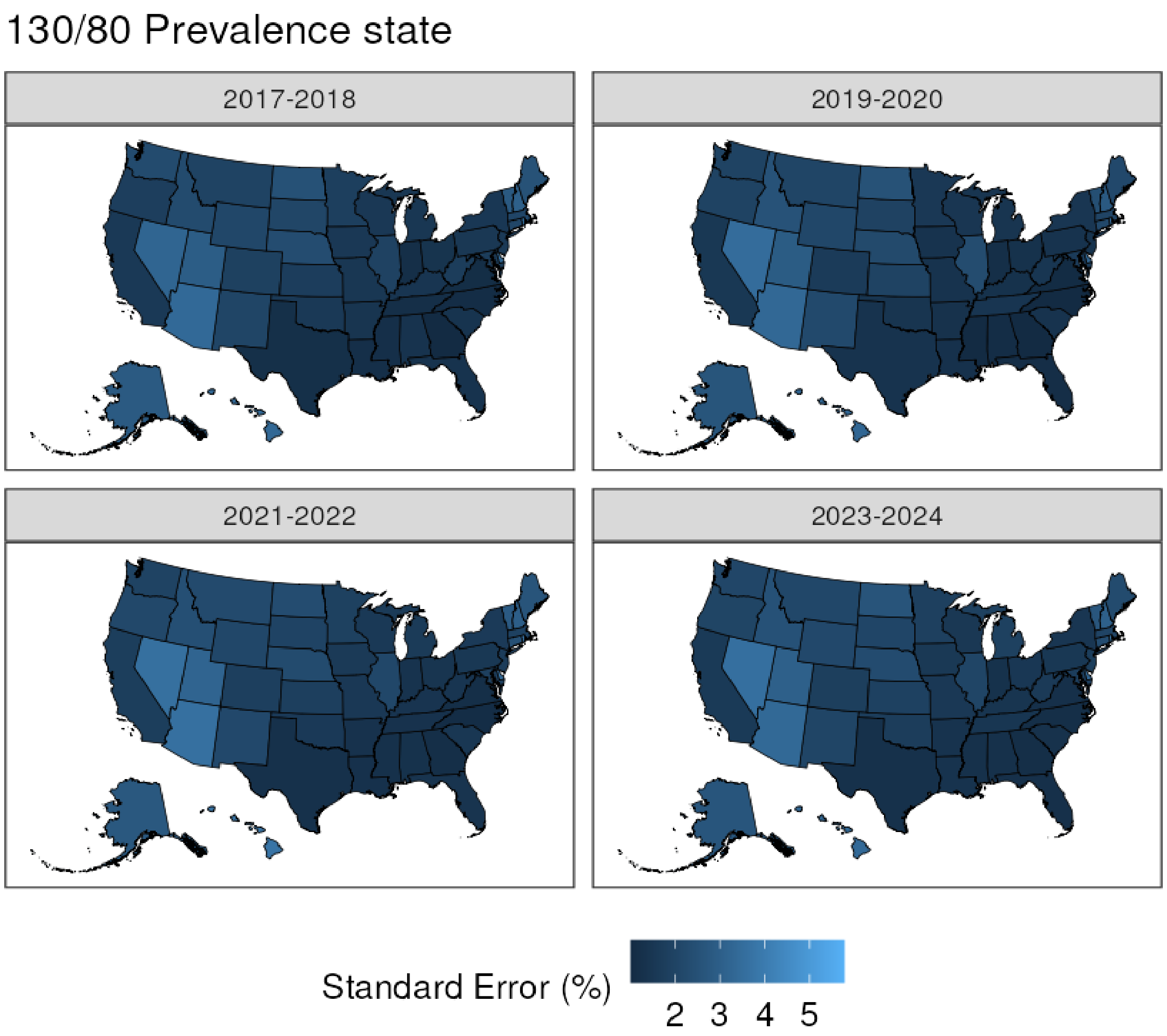
130/80 Prevalence state
2017-2018
2019-2020
2021-2022
2023-2024
Standard Error (%)
2
3
4
5

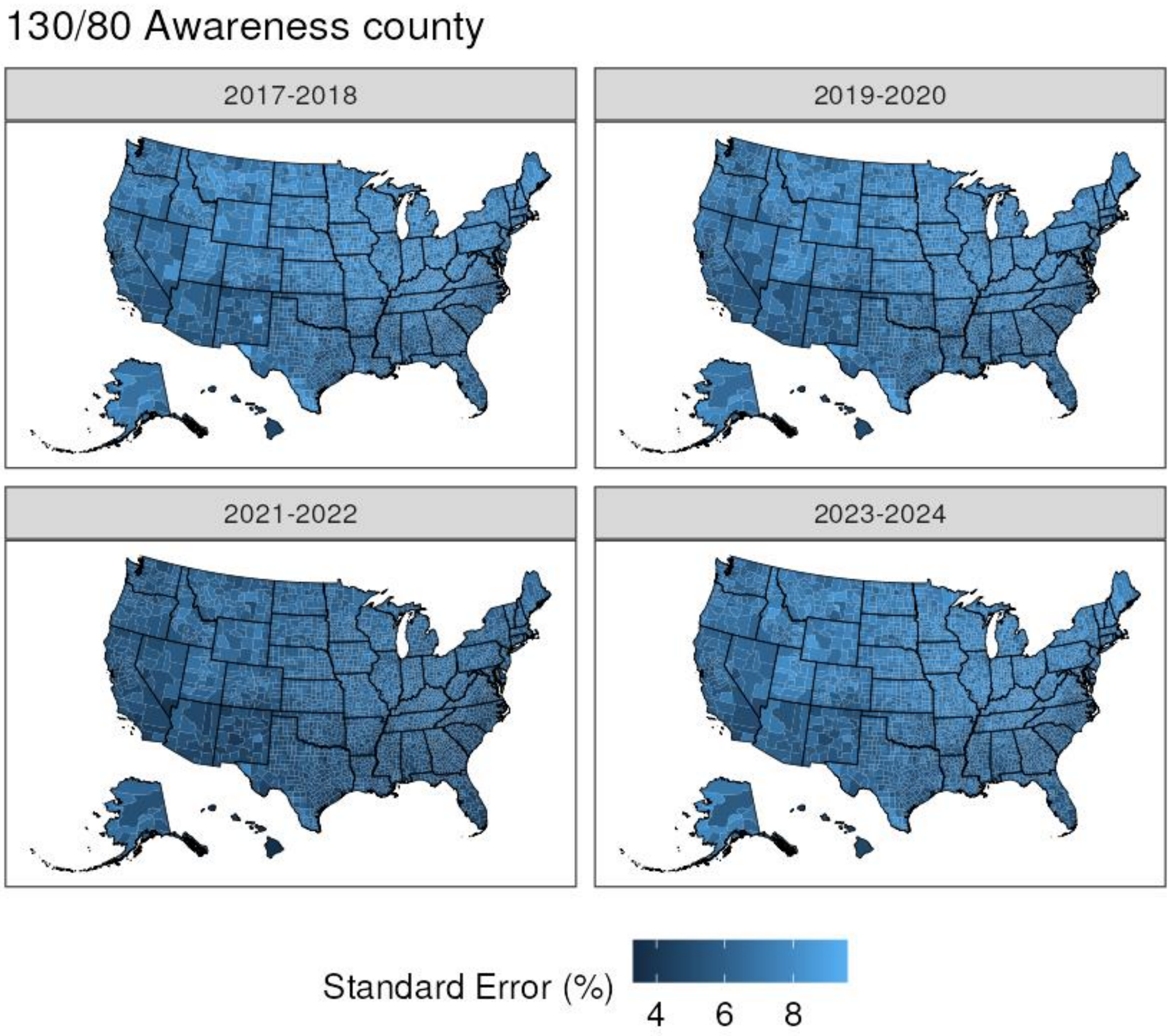

130/80 Awareness county
2017-2018
2019-2020
2021-2022
2023-2024
Standard Error (%)
4
6
8

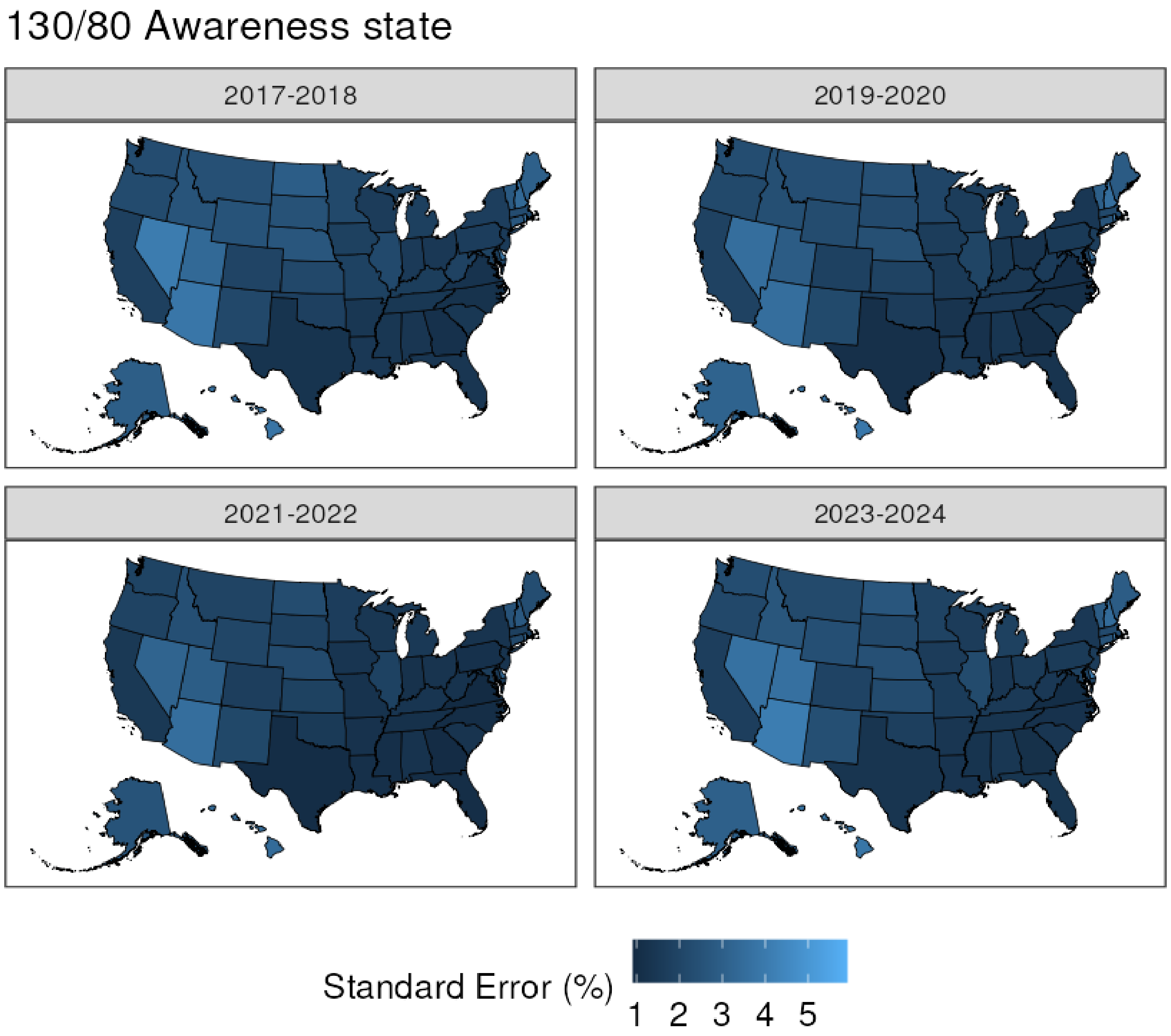
130/80 Awareness state
2017-2018
2019-2020
2021-2022
2023-2024
Standard Error (%)
1 2 3 4 5

130/80 Controlled county

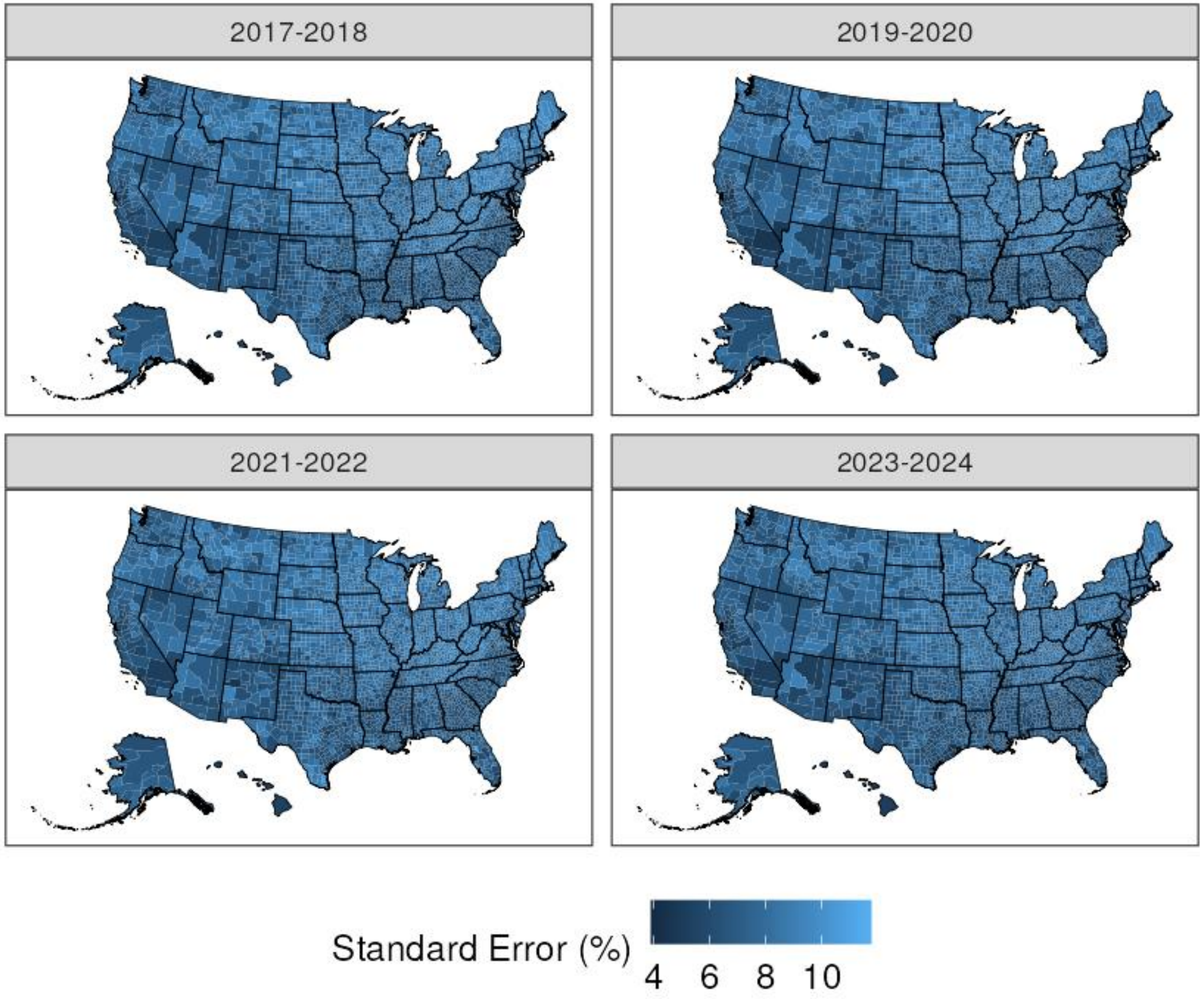

130/80 Controlled state

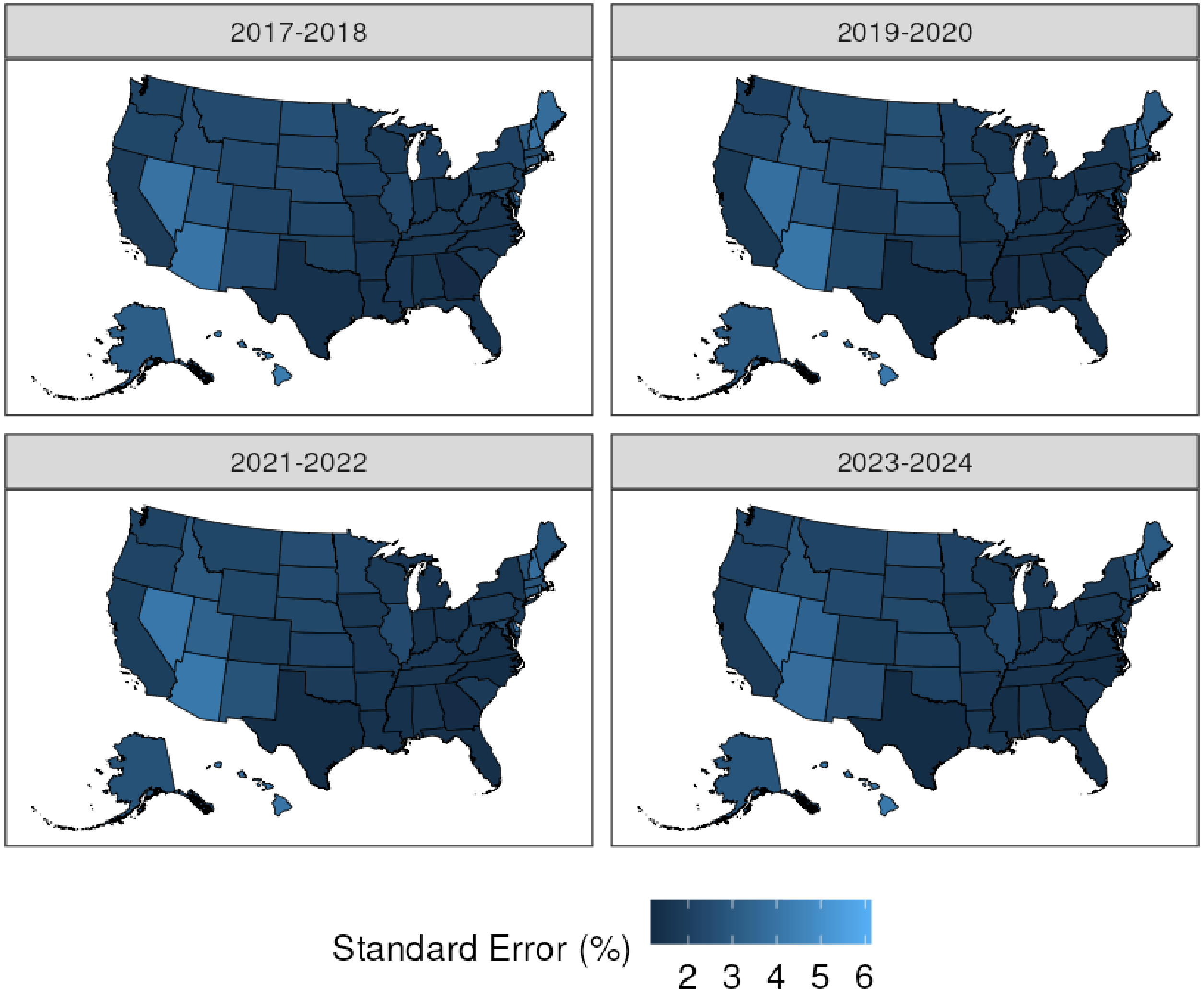

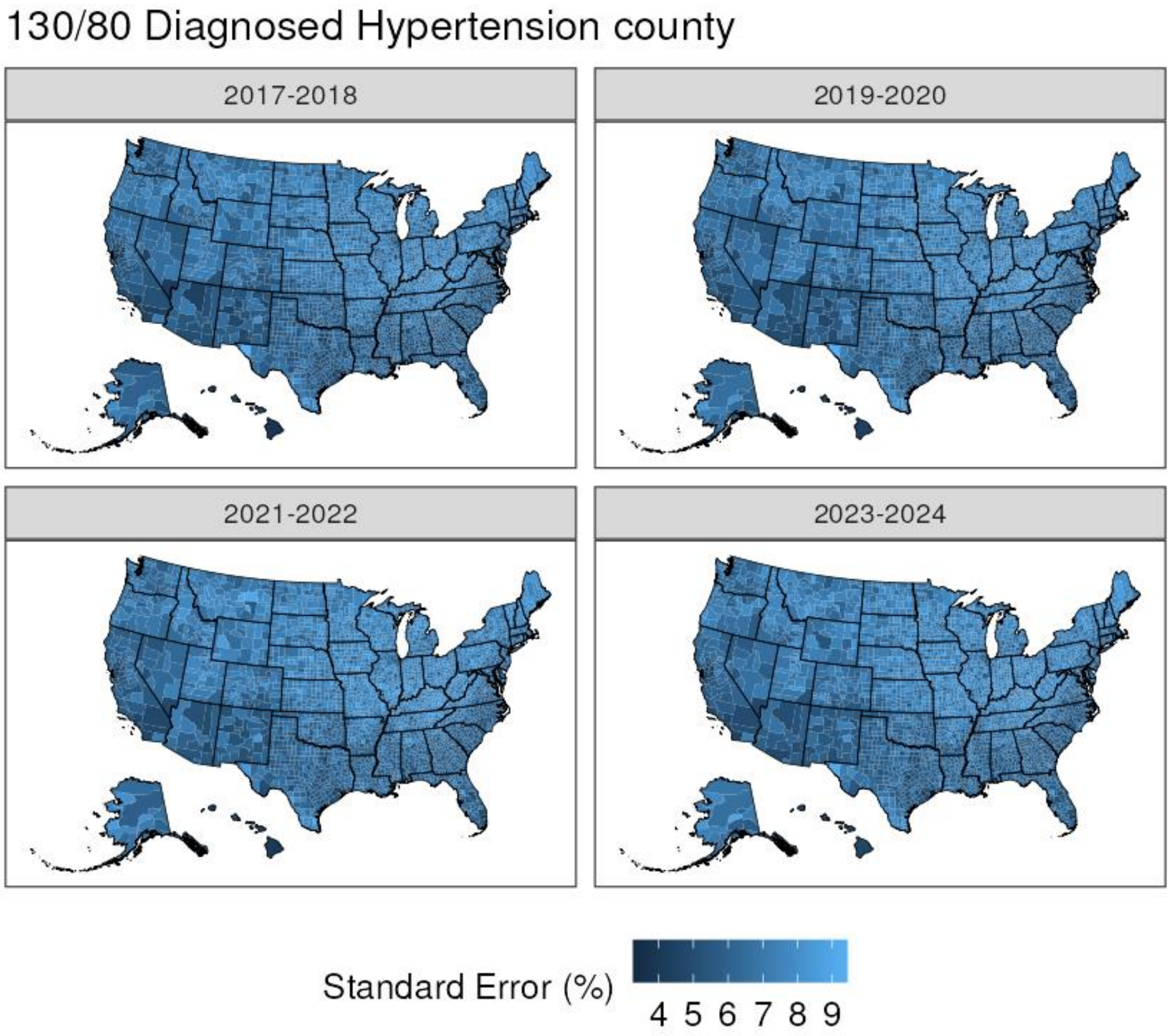
130/80 Diagnosed Hypertension county
2017-2018
2019-2020
2021-2022
2023-2024
Standard Error (%)
4 5 6 7 8 9

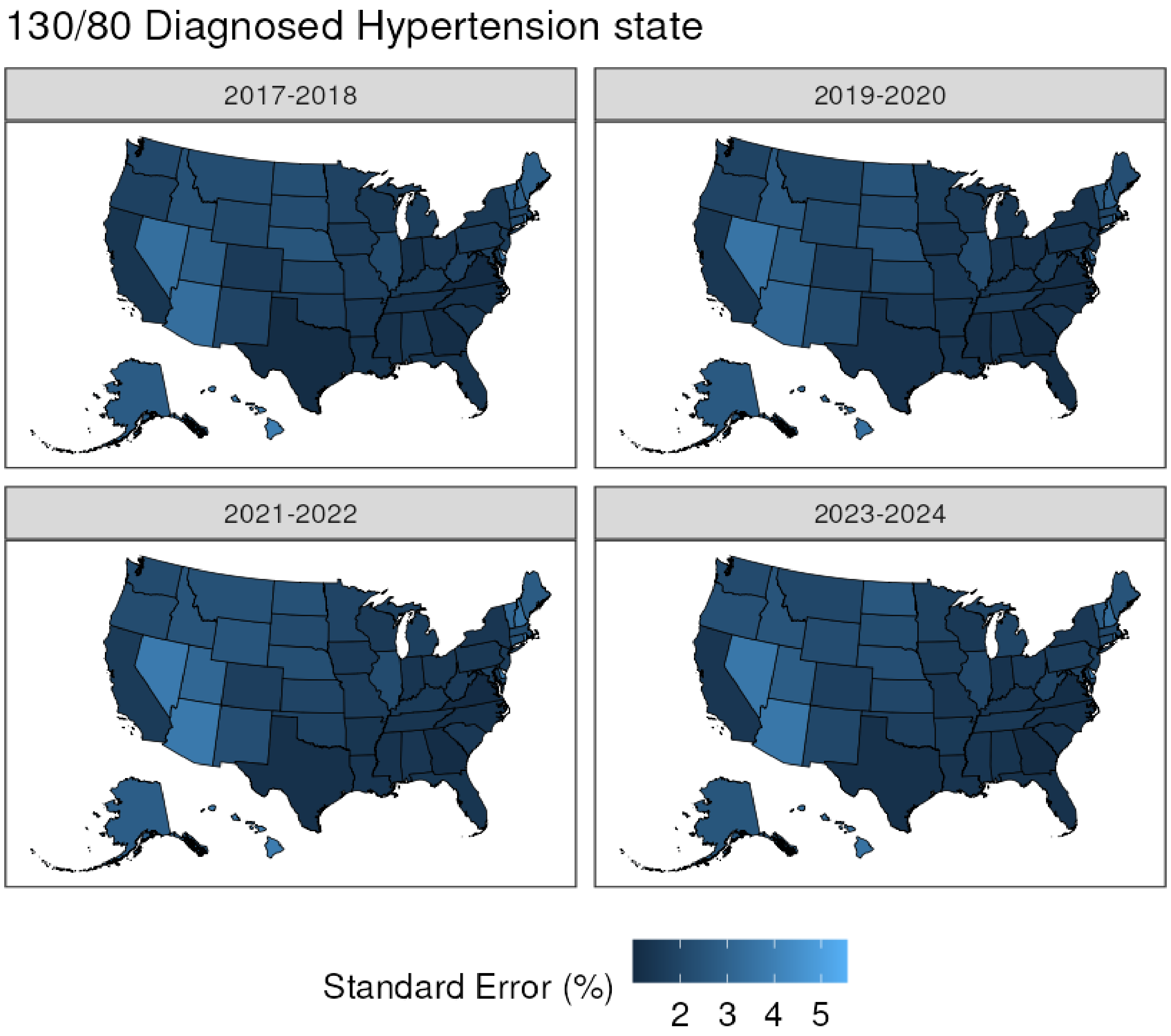

130/80 Diagnosed Hypertension state
2017-2018
2019-2020
2021-2022
2023-2024
Standard Error (%)
2
3
4
5

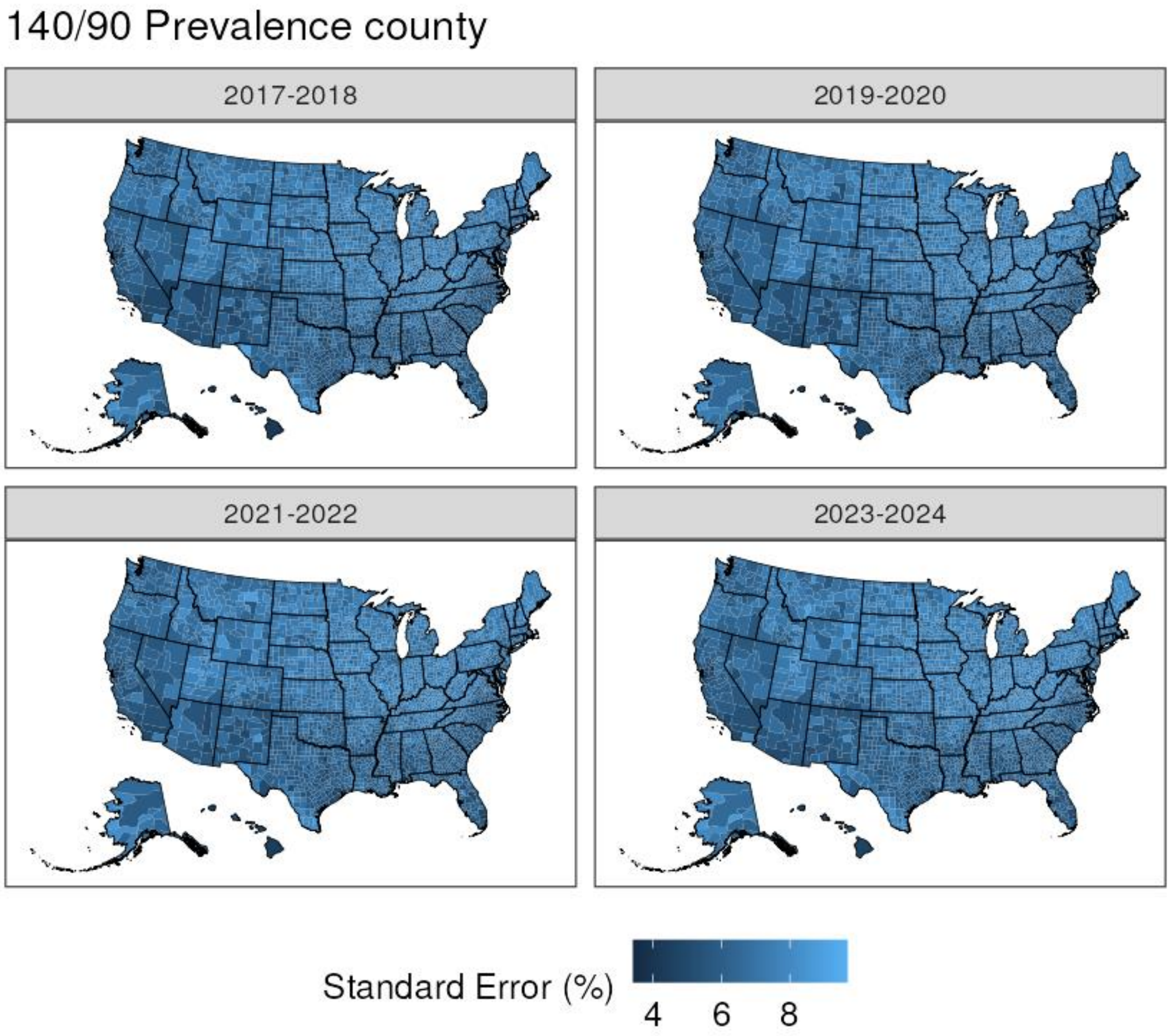
140/90 Prevalence county
2017-2018
2019-2020
2021-2022
2023-2024
Standard Error (%)
4
6
8

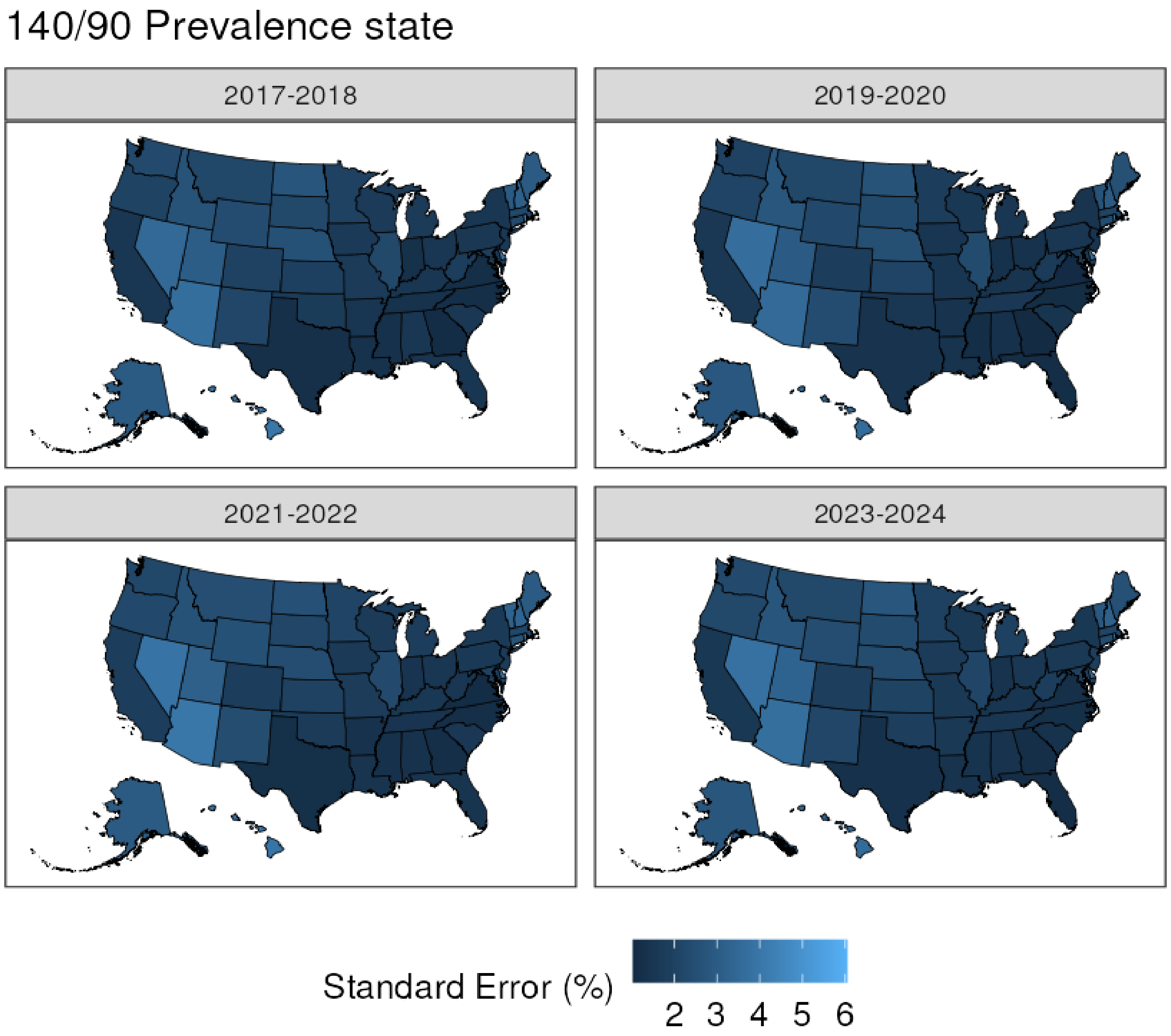
140/90 Prevalence state
2017-2018
2019-2020
2021-2022
2023-2024
Standard Error (%)
2 3 4 5 6

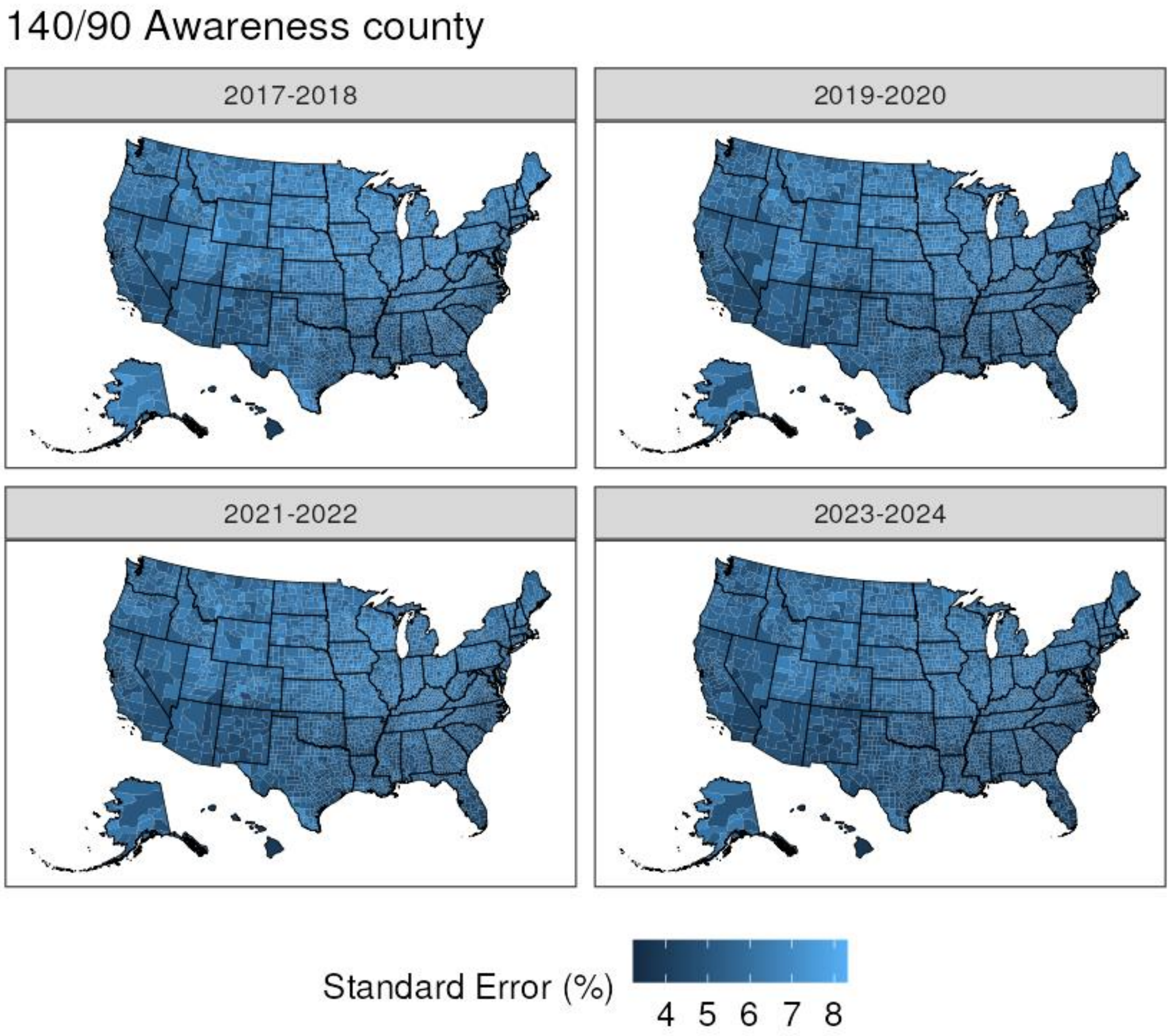

140/90 Awareness county
2017-2018
2019-2020
2021-2022
2023-2024
Standard Error (%)
4
5
6
7
8

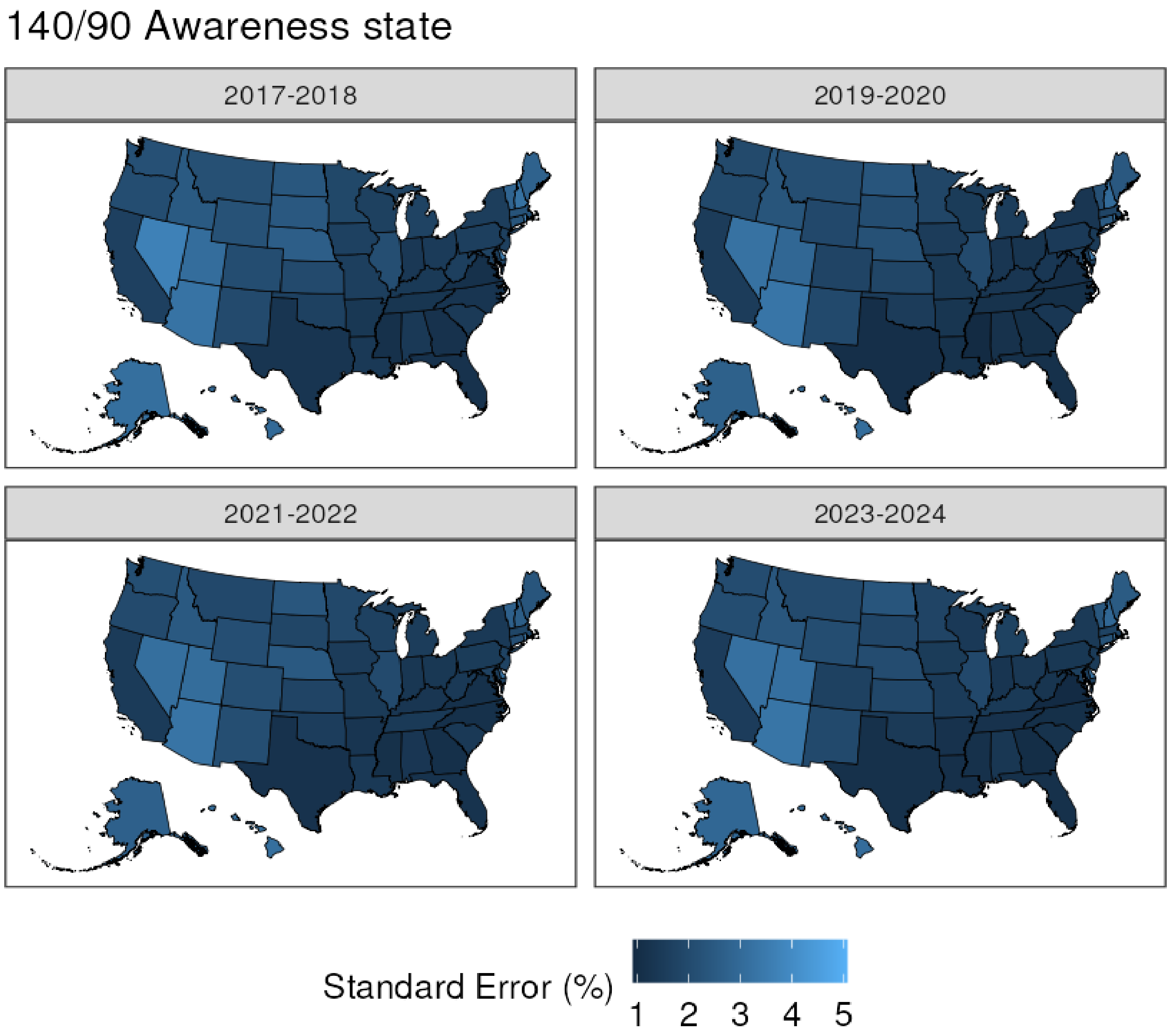
140/90 Awareness state
2017-2018
2019-2020
2021-2022
2023-2024
Standard Error (%)
1
2
3
4
5

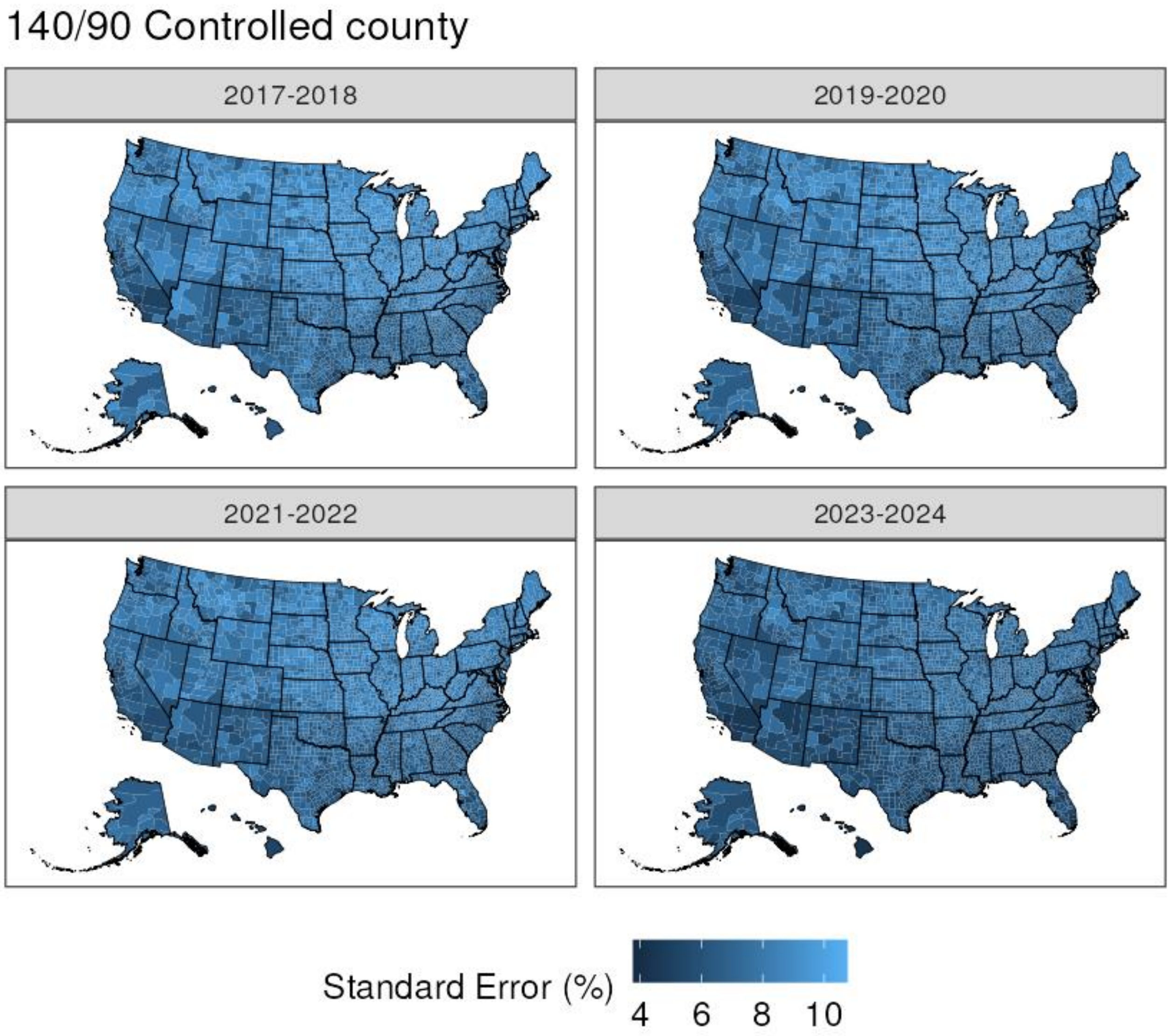
140/90 Controlled county
2017-2018
2019-2020
2021-2022
2023-2024
Standard Error (%)
4
6
8
10

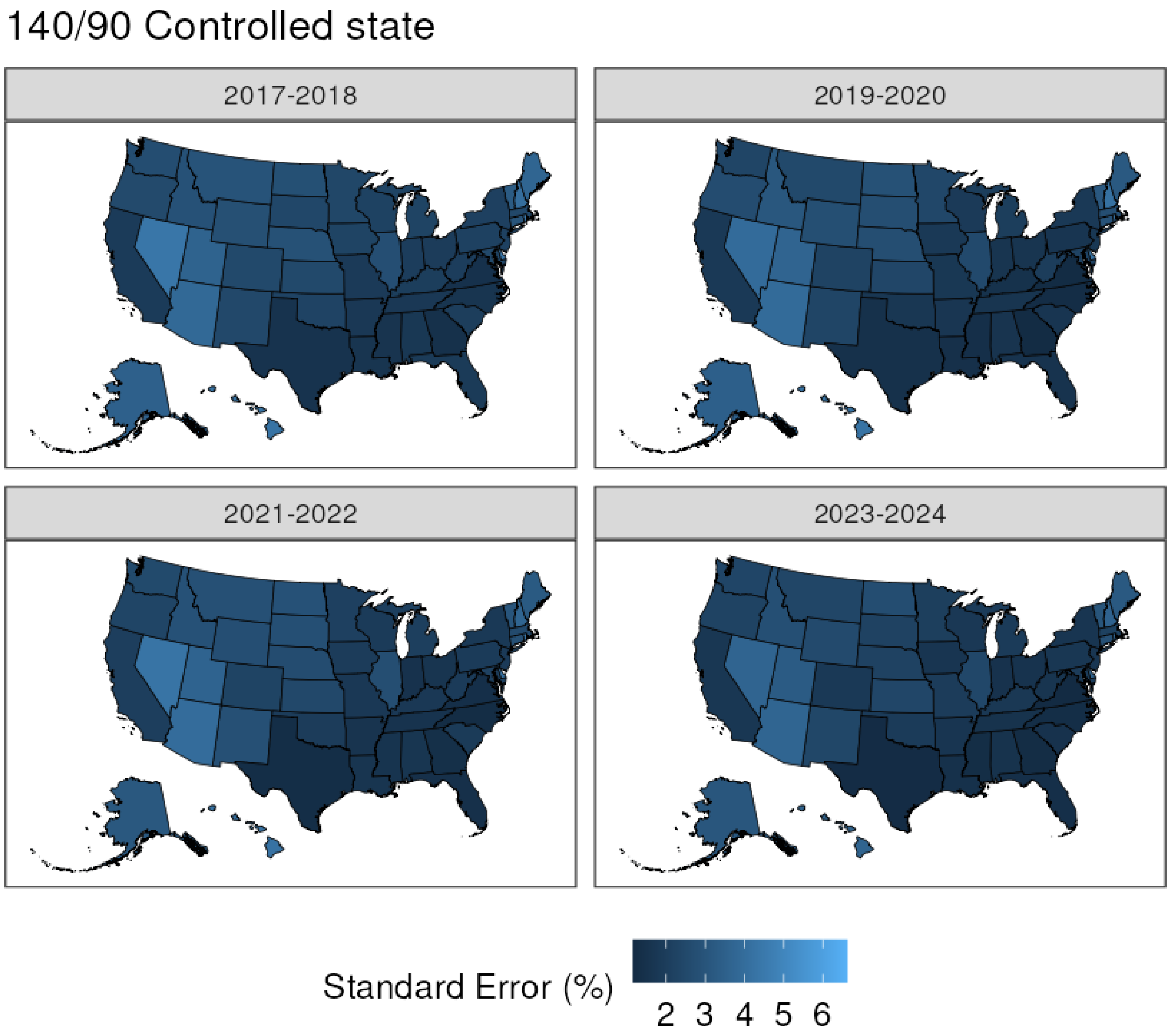
140/90 Controlled state
2017-2018
2019-2020
2021-2022
2023-2024
Standard Error (%)
2 3 4 5 6

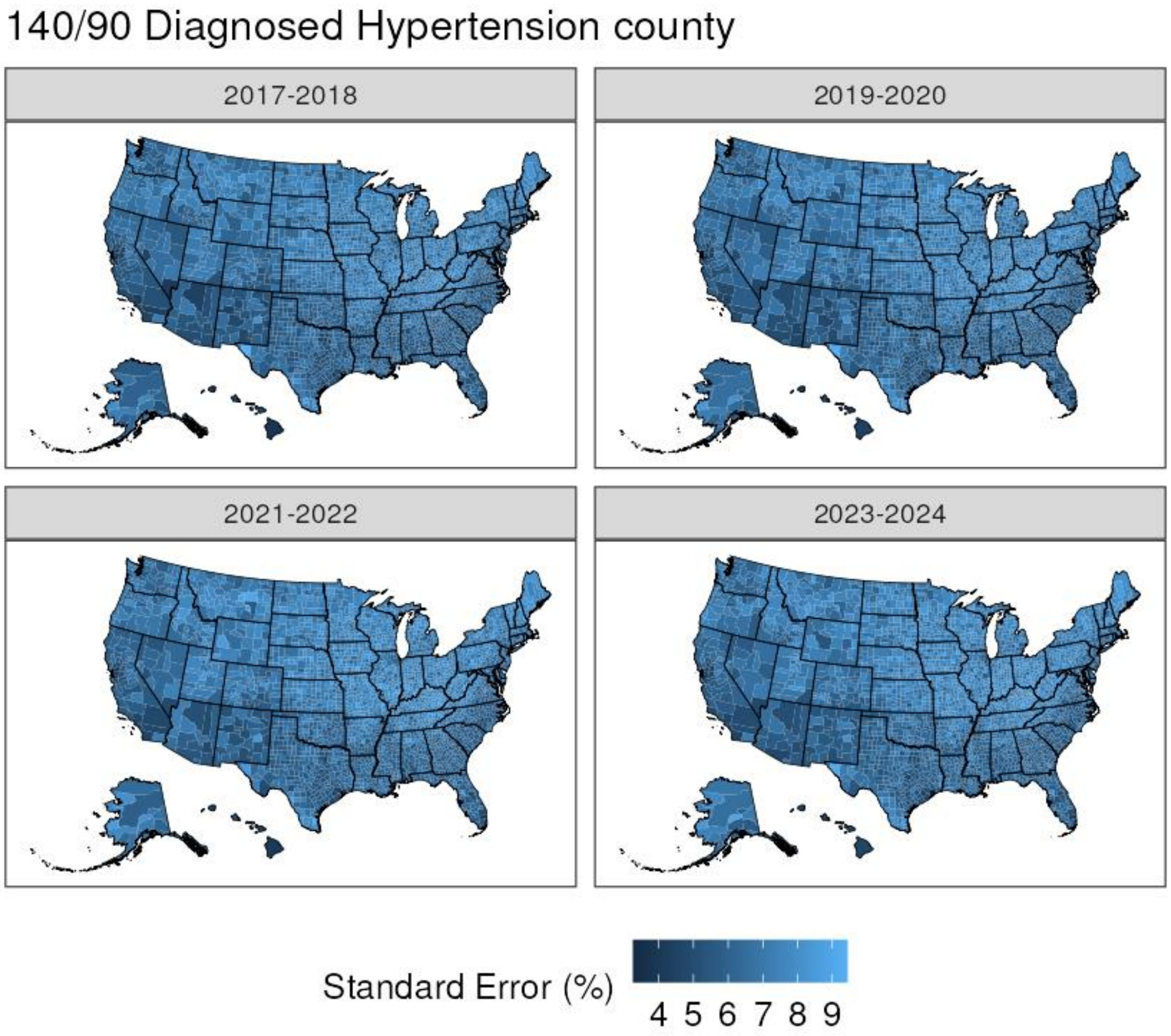
140/90 Diagnosed Hypertension county
2017-2018
2019-2020
2021-2022
2023-2024
Standard Error (%)
4 5 6 7 8 9

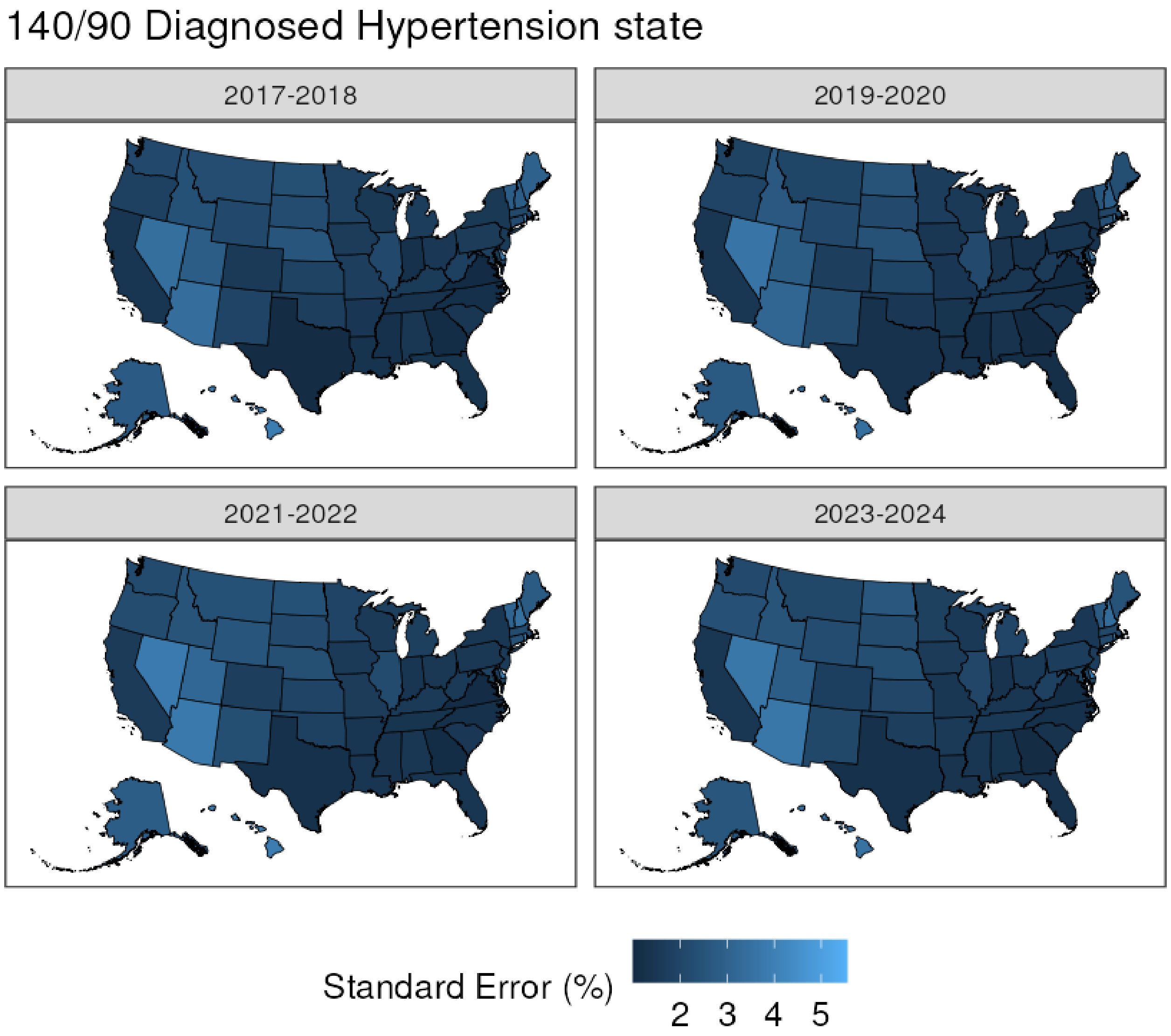
140/90 Diagnosed Hypertension state
2017-2018
2019-2020
2021-2022
2023-2024
Standard Error (%)
2
3
4
5
