## Supplementary File 3 for "County Prevalence and Control of High Blood Pressure from Health Kiosks, 2017-2024"

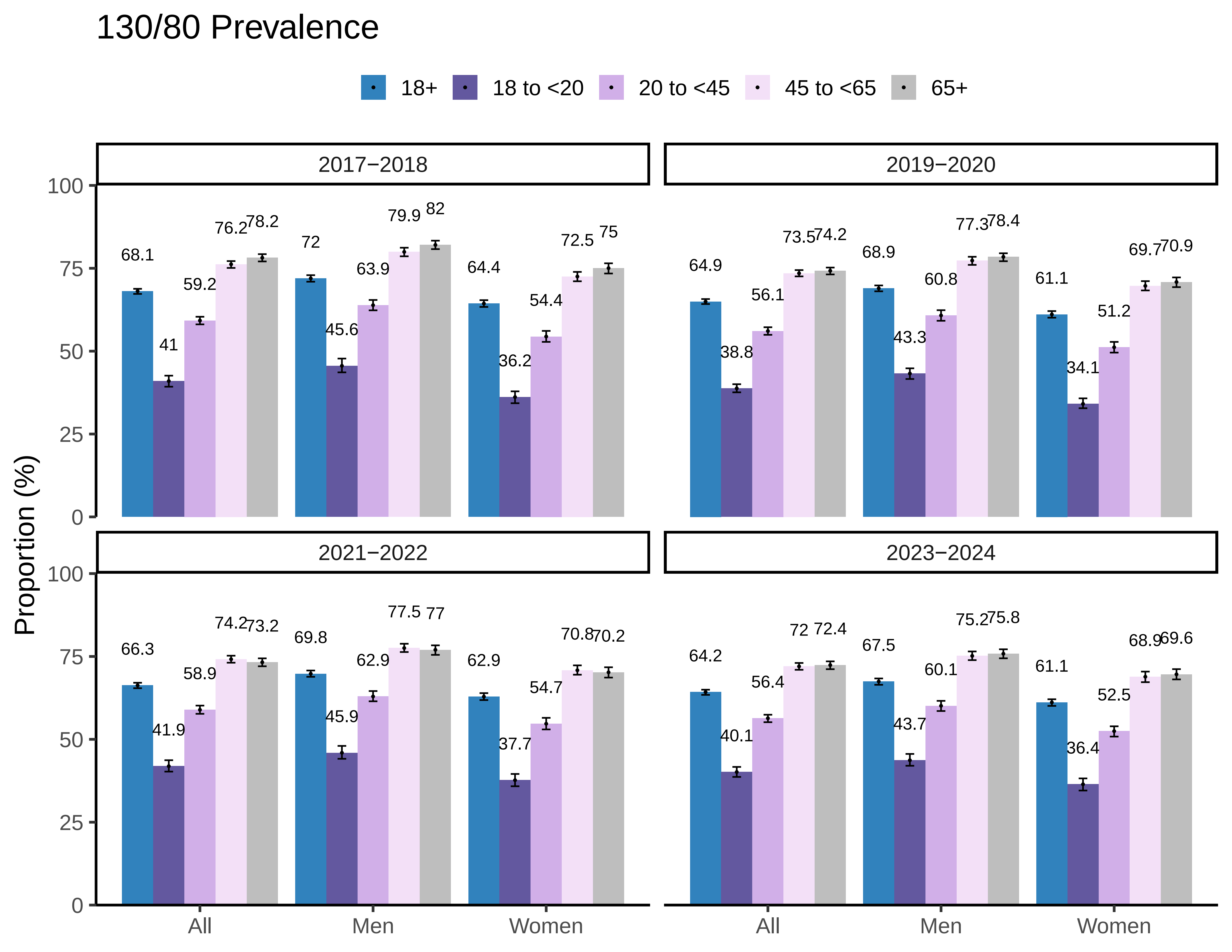
130/80 Prevalence
18+
18 to <20
20 to <45
45 to <65
65+
2017−2018
2019−2020
2021−2022
2023−2024
Proportion (%)
0
25
50
75
100
All
Men
Women
68.1
41
59.2
76.2
78.2
72
45.6
63.9
79.9
82
64.4
36.2
54.4
72.5
75
64.9
38.8
56.1
73.5
74.2
68.9
43.3
60.8
77.3
78.4
61.1
34.1
51.2
69.7
70.9
66.3
41.9
58.9
74.2
73.2
69.8
45.9
62.9
77.5
77
62.9
37.7
54.7
70.8
70.2
64.2
40.1
56.4
72
72.4
67.5
43.7
60.1
75.2
75.8
61.1
36.4
52.5
68.9
69.6

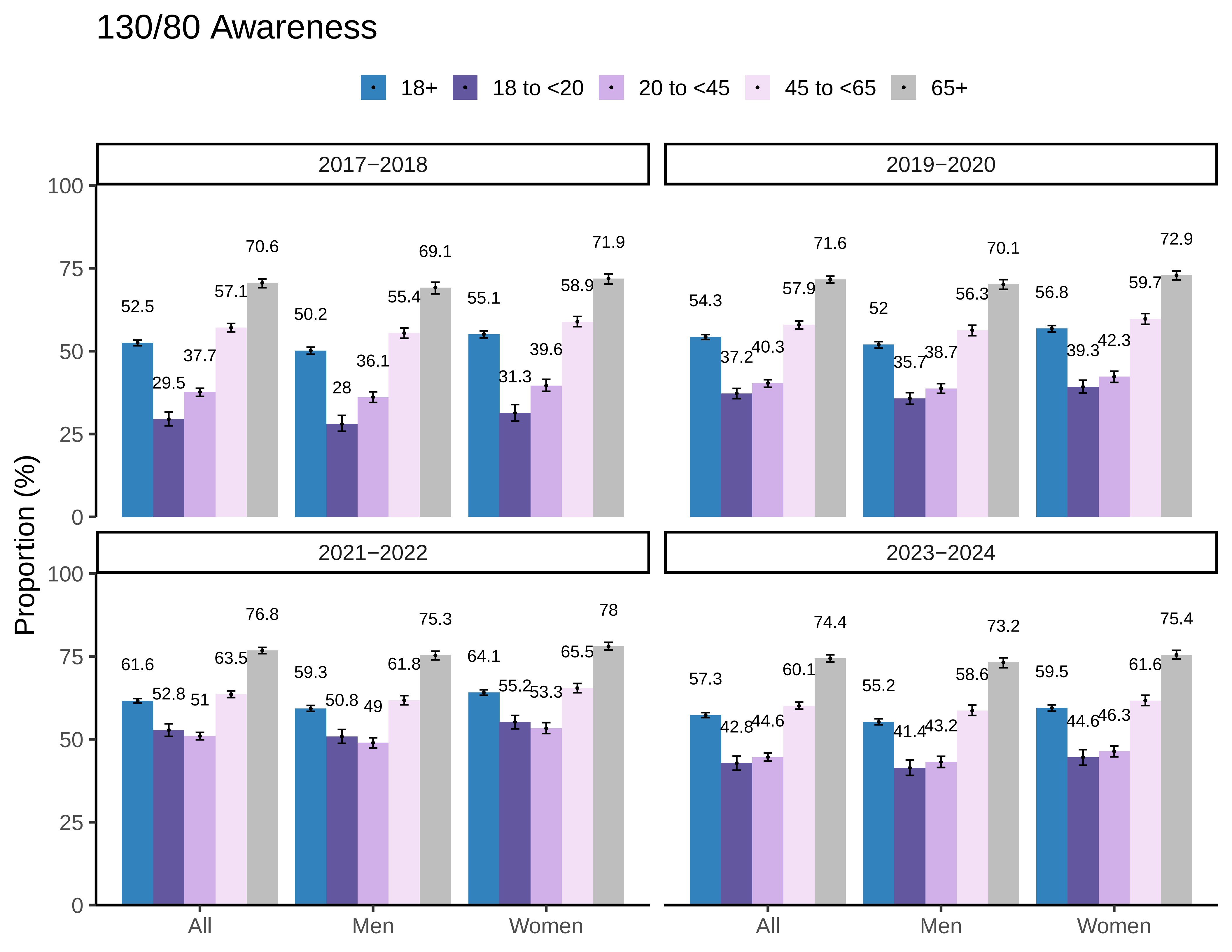

130/80 Awareness
18+
18 to <20
20 to <45
45 to <65
65+
2017−2018
2019−2020
2021−2022
2023−2024
Proportion (%)
100
75
50
25
0
52.5
29.5
37.7
57.1
70.6
50.2
28
36.1
55.4
69.1
55.1
31.3
39.6
58.9
71.9
54.3
37.2
40.3
57.9
71.6
52
35.7
38.7
56.3
70.1
56.8
39.3
42.3
59.7
72.9
61.6
52.8
51
63.5
76.8
59.3
50.8
49
61.8
75.3
64.1
55.2
53.3
65.5
78
57.3
42.8
44.6
60.1
74.4
55.2
41.4
43.2
58.6
73.2
59.5
44.6
46.3
61.6
75.4
All
Men
Women

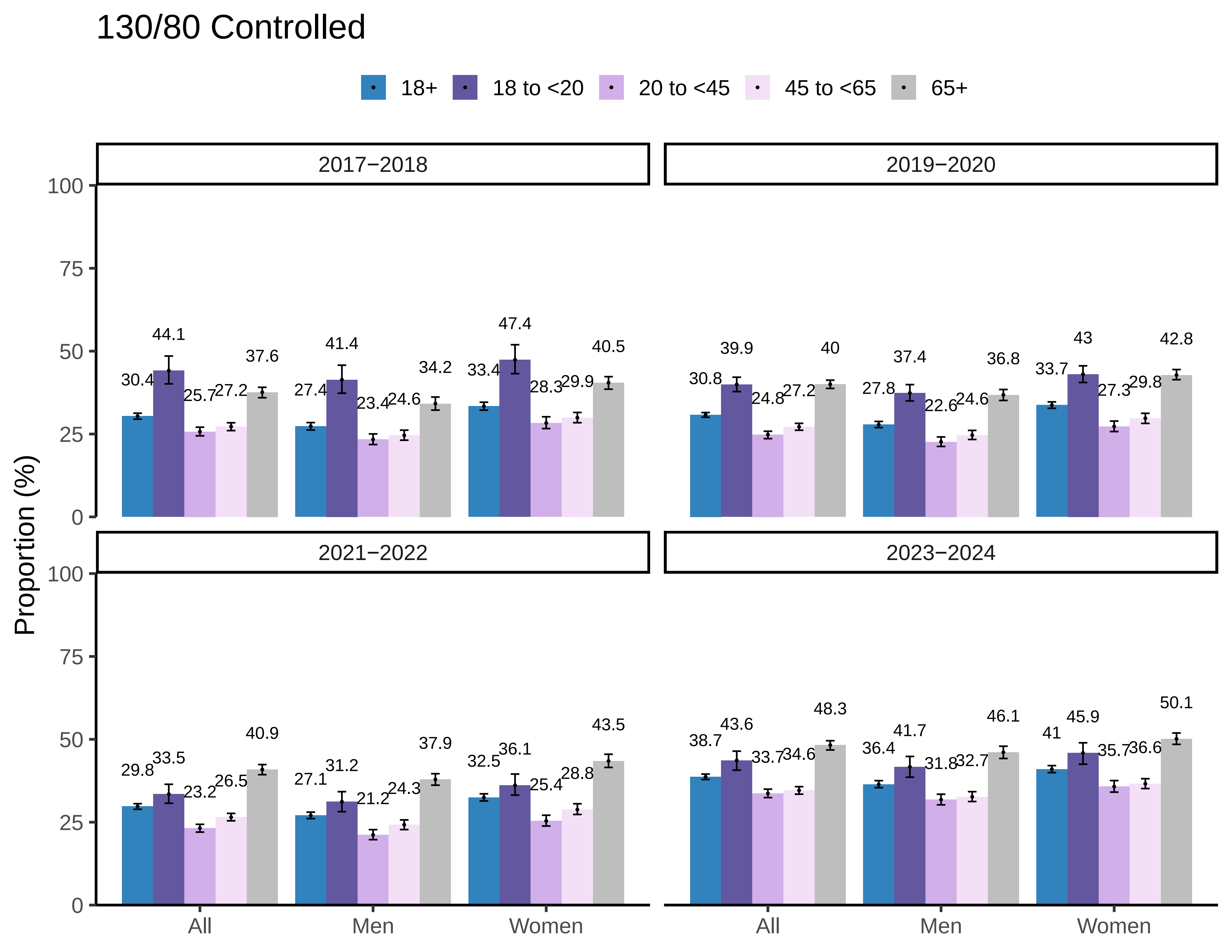
130/80 Controlled
18+
18 to <20
20 to <45
45 to <65
65+
2017−2018
2019−2020
2021−2022
2023−2024
Proportion (%)
0
25
50
75
100
All
Men
Women
30.4
44.1
25.7
27.2
37.6
27.4
41.4
23.4
24.6
34.2
33.4
47.4
28.3
29.9
40.5
30.8
39.9
24.8
27.2
40
27.8
37.4
22.6
24.6
36.8
33.7
43
27.3
29.8
42.8
29.8
33.5
23.2
26.5
40.9
27.1
31.2
21.2
24.3
37.9
32.5
36.1
25.4
28.8
43.5
38.7
43.6
33.7
34.6
48.3
36.4
41.7
31.8
32.7
46.1
41
45.9
35.7
36.6
50.1

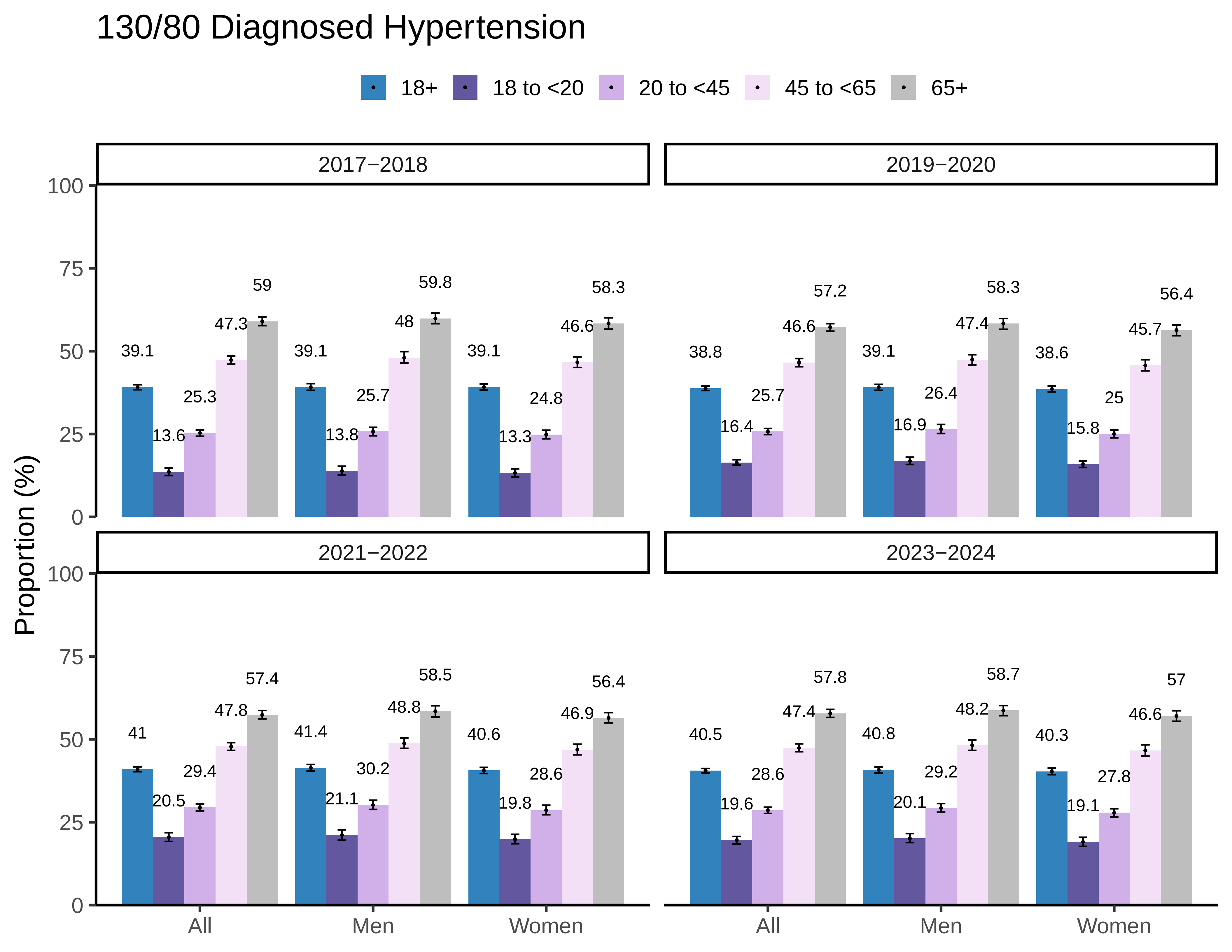

130/80 Diagnosed Hypertension
18+
18 to <20
20 to <45
45 to <65
65+
Proportion (%)
2017−2018
2019−2020
2021−2022
2023−2024
0
25
50
75
100
All
Men
Women
39.1
13.6
25.3
47.3
59
39.1
13.8
25.7
48
59.8
39.1
13.3
24.8
46.6
58.3
38.8
16.4
25.7
46.6
57.2
39.1
16.9
26.4
47.4
58.3
38.6
15.8
25
45.7
56.4
41
20.5
29.4
47.8
57.4
41.4
21.1
30.2
48.8
58.5
40.6
19.8
28.6
46.9
56.4
40.5
19.6
28.6
47.4
57.8
40.8
20.1
29.2
48.2
58.7
40.3
19.1
27.8
46.6
57

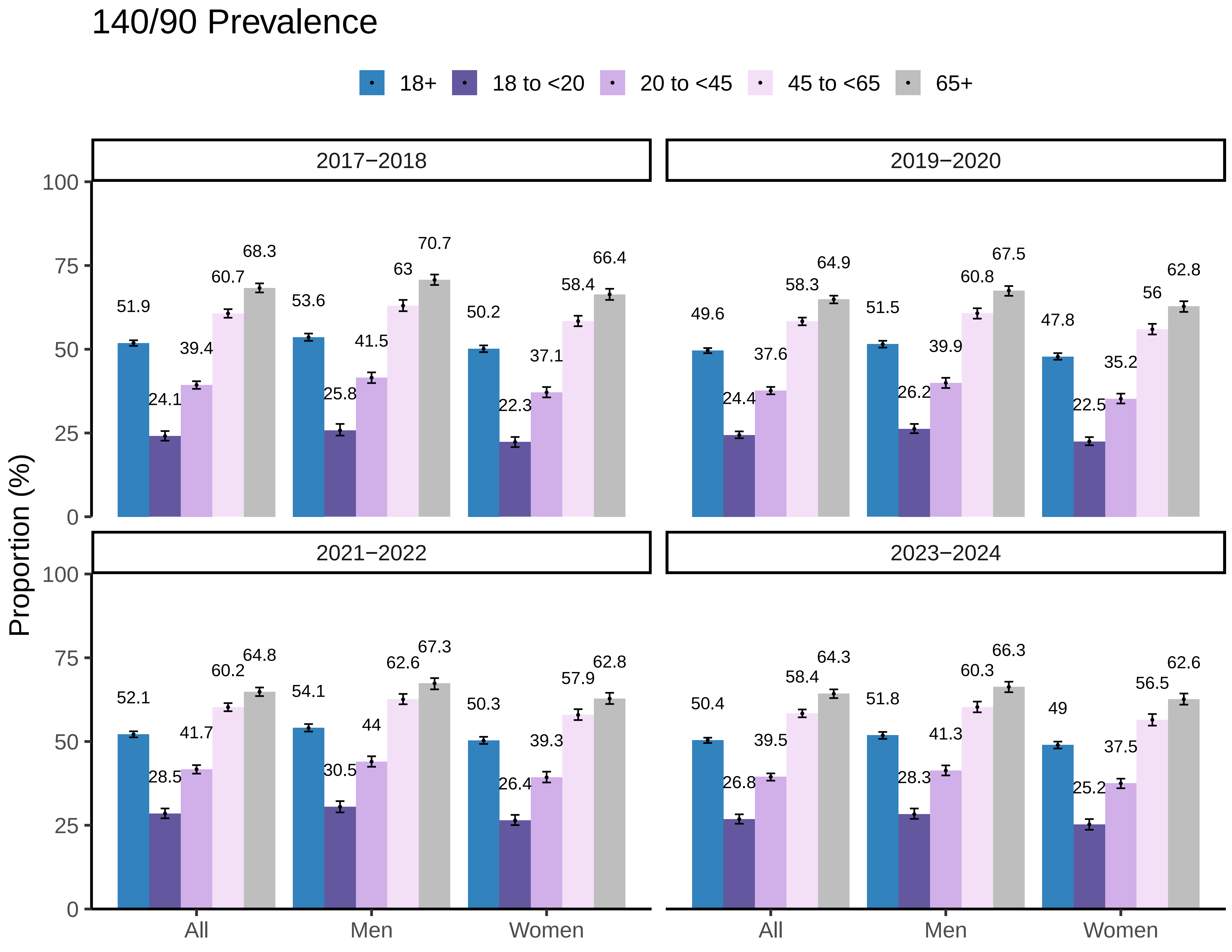

140/90 Prevalence
18+
18 to <20
20 to <45
45 to <65
65+
2017−2018
2019−2020
2021−2022
2023−2024
Proportion (%)
0
25
50
75
100
All
Men
Women
51.9
24.1
39.4
60.7
68.3
53.6
25.8
41.5
63
70.7
50.2
22.3
37.1
58.4
66.4
49.6
24.4
37.6
58.3
64.9
51.5
26.2
39.9
60.8
67.5
47.8
22.5
35.2
56
62.8
52.1
28.5
41.7
60.2
64.8
54.1
30.5
44
62.6
67.3
50.3
26.4
39.3
57.9
62.8
50.4
26.8
39.5
58.4
64.3
51.8
28.3
41.3
60.3
66.3
49
25.2
37.5
56.5
62.6

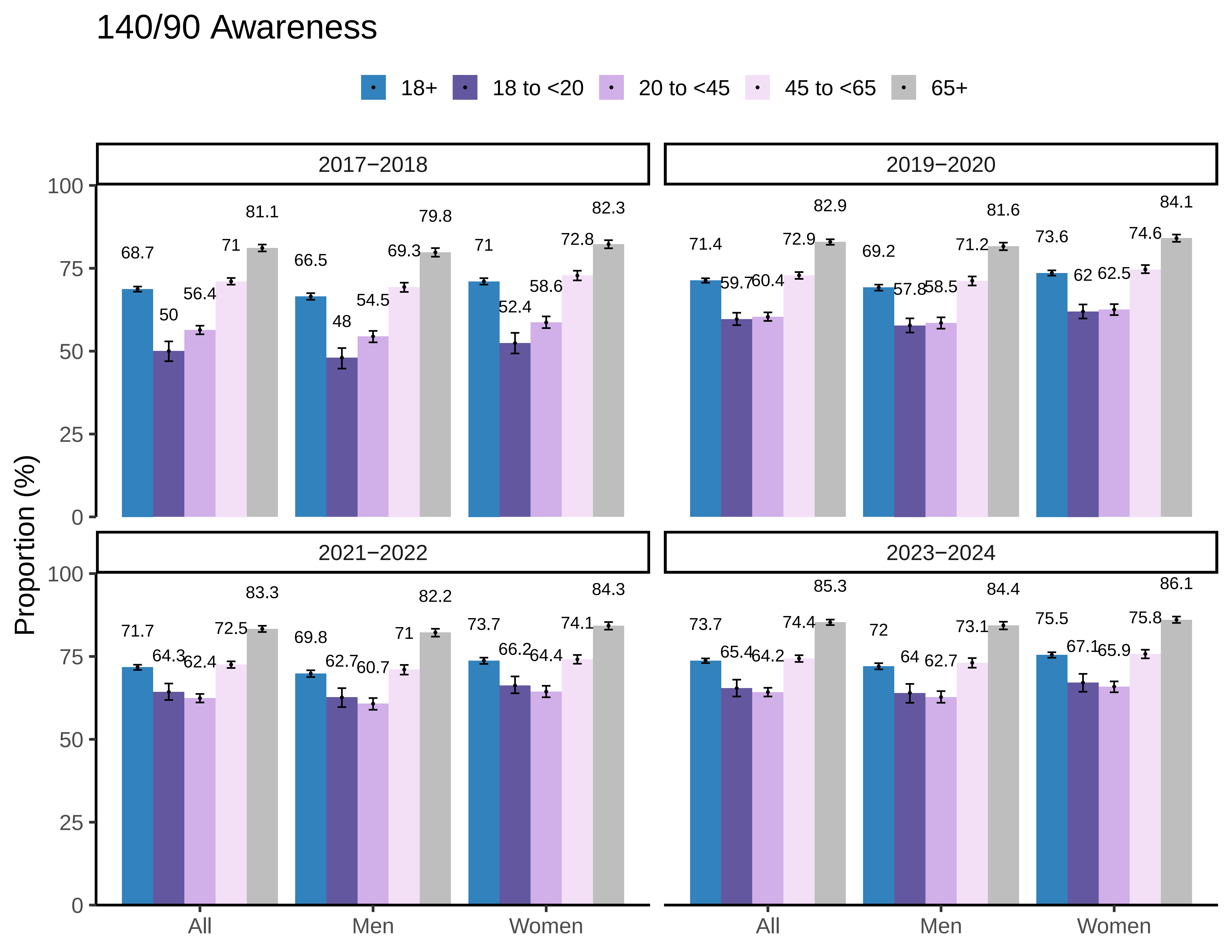

140/90 Awareness
18+
18 to <20
20 to <45
45 to <65
65+
2017−2018
2019−2020
2021−2022
2023−2024
Proportion (%)
0
25
50
75
100
All
Men
Women
68.7
50
56.4
71
81.1
66.5
48
54.5
69.3
79.8
71
52.4
58.6
72.8
82.3
71.4
59.7
60.4
72.9
82.9
69.2
57.8
58.5
71.2
81.6
73.6
62
62.5
74.6
84.1
71.7
64.3
62.4
72.5
83.3
69.8
62.7
60.7
71
82.2
73.7
66.2
64.4
74.1
84.3
73.7
65.4
64.2
74.4
85.3
72
64
62.7
73.1
84.4
75.5
67.1
65.9
75.8
86.1

### 140/90 Controlled

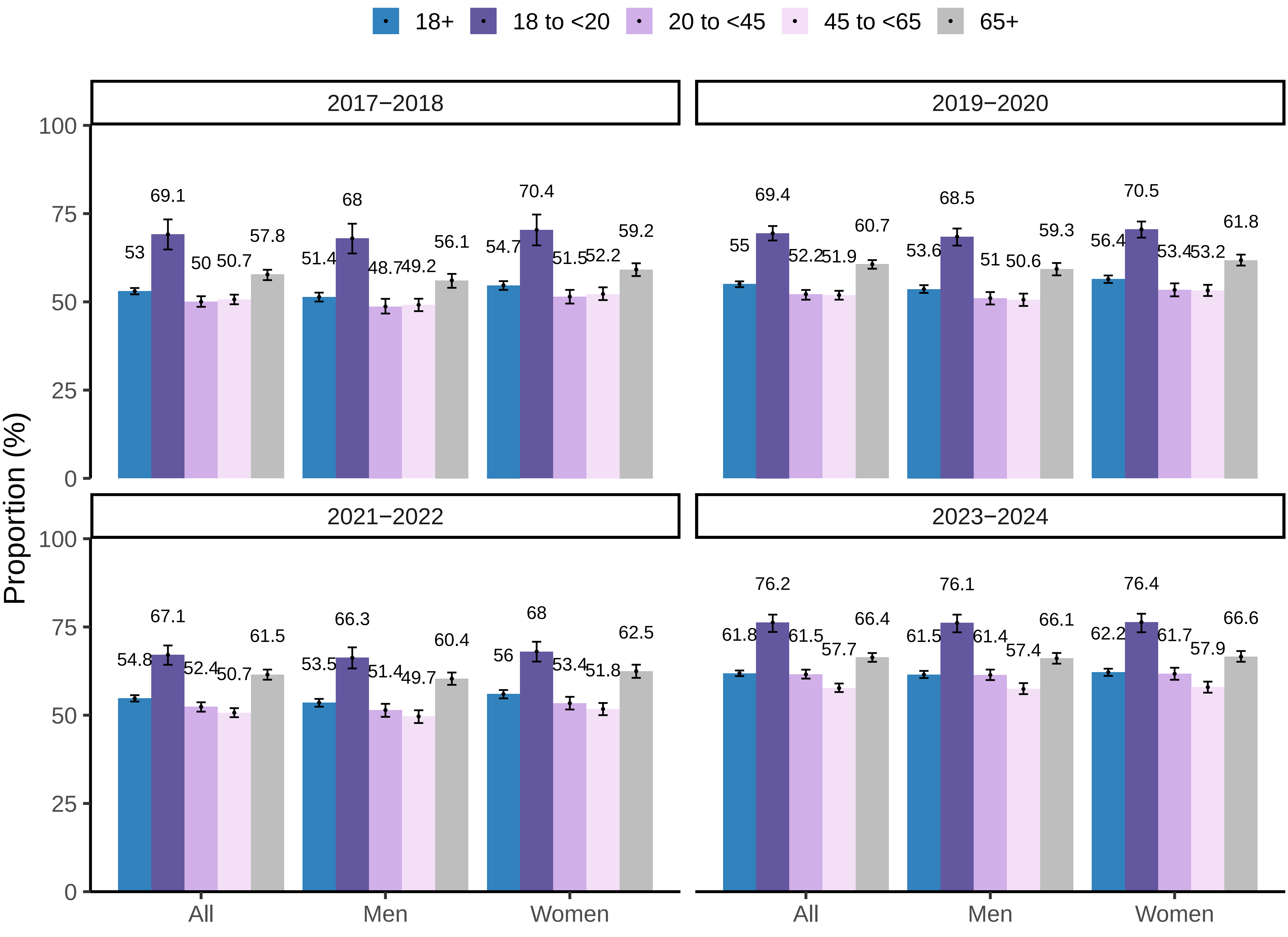

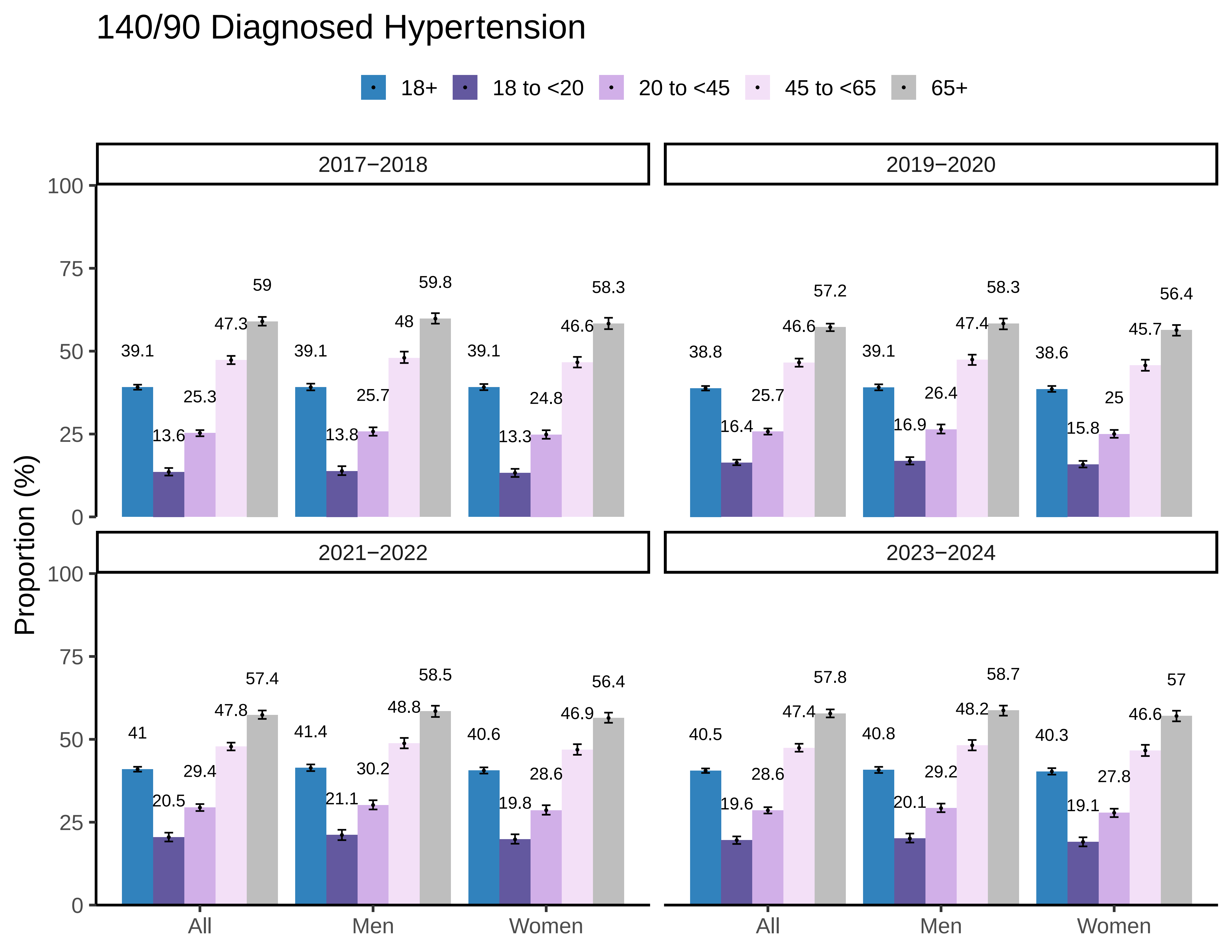
140/90 Diagnosed Hypertension
18+
18 to <20
20 to <45
45 to <65
65+
2017−2018
2019−2020
2021−2022
2023−2024
Proportion (%)
0
25
50
75
100
All
Men
Women
39.1
13.6
25.3
47.3
59
39.1
13.8
25.7
48
59.8
39.1
13.3
24.8
46.6
58.3
38.8
16.4
25.7
46.6
57.2
39.1
16.9
26.4
47.4
58.3
38.6
15.8
25
45.7
56.4
41
20.5
29.4
47.8
57.4
41.4
21.1
30.2
48.8
58.5
40.6
19.8
28.6
46.9
56.4
40.5
19.6
28.6
47.4
57.8
40.8
20.1
29.2
48.2
58.7
40.3
19.1
27.8
46.6
57

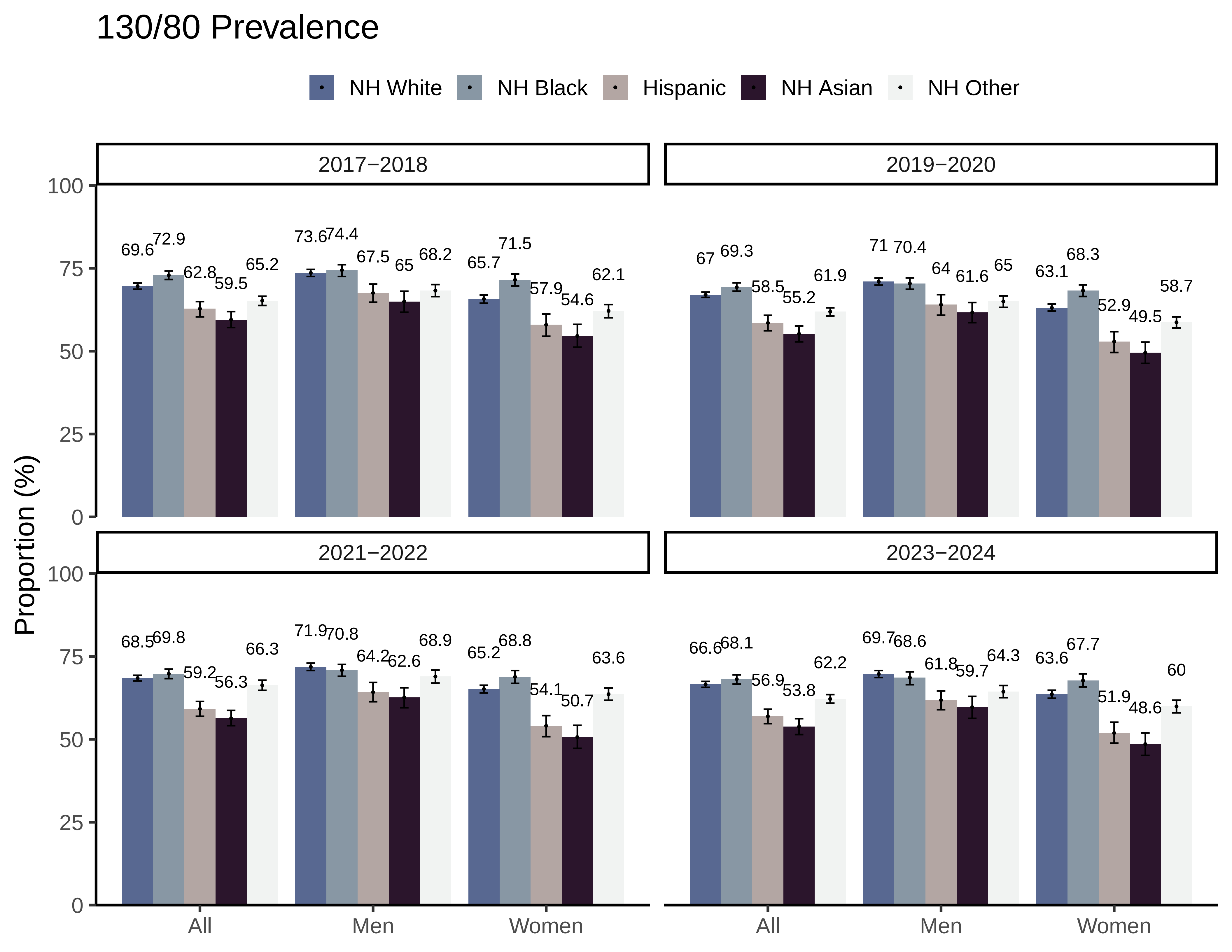

130/80 Prevalence
NH White
NH Black
Hispanic
NH Asian
NH Other
2017−2018
2019−2020
2021−2022
2023−2024
Proportion (%)
0
25
50
75
100
All
Men
Women
69.6
72.9
62.8
59.5
65.2
73.6
74.4
67.5
65
68.2
65.7
71.5
57.9
54.6
62.1
67
69.3
58.5
55.2
61.9
71
70.4
64
61.6
65
63.1
68.3
52.9
49.5
58.7
68.5
69.8
59.2
56.3
66.3
71.9
70.8
64.2
62.6
68.9
65.2
68.8
54.1
50.7
63.6
66.6
68.1
56.9
53.8
62.2
69.7
68.6
61.8
59.7
64.3
63.6
67.7
51.9
48.6
60

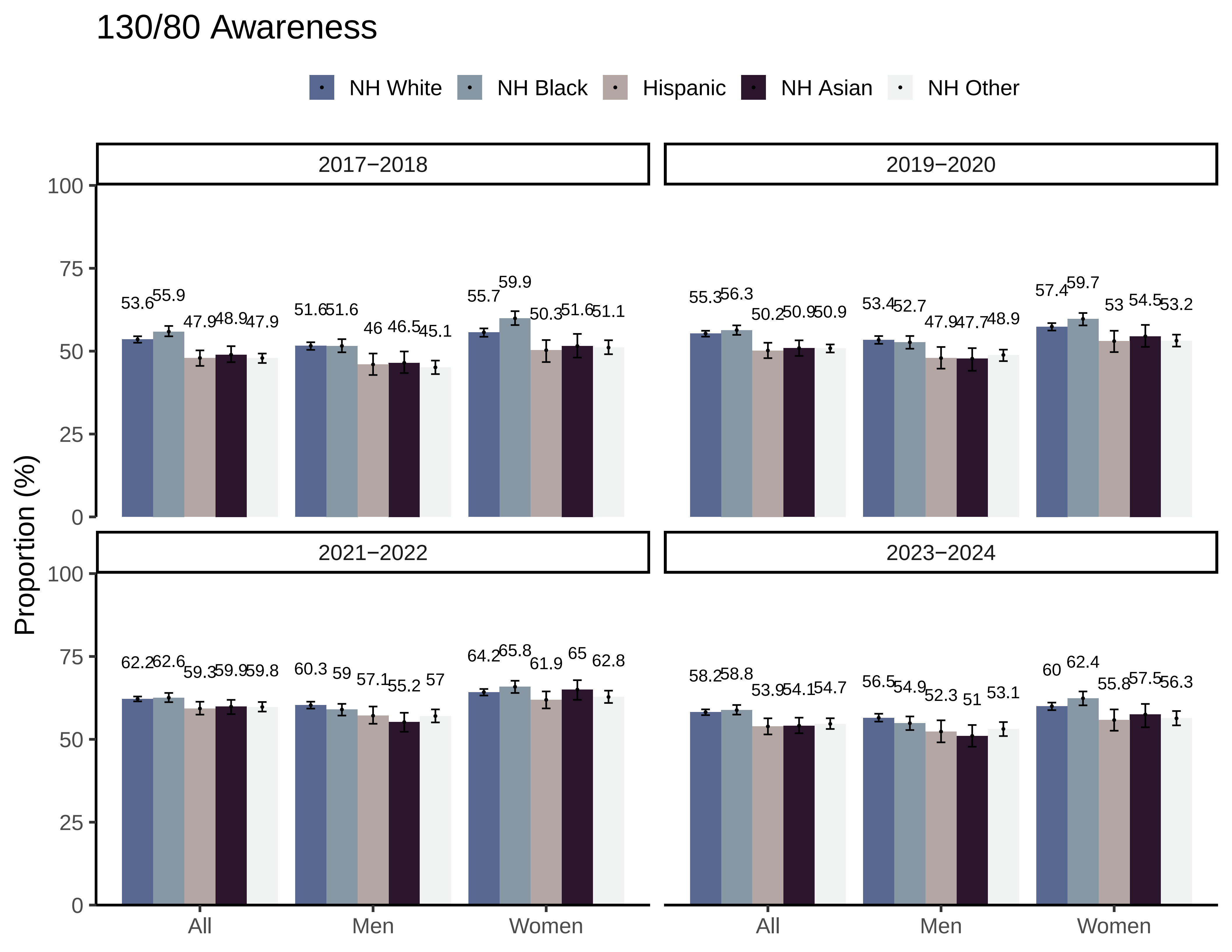
130/80 Awareness
NH White
NH Black
Hispanic
NH Asian
NH Other
2017−2018
2019−2020
2021−2022
2023−2024
Proportion (%)
0
25
50
75
100
All
Men
Women
53.6
55.9
47.9
48.9
47.9
51.6
51.6
46
46.5
45.1
55.7
59.9
50.3
51.6
51.1
55.3
56.3
50.2
50.9
50.9
53.4
52.7
47.9
47.7
48.9
57.4
59.7
53
54.5
53.2
62.2
62.6
59.3
59.9
59.8
60.3
59
57.1
55.2
57
64.2
65.8
61.9
65
62.8
58.2
58.8
53.9
54.1
54.7
56.5
54.9
52.3
51
53.1
60
62.4
55.8
57.5
56.3

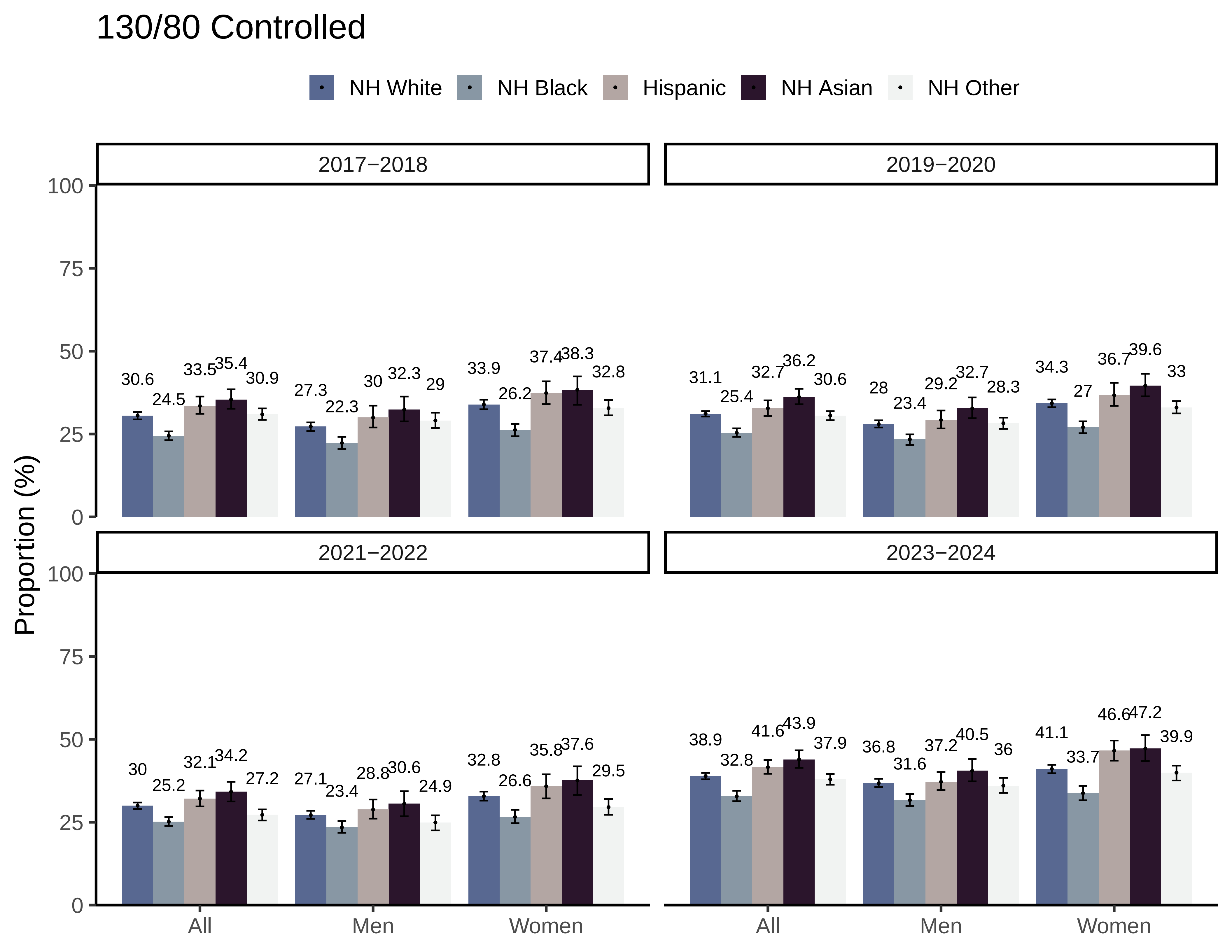

130/80 Controlled
NH White
NH Black
Hispanic
NH Asian
NH Other
2017−2018
2019−2020
2021−2022
2023−2024
Proportion (%)
100
75
50
25
0
All
Men
Women
30.6
24.5
33.5
35.4
30.9
27.3
22.3
30
32.3
29
33.9
26.2
37.4
38.3
32.8
31.1
25.4
32.7
36.2
30.6
28
23.4
29.2
32.7
28.3
34.3
27
36.7
39.6
33
30
25.2
32.1
34.2
27.2
27.1
23.4
28.8
30.6
24.9
32.8
26.6
35.8
37.6
29.5
38.9
32.8
41.6
43.9
37.9
36.8
31.6
37.2
40.5
36
41.1
33.7
46.6
47.2
39.9

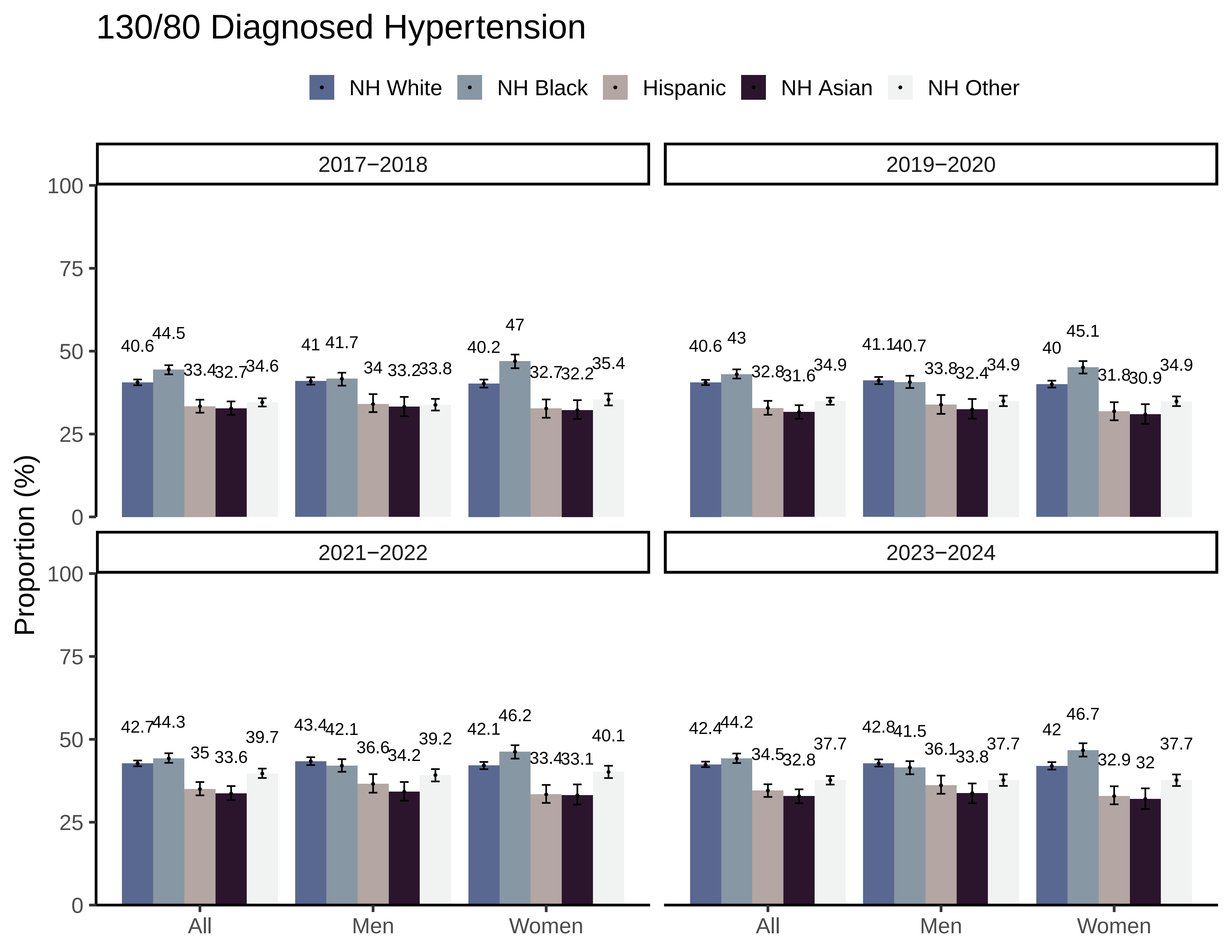
130/80 Diagnosed Hypertension
NH White
NH Black
Hispanic
NH Asian
NH Other
2017−2018
2019−2020
2021−2022
2023−2024
Proportion (%)
0
25
50
75
100
All
Men
Women
40.6
44.5
33.4
32.7
34.6
41
41.7
34
33.2
33.8
40.2
47
32.7
32.2
35.4
40.6
43
32.8
31.6
34.9
41.1
40.7
33.8
32.4
34.9
40
45.1
31.8
30.9
34.9
42.7
44.3
35
33.6
39.7
43.4
42.1
36.6
34.2
39.2
42.1
46.2
33.4
33.1
40.1
42.4
44.2
34.5
32.8
37.7
42.8
41.5
36.1
33.8
37.7
42
46.7
32.9
32
37.7

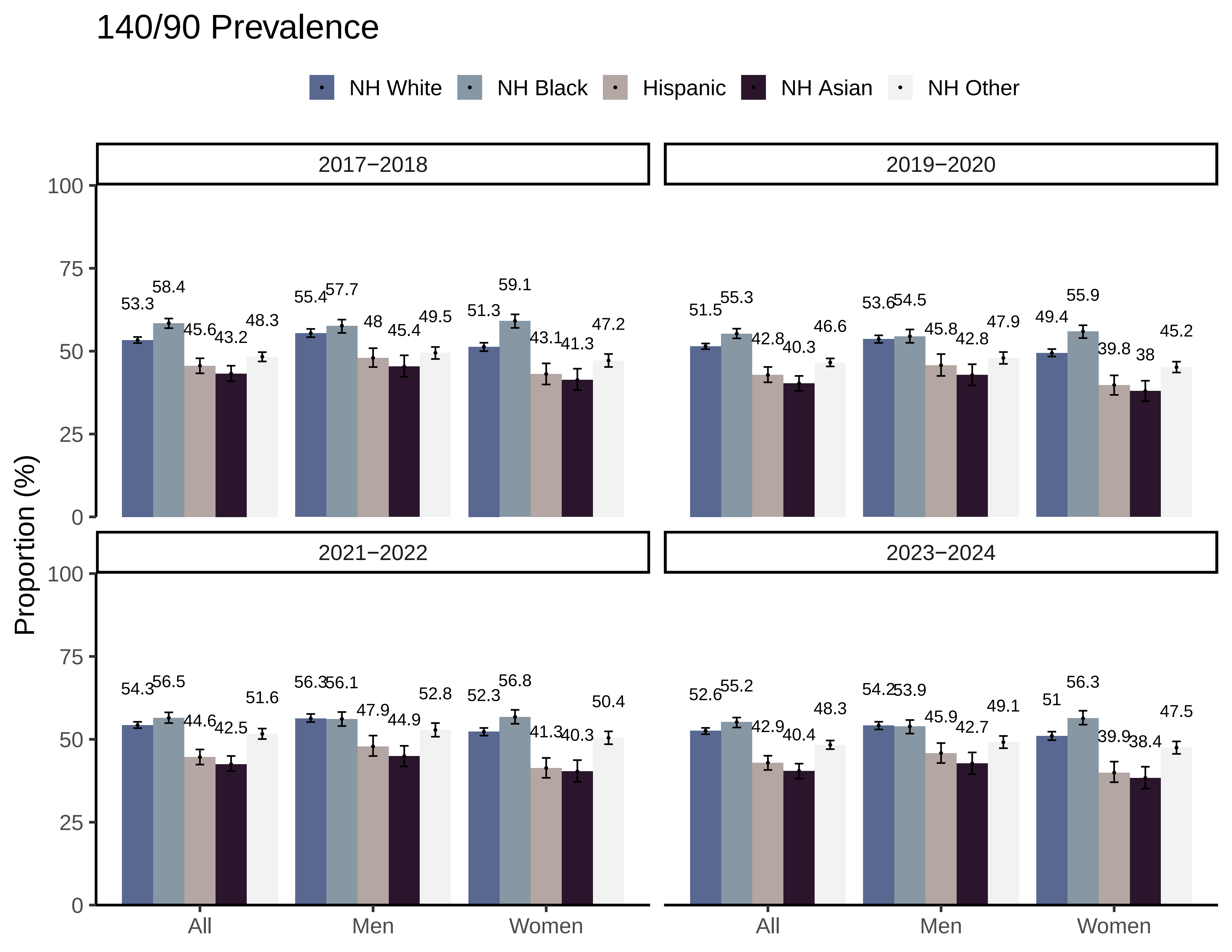
140/90 Prevalence
NH White
NH Black
Hispanic
NH Asian
NH Other
2017−2018
2019−2020
2021−2022
2023−2024
Proportion (%)
100
75
50
25
0
53.3
58.4
45.6
43.2
48.3
55.4
57.7
48
45.4
49.5
51.3
59.1
43.1
41.3
47.2
51.5
55.3
42.8
40.3
46.6
53.6
54.5
45.8
42.8
47.9
49.4
55.9
39.8
38
45.2
54.3
56.5
44.6
42.5
51.6
56.3
56.1
47.9
44.9
52.8
52.3
56.8
41.3
40.3
50.4
52.6
55.2
42.9
40.4
48.3
54.2
53.9
45.9
42.7
49.1
51
56.3
39.9
38.4
47.5
All
Men
Women

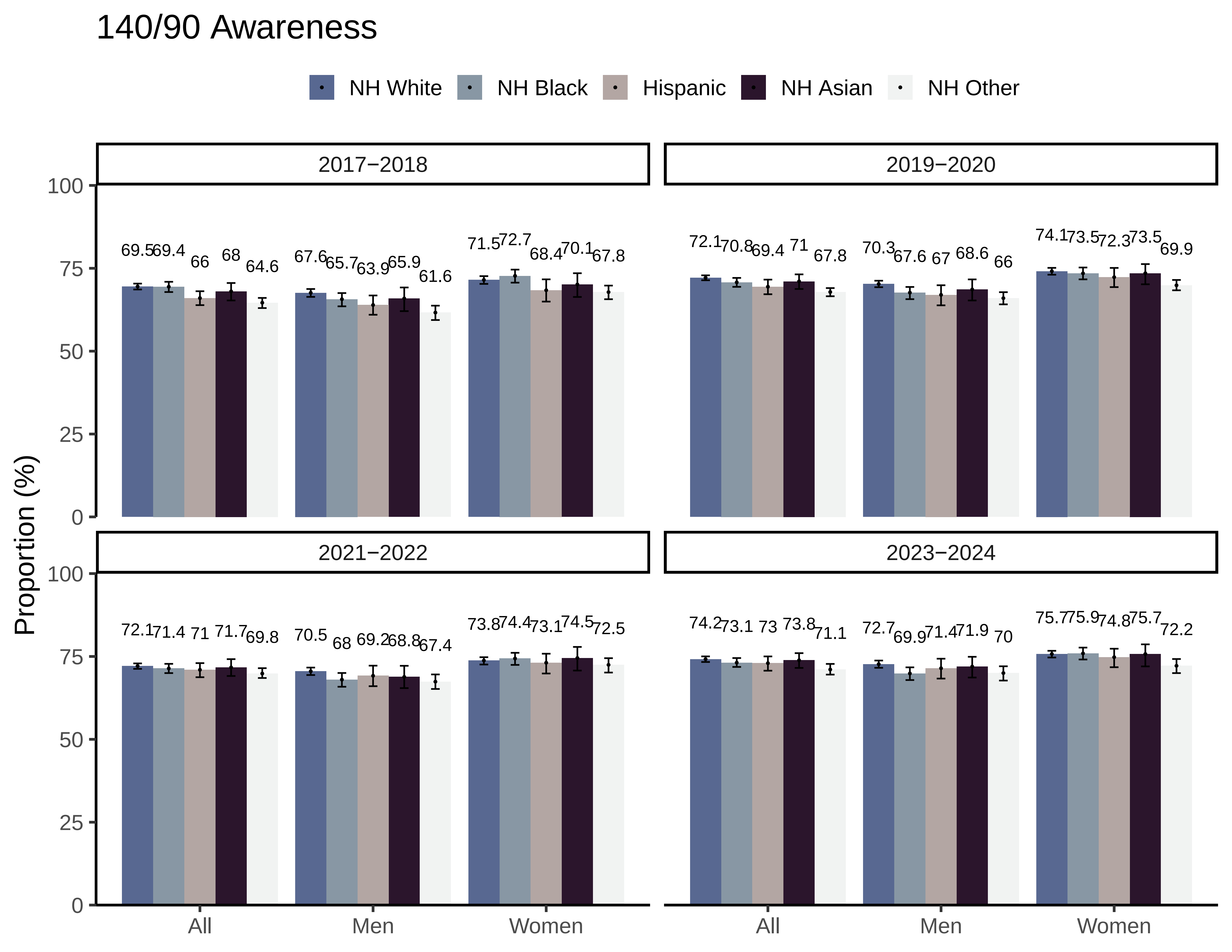

140/90 Awareness
NH White
NH Black
Hispanic
NH Asian
NH Other
2017−2018
2019−2020
2021−2022
2023−2024
Proportion (%)
0
25
50
75
100
69.5 69.4 66 68 64.6
67.6 65.7 63.9 65.9 61.6
71.5 72.7 68.4 70.1 67.8
72.1 70.8 69.4 71 67.8
70.3 67.6 67 68.6 66
74.1 73.5 72.3 73.5 69.9
72.1 71.4 71 71.7 69.8
70.5 68 69.2 68.8 67.4
73.8 74.4 73.1 74.5 72.5
74.2 73.1 73 73.8 71.1
72.7 69.9 71.4 71.9 70
75.7 75.9 74.8 75.7 72.2
All
Men
Women

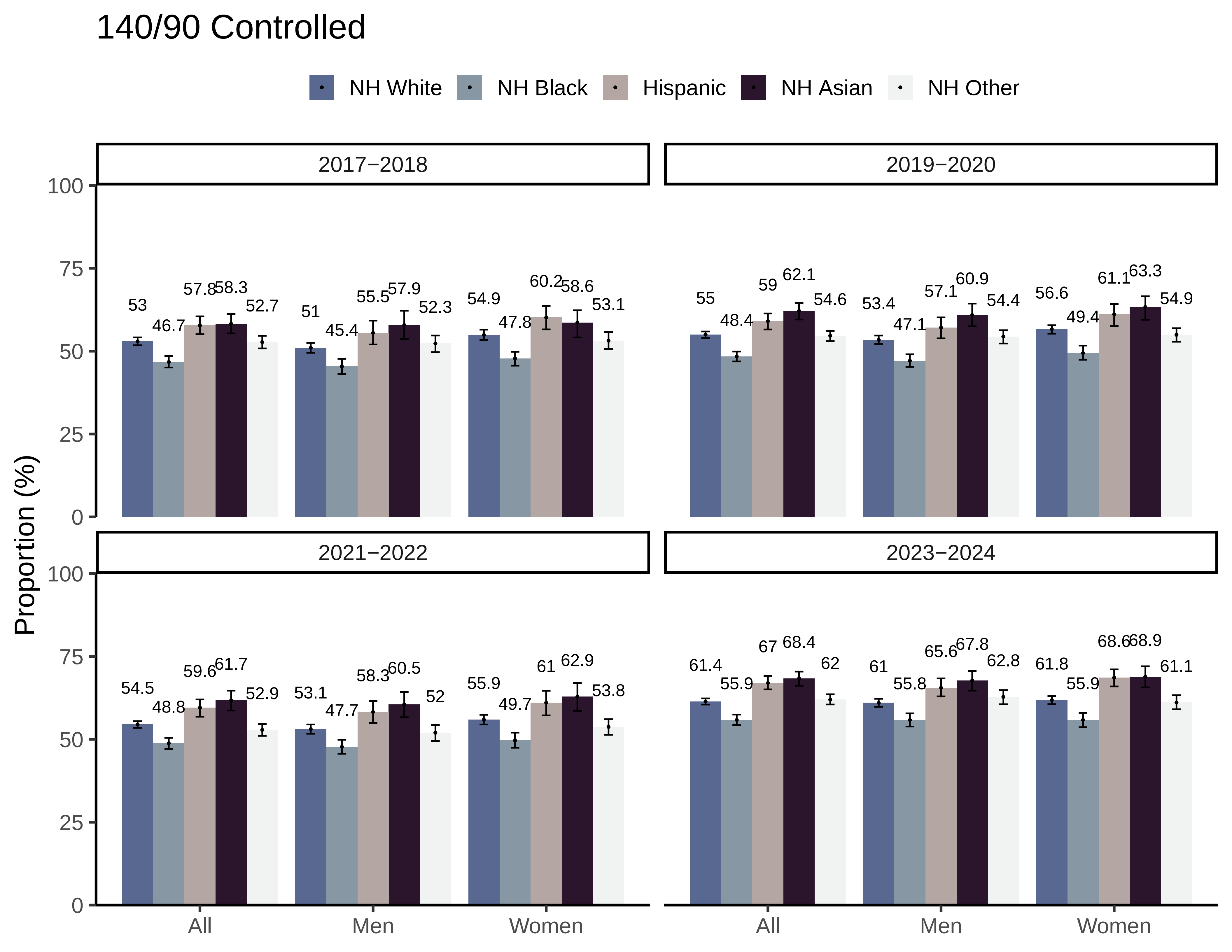

140/90 Controlled
NH White
NH Black
Hispanic
NH Asian
NH Other
2017−2018
2019−2020
2021−2022
2023−2024
Proportion (%)
100
75
50
25
0
All
Men
Women
53
46.7
57.8
58.3
52.7
51
45.4
55.5
57.9
52.3
54.9
47.8
60.2
58.6
53.1
55
48.4
59
62.1
54.6
53.4
47.1
57.1
60.9
54.4
56.6
49.4
61.1
63.3
54.9
54.5
48.8
59.6
61.7
52.9
53.1
47.7
58.3
60.5
52
55.9
49.7
61
62.9
53.8
61.4
55.9
67
68.4
62
61
55.8
65.6
67.8
62.8
61.8
55.9
68.6
68.9
61.1

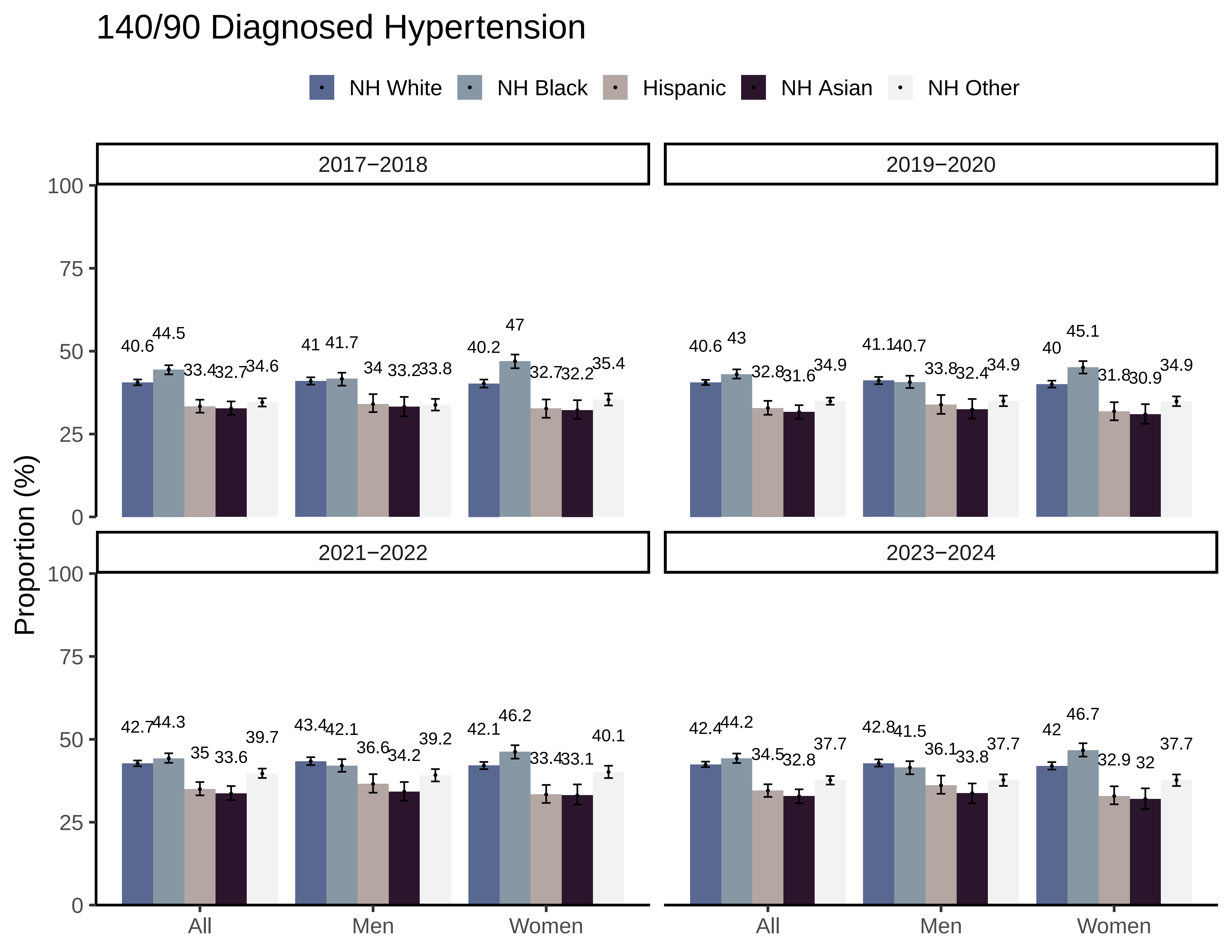

140/90 Diagnosed Hypertension
NH White
NH Black
Hispanic
NH Asian
NH Other
2017−2018
2019−2020
2021−2022
2023−2024
Proportion (%)
0
25
50
75
100
All
Men
Women
40.6 44.5 33.4 32.7 34.6
41 41.7 34 33.2 33.8
40.2 47 32.7 32.2 35.4
40.6 43 32.8 31.6 34.9
41.1 40.7 33.8 32.4 34.9
40 45.1 31.8 30.9 34.9
42.7 44.3 35 33.6 39.7
43.4 42.1 36.6 34.2 39.2
42.1 46.2 33.4 33.1 40.1
42.4 44.2 34.5 32.8 37.7
42.8 41.5 36.1 33.8 37.7
42 46.7 32.9 32 37.7
