## Supplementary File 4 for "County Prevalence and Control of High Blood Pressure from Health Kiosks, 2017-2024"

### 130/80 Prevalence

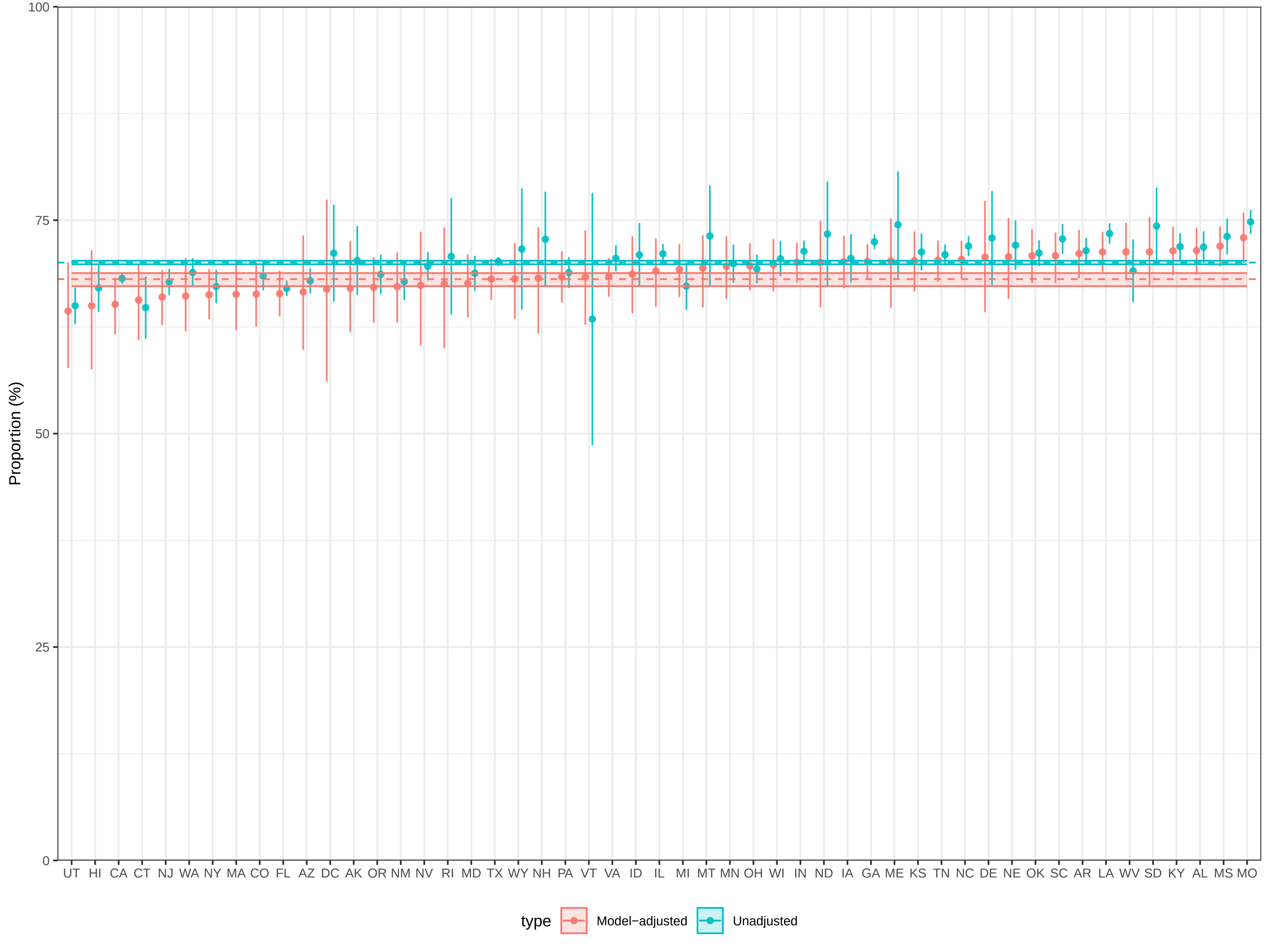

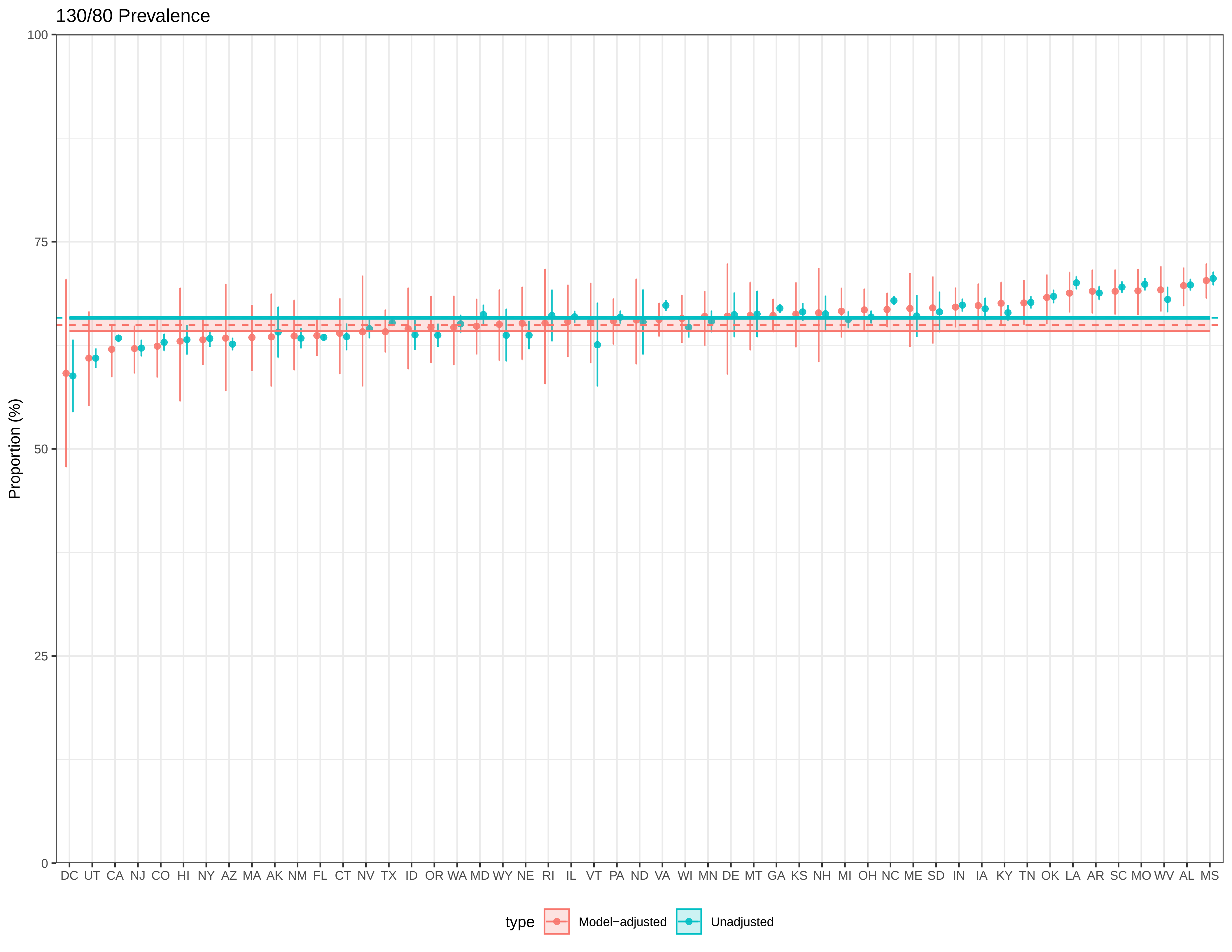

130/80 Prevalence
Proportion (%)
100
75
50
25
0
DC UT CA NJ CO HI NY AZ MA AK NM FL CT NV TX ID OR WA MD WY NE RI IL VT PA ND VA WI MN DE MT GA KS NH MI OH NC ME SD IN IA KY TN OK LA AR SC MO WV AL MS
type
Model−adjusted
Unadjusted

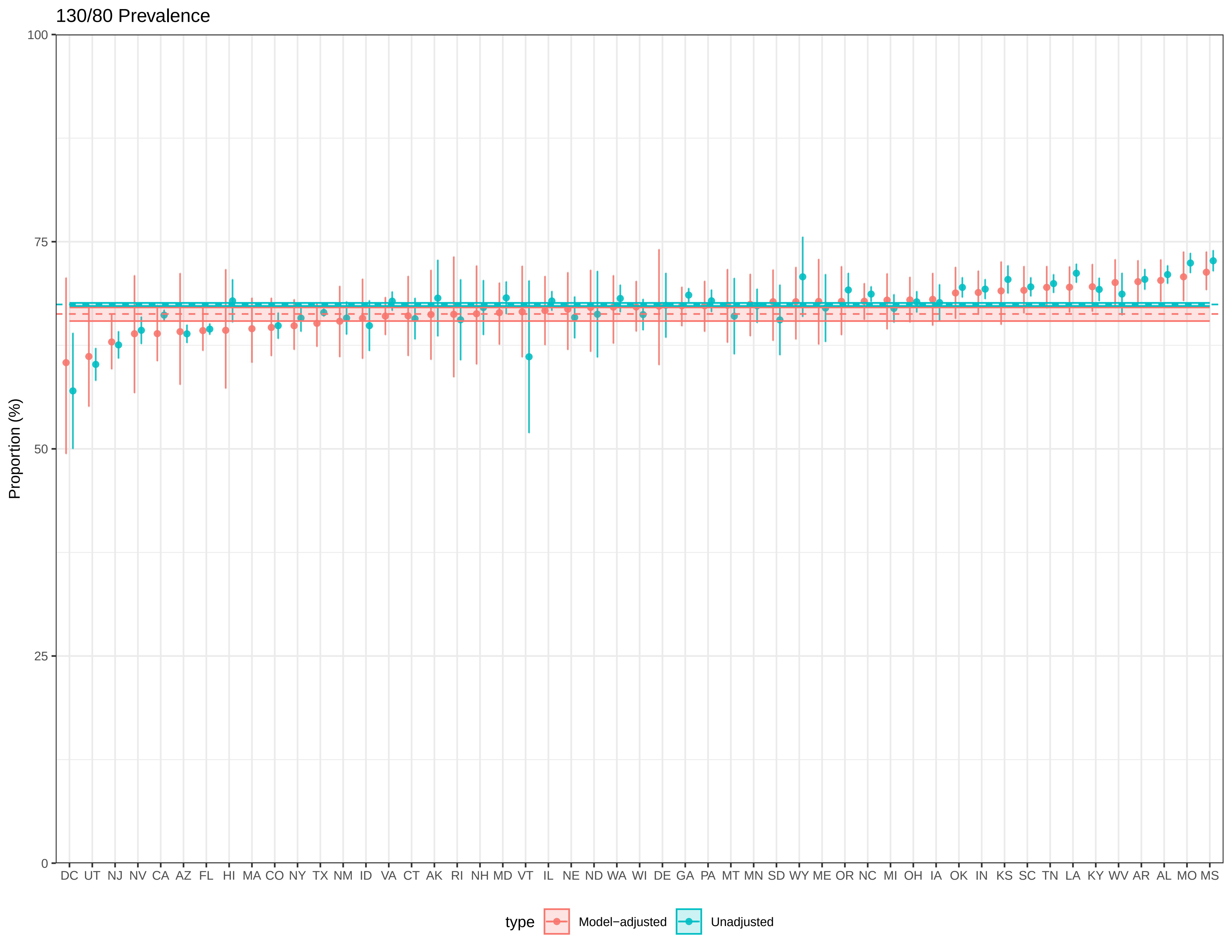

130/80 Prevalence
Proportion (%)
100
75
50
25
0
DC UT NJ NV CA AZ FL HI MA CO NY TX NM ID VA CT AK RI NH MD VT IL NE ND WA WI DE GA PA MT MN SD WY ME OR NC MI OH IA OK IN KS SC TN LA KY WV AR AL MO MS
type
Model−adjusted
Unadjusted

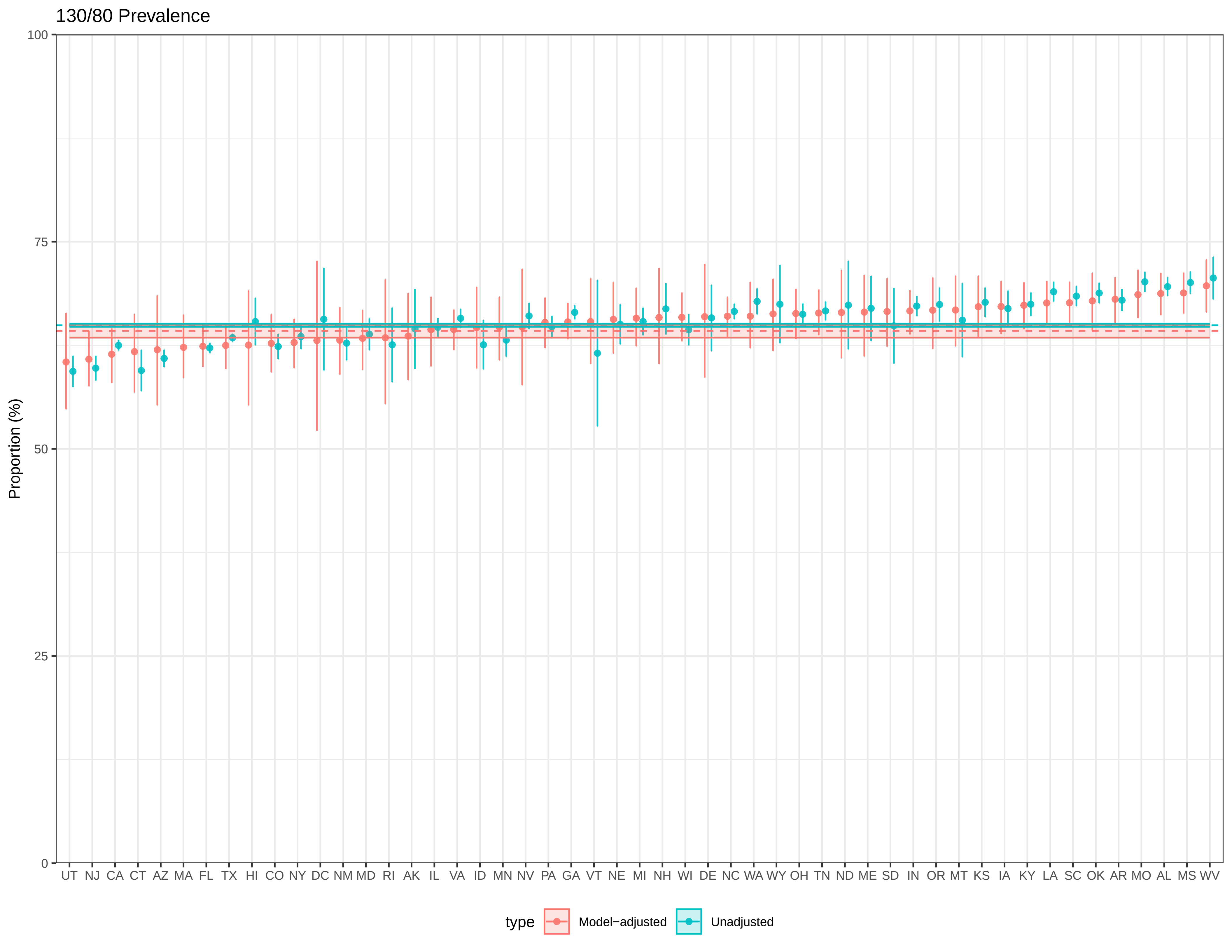
130/80 Prevalence
Proportion (%)
100
75
50
25
0
UT NJ CA CT AZ MA FL TX HI CO NY DC NM MD RI AK IL VA ID MN NV PA GA VT NE MI NH WI DE NC WA WY OH TN ND ME SD IN OR MT KS IA KY LA SC OK AR MO AL MS WV
type
Model−adjusted
Unadjusted

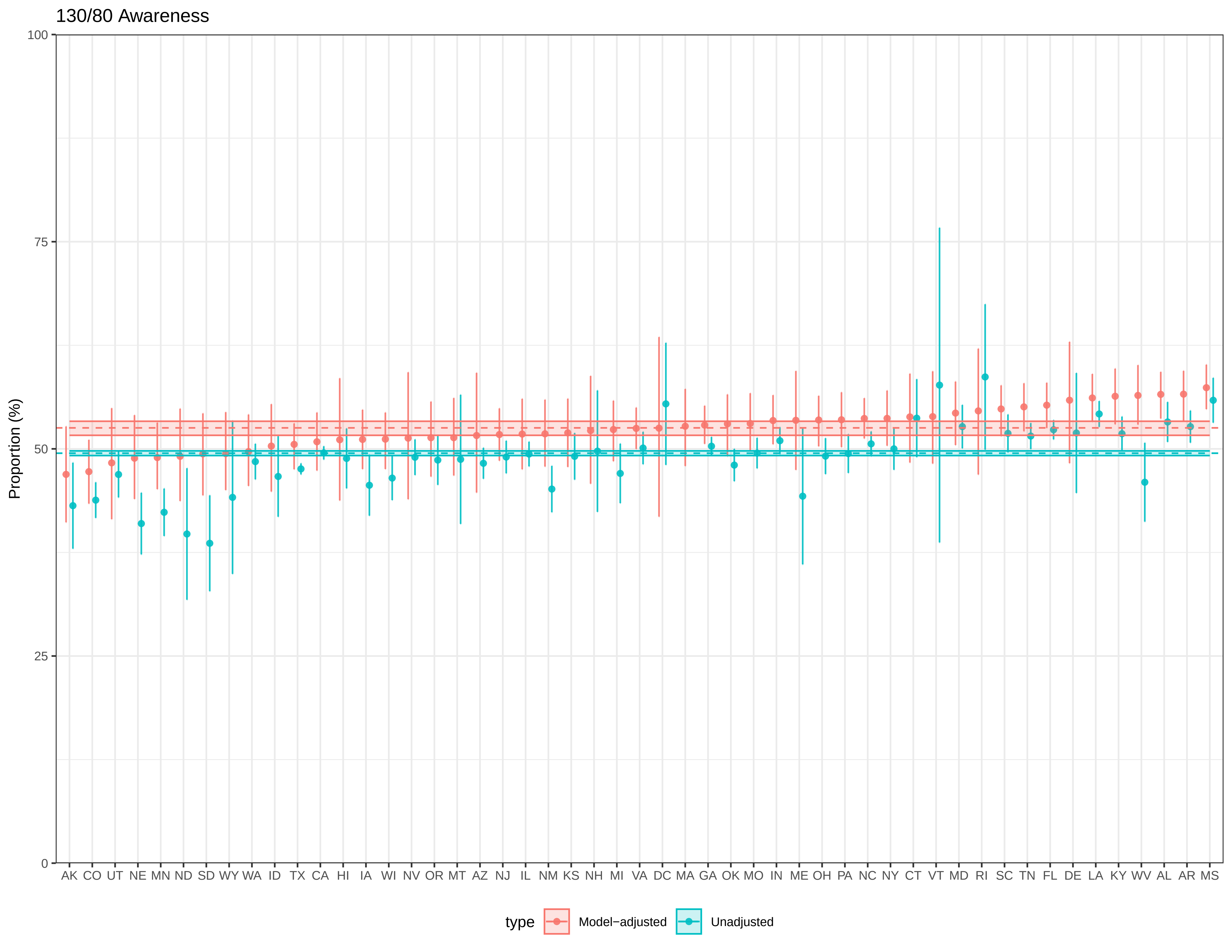

130/80 Awareness
Proportion (%)
100
75
50
25
0
AK CO UT NE MN ND SD WY WA ID TX CA HI IA WI NV OR MT AZ NJ IL NM KS NH MI VA DC MA GA OK MO IN ME OH PA NC NY CT VT MD RI SC TN FL DE LA KY WV AL AR MS
type
Model−adjusted
Unadjusted

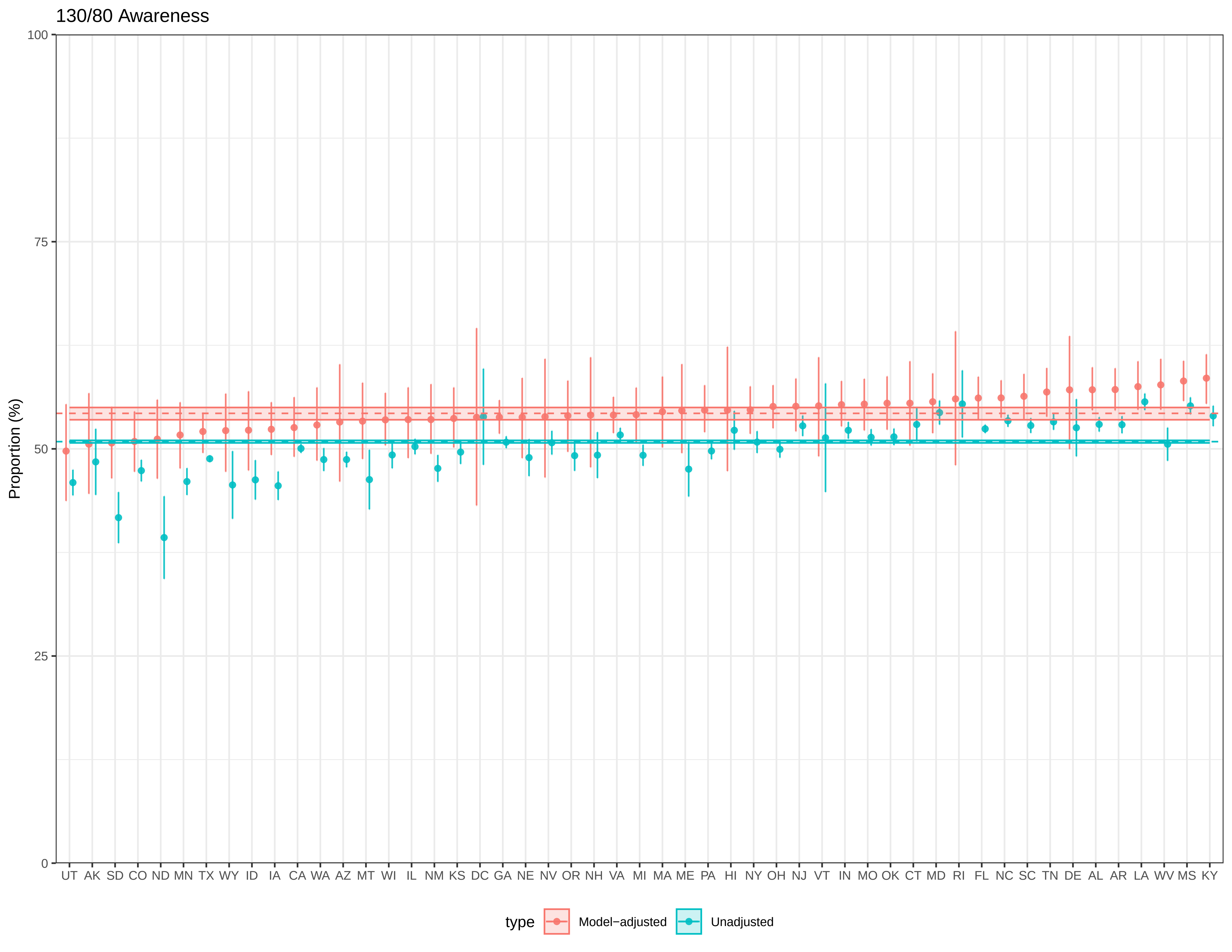
130/80 Awareness
Proportion (%)
100
75
50
25
0
UT AK SD CO ND MN TX WY ID IA CA WA AZ MT WI IL NM KS DC GA NE NV OR NH VA MI MA ME PA HI NY OH NJ VT IN MO OK CT MD RI FL NC SC TN DE AL AR LA WV MS KY
type
Model−adjusted
Unadjusted

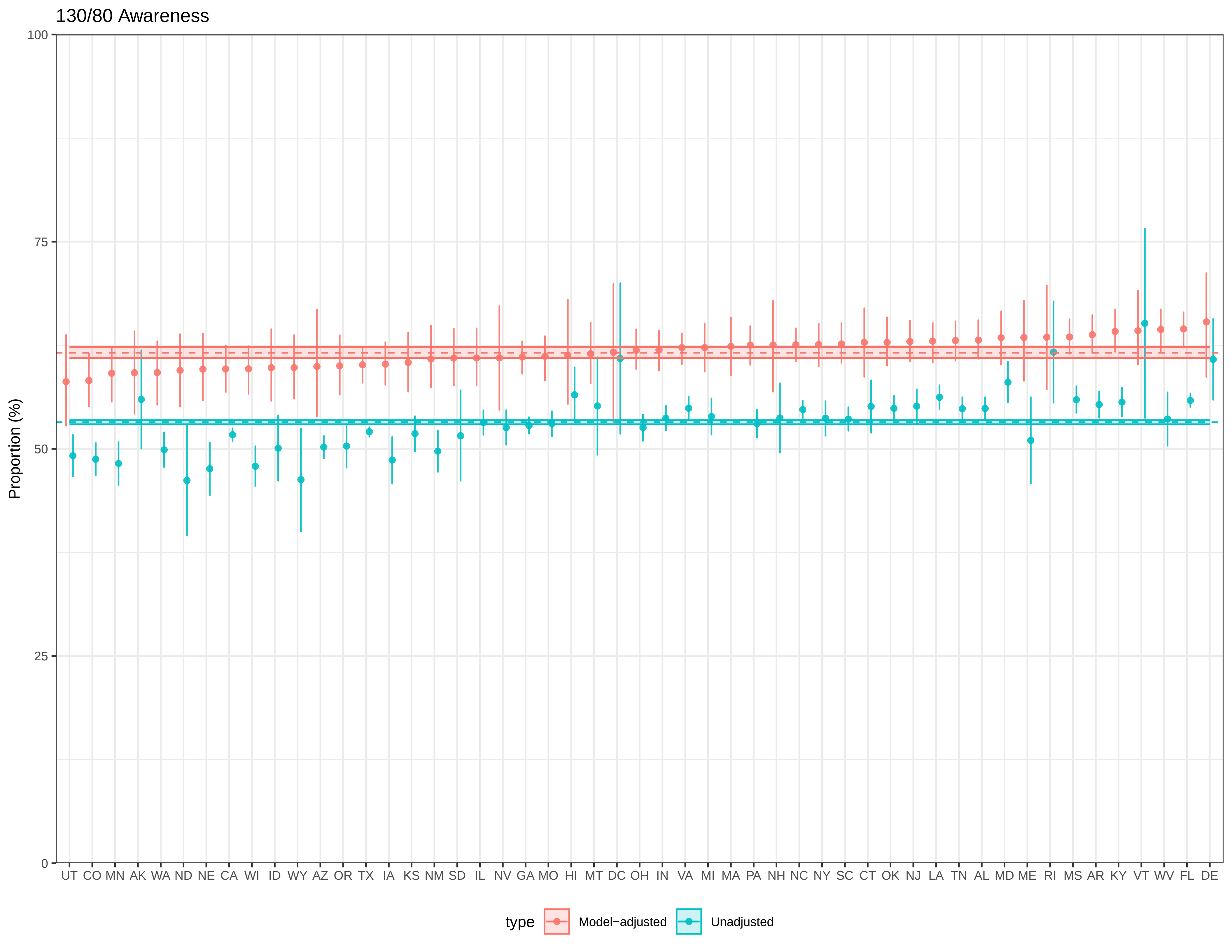

130/80 Awareness
Proportion (%)
100
75
50
25
0
UT CO MN AK WA ND NE CA WI ID WY AZ OR TX IA KS NM SD IL NV GA MO HI MT DC OH IN VA MI MA PA NH NC NY SC CT OK NJ LA TN AL MD ME RI MS AR KY VT WV FL DE
type
Model−adjusted
Unadjusted

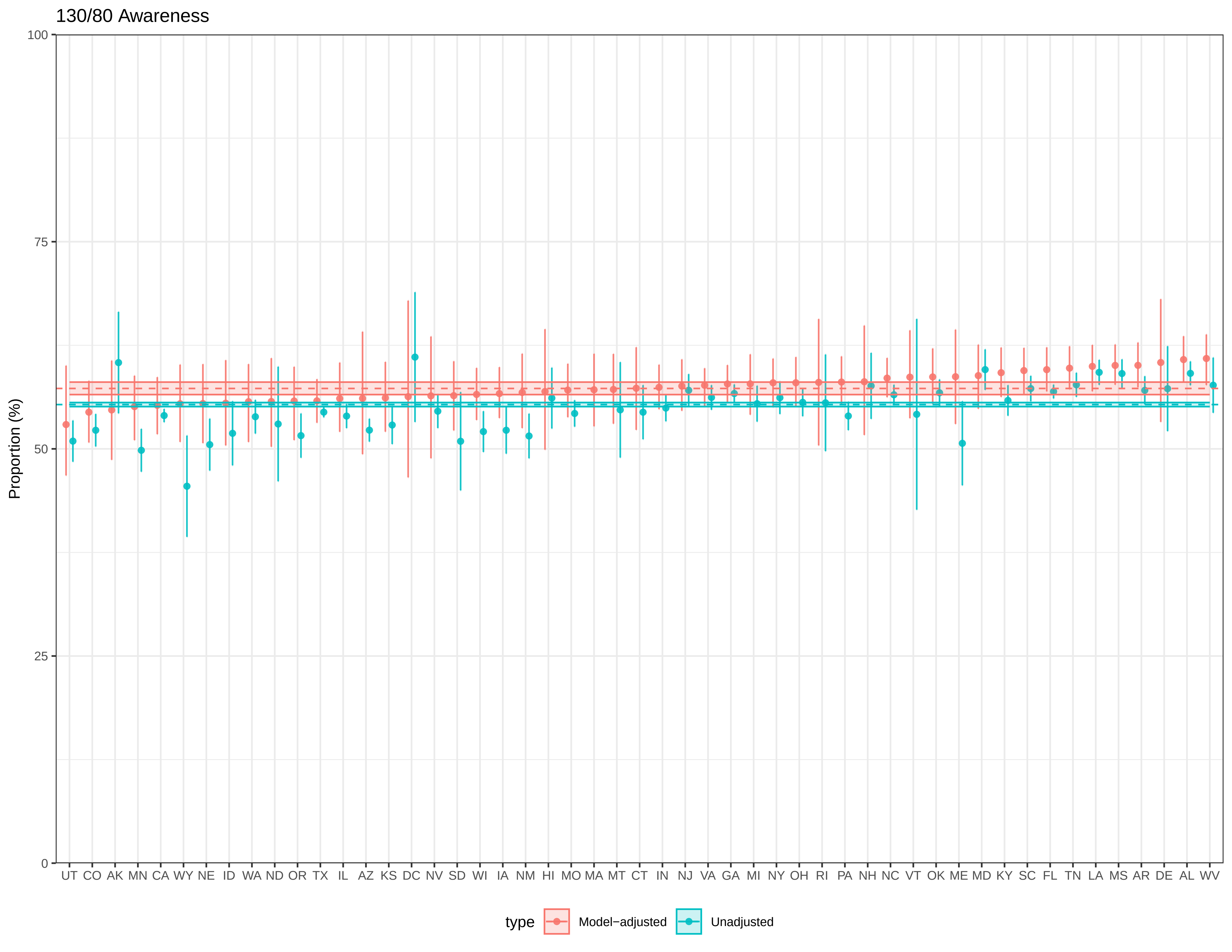

130/80 Awareness
Proportion (%)
100
75
50
25
0
UT CO AK MN CA WY NE ID WA ND OR TX IL AZ KS DC NV SD WI IA NM HI MO MA MT CT IN NJ VA GA MI NY OH RI PA NH NC VT OK ME MD KY SC FL TN LA MS AR DE AL WV
type
Model−adjusted
Unadjusted

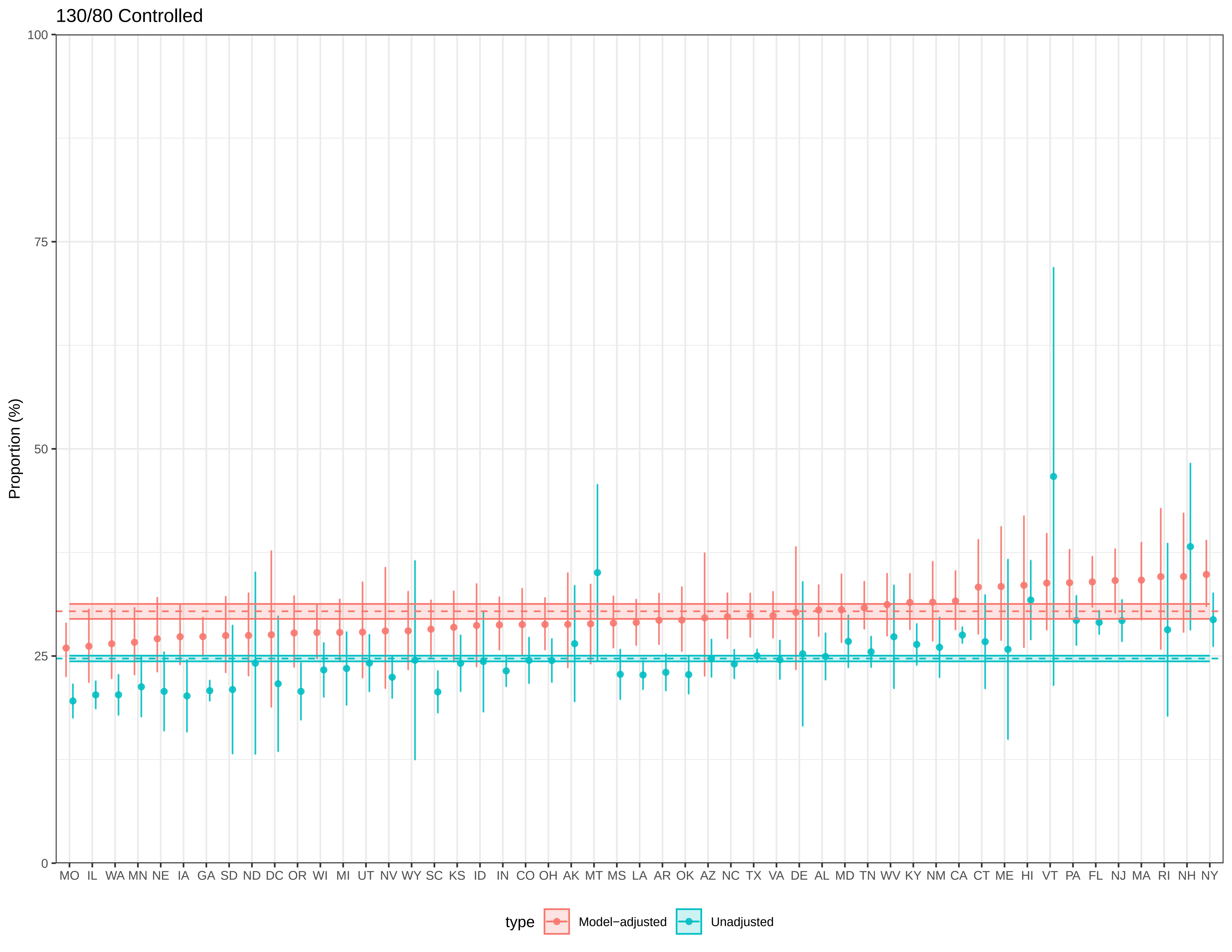

130/80 Controlled
Proportion (%)
100
75
50
25
0
MO IL WA MN NE IA GA SD ND DC OR WI MI UT NV WY SC KS ID IN CO OH AK MT MS LA AR OK AZ NC TX VA DE AL MD TN WV KY NM CA CT ME HI VT PA FL NJ MA RI NH NY
type
Model−adjusted
Unadjusted

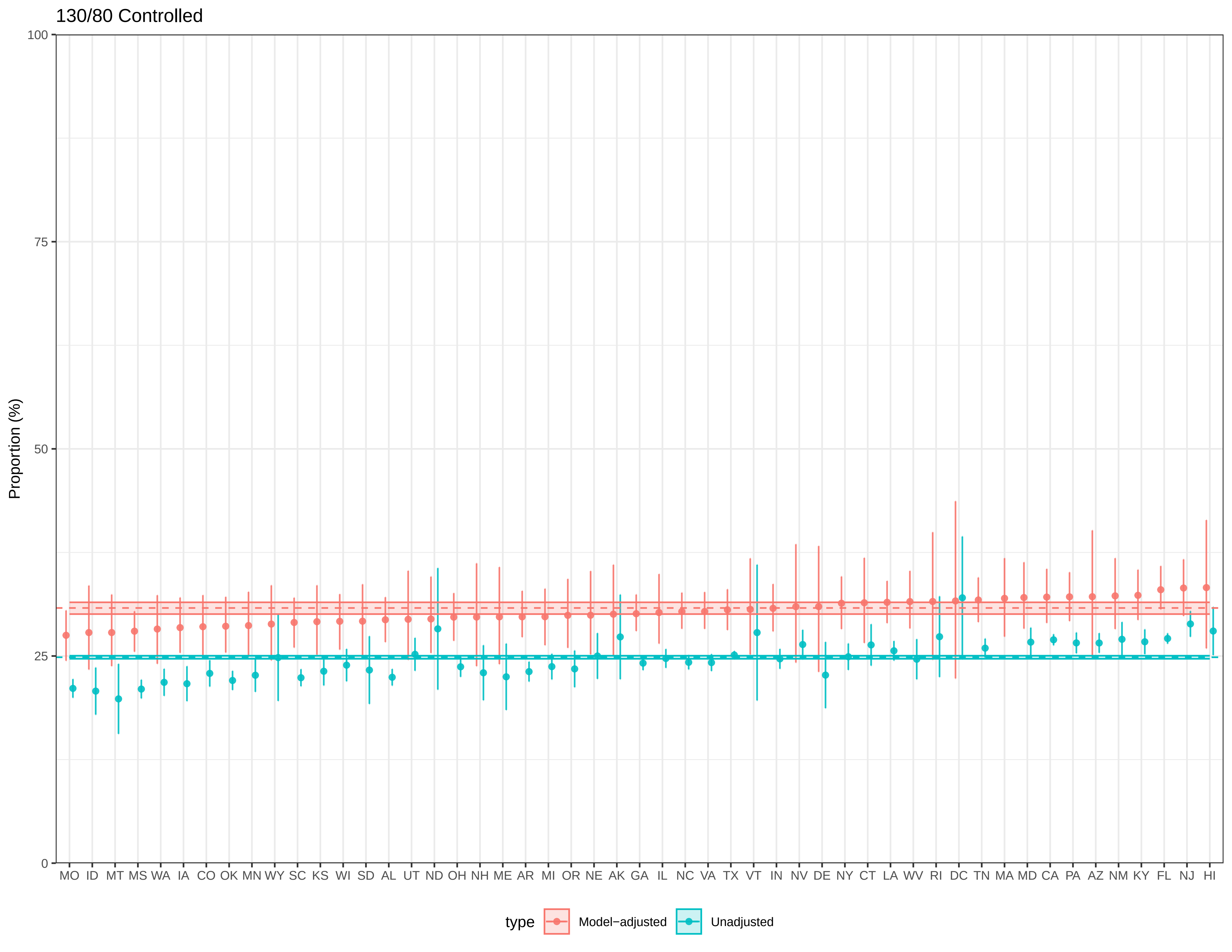
130/80 Controlled
Proportion (%)
100
75
50
25
0
MO ID MT MS WA IA CO OK MN WY SC KS WI SD AL UT ND OH NH ME AR MI OR NE AK GA IL NC VA TX VT IN NV DE NY CT LA WV RI DC TN MA MD CA PA AZ NM KY FL NJ HI
type
Model−adjusted
Unadjusted

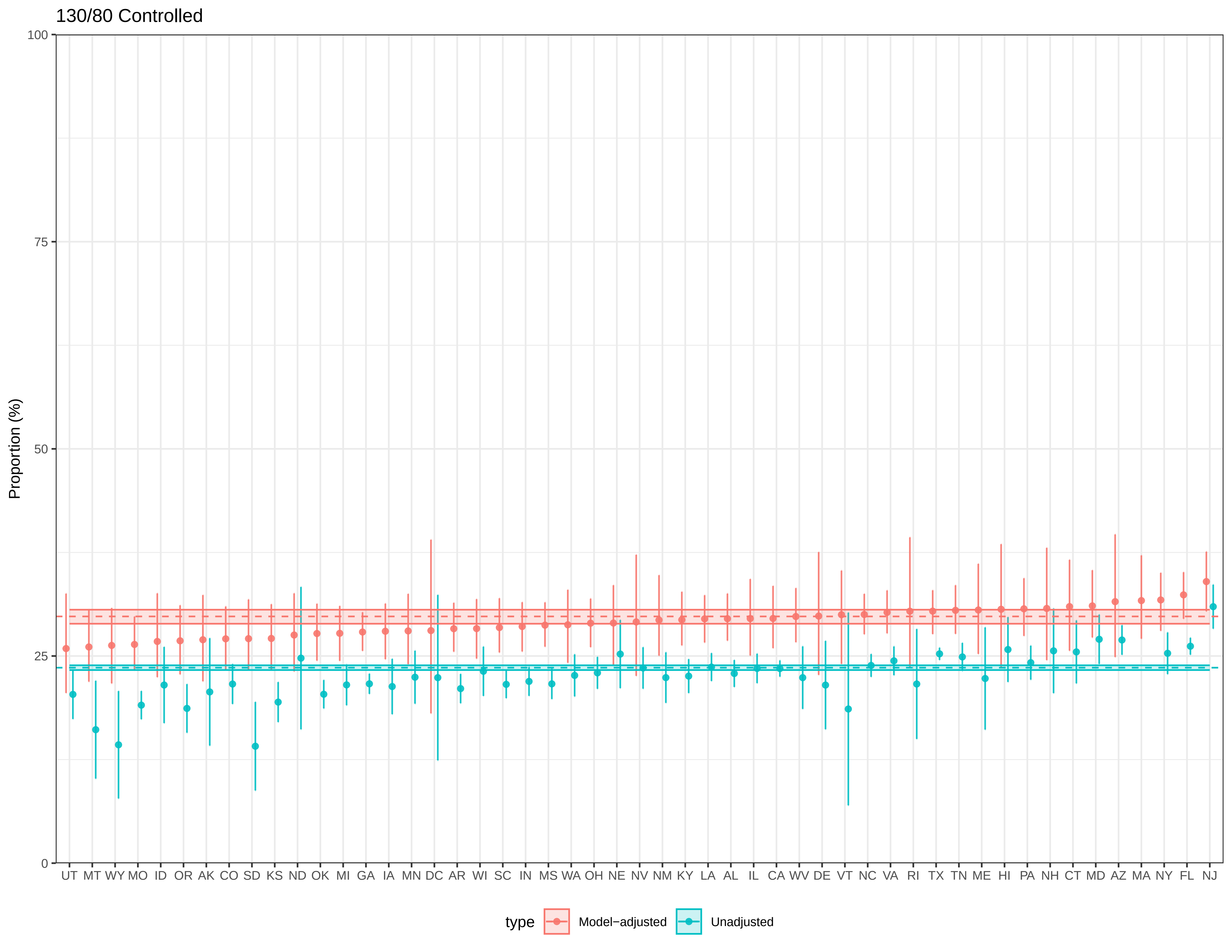
130/80 Controlled
Proportion (%)
100
75
50
25
0
UT MT WY MO ID OR AK CO SD KS ND OK MI GA IA MN DC AR WI SC IN MS WA OH NE NV NM KY LA AL IL CA WV DE VT NC VA RI TX TN ME HI PA NH CT MD AZ MA NY FL NJ
type
Model−adjusted
Unadjusted

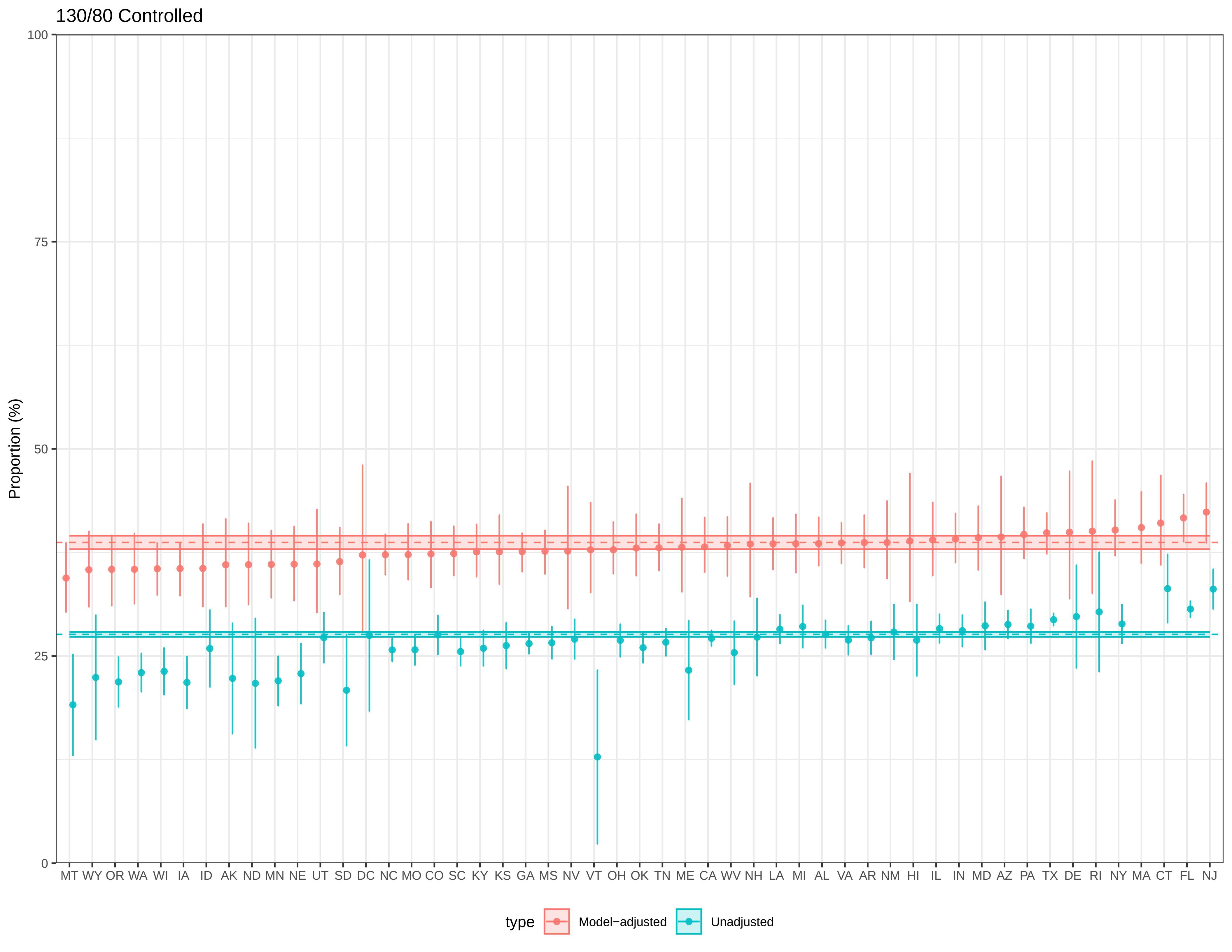

130/80 Controlled
Proportion (%)
100
75
50
25
0
MT WY OR WA WI IA ID AK ND MN NE UT SD DC NC MO CO SC KY KS GA MS NV VT OH OK TN ME CA WV NH LA MI AL VA AR NM HI IL IN MD AZ PA TX DE RI NY MA CT FL NJ
type
Model−adjusted
Unadjusted

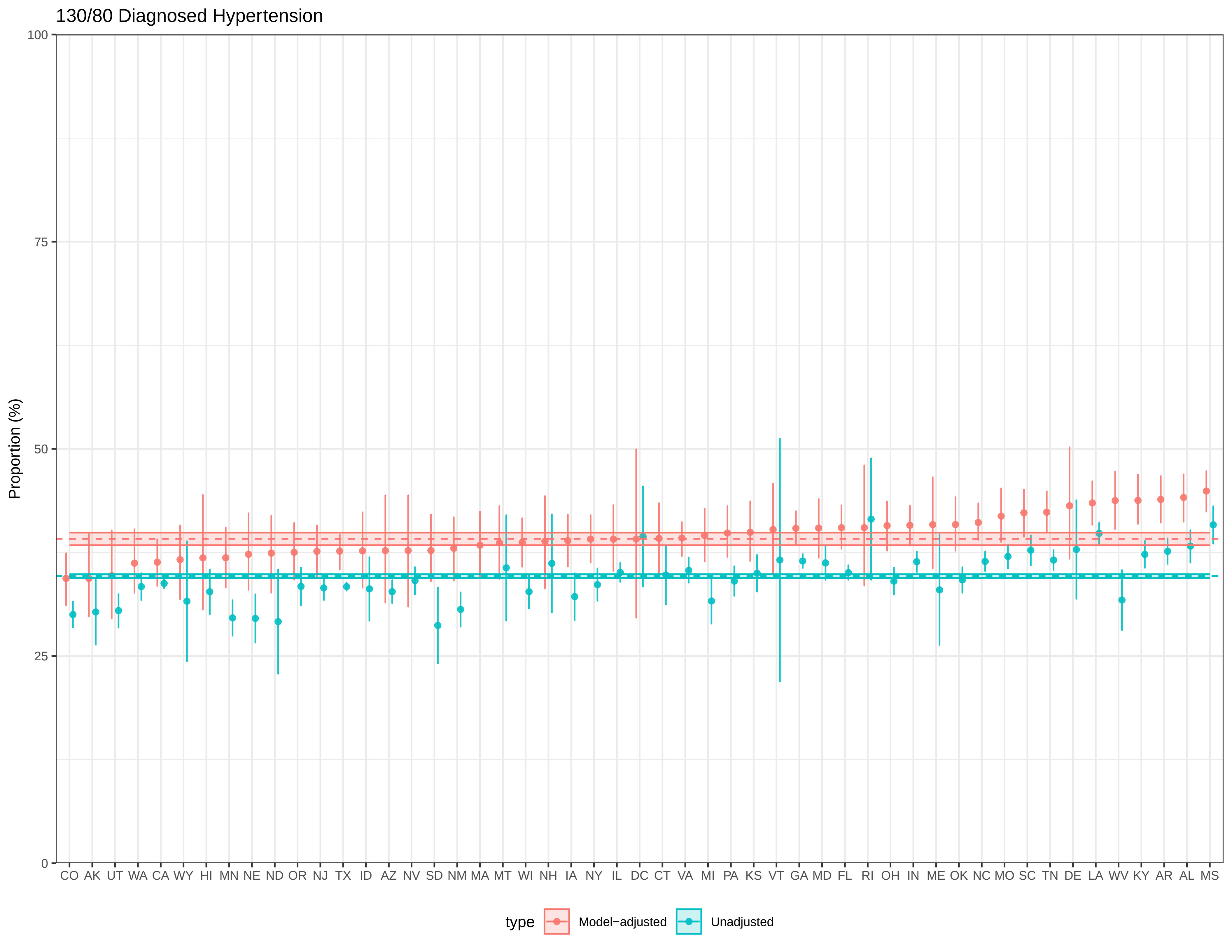
130/80 Diagnosed Hypertension
Proportion (%)
100
75
50
25
0
CO AK UT WA CA WY HI MN NE ND OR NJ TX ID AZ NV SD NM MA MT WI NH IA NY IL DC CT VA MI PA KS VT GA MD FL RI OH IN ME OK NC MO SC TN DE LA WV KY AR AL MS
type
Model−adjusted
Unadjusted

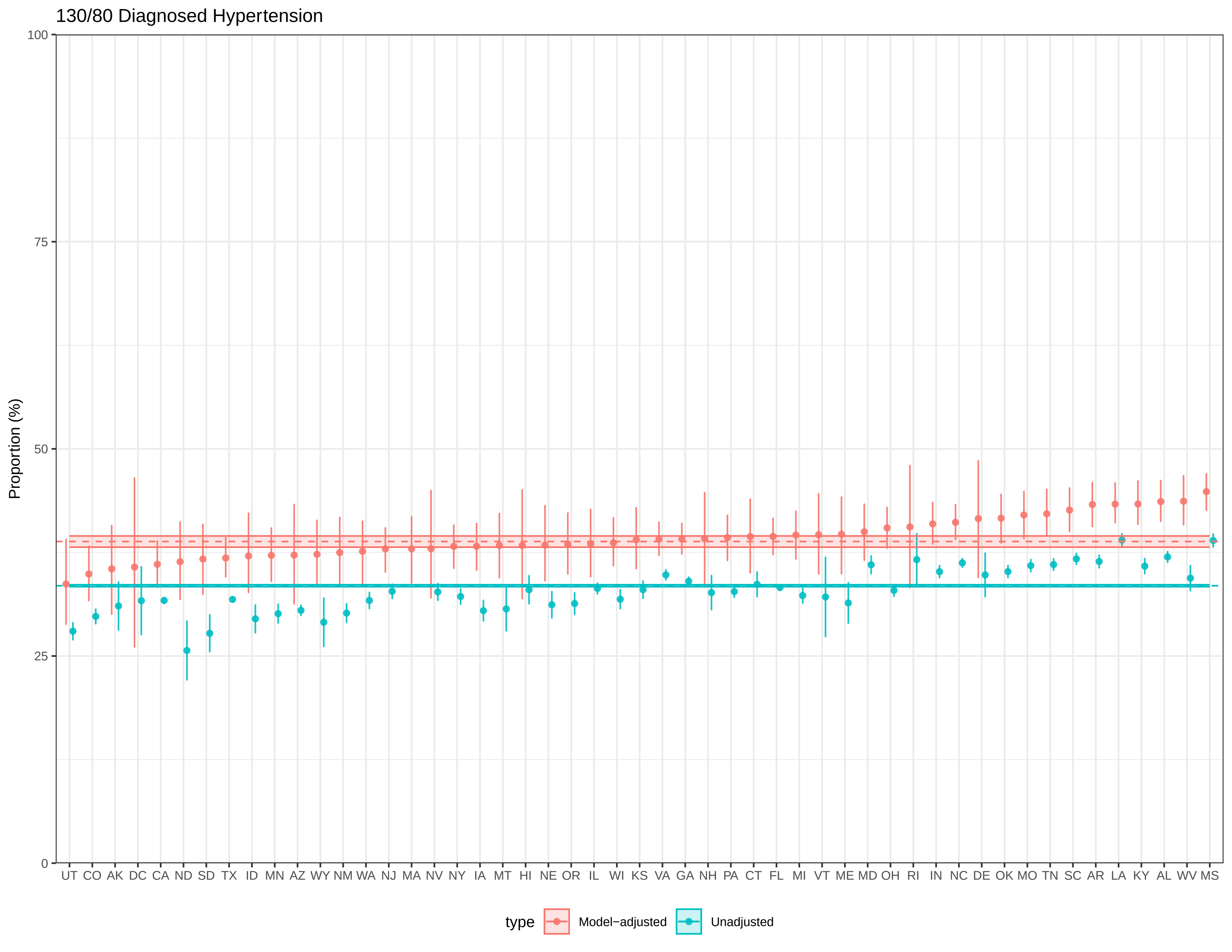
130/80 Diagnosed Hypertension
Proportion (%)
100
75
50
25
0
UT CO AK DC CA ND SD TX ID MN AZ WY NM WA NJ MA NV NY IA MT HI NE OR IL WI KS VA GA NH PA CT FL MI VT ME MD OH RI IN NC DE OK MO TN SC AR LA KY AL WV MS
type
Model−adjusted
Unadjusted

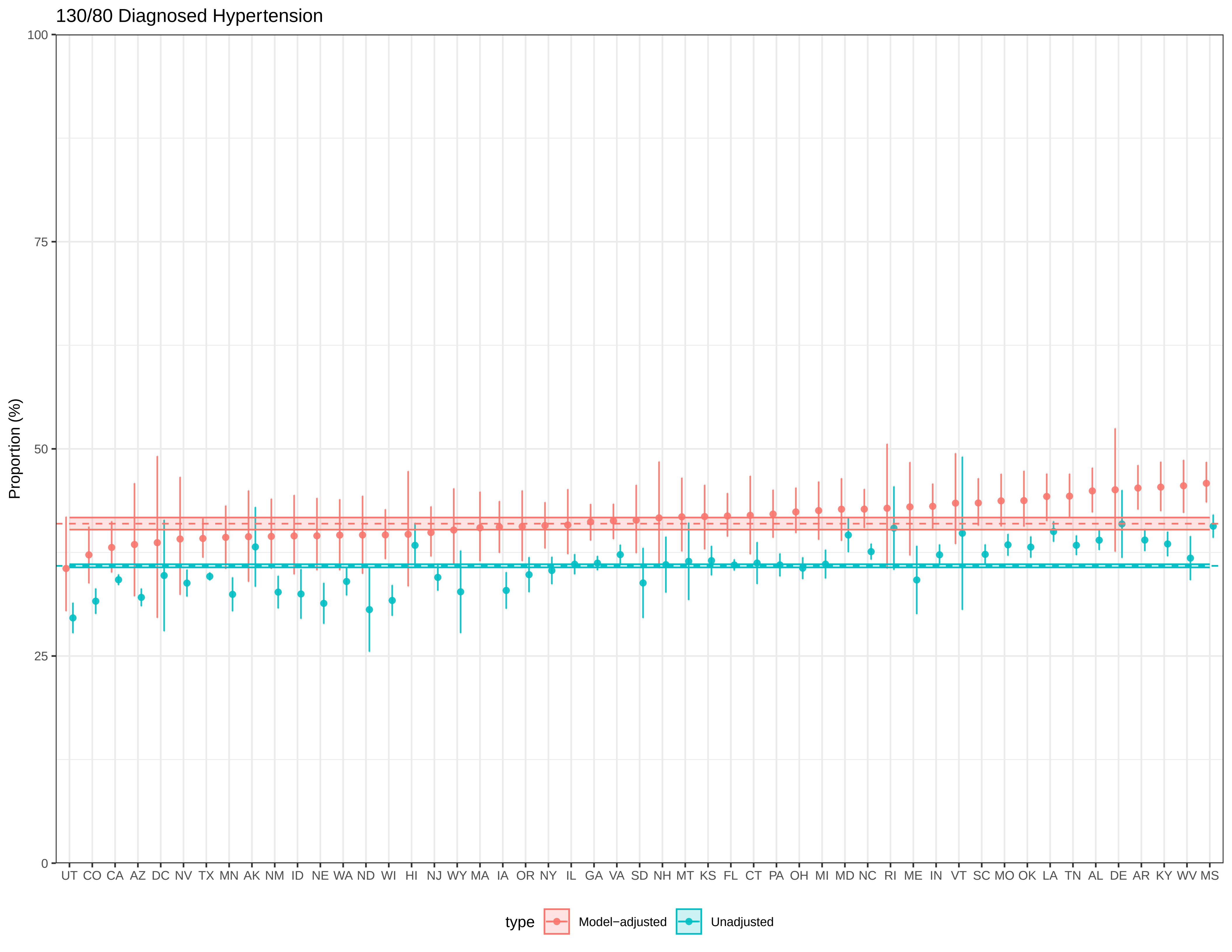

130/80 Diagnosed Hypertension
Proportion (%)
100
75
50
25
0
UT CO CA AZ DC NV TX MN AK NM ID NE WA ND WI HI NJ WY MA IA OR NY IL GA VA SD NH MT KS FL CT PA OH MI MD NC RI ME IN VT SC MO OK LA TN AL DE AR KY WV MS
type
Model−adjusted
Unadjusted

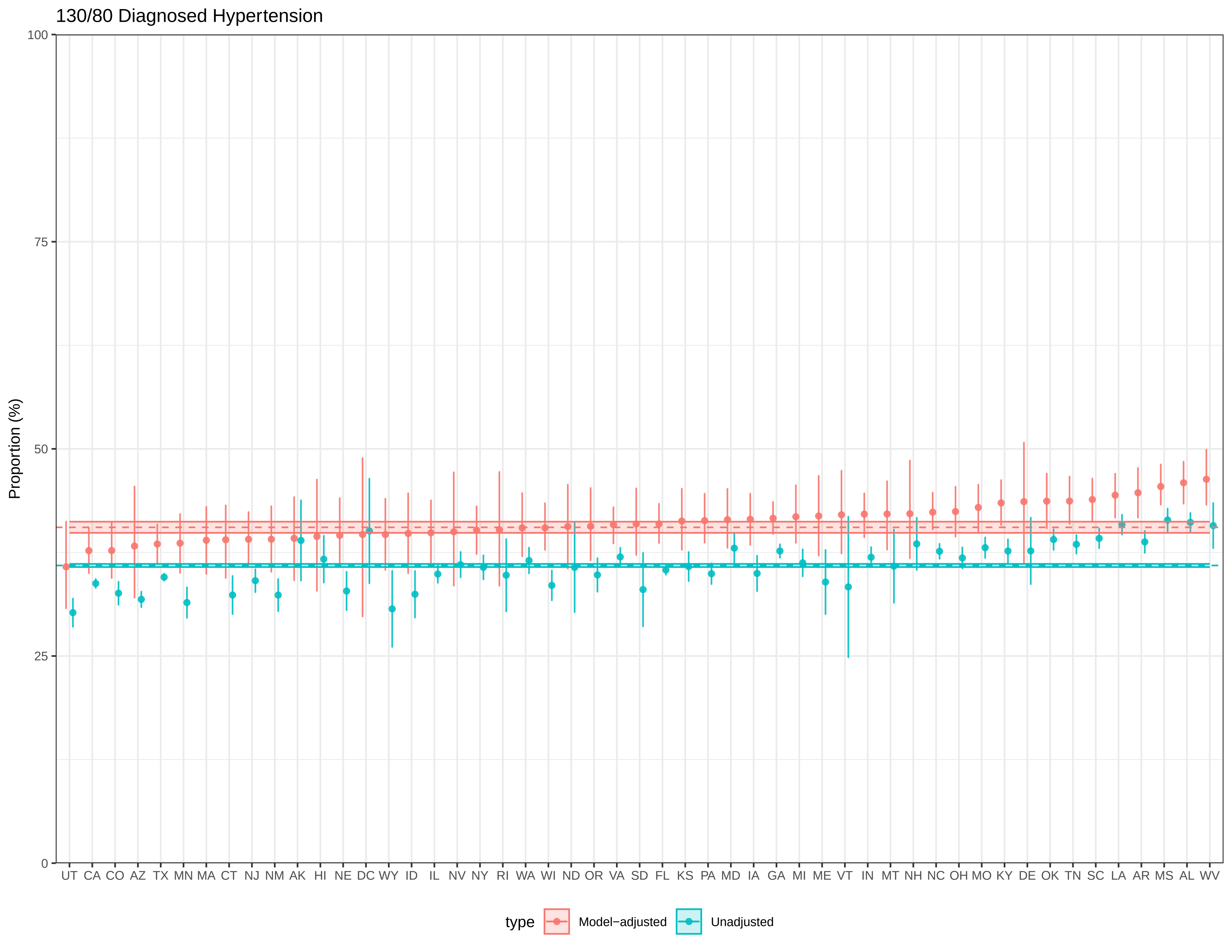

130/80 Diagnosed Hypertension
Proportion (%)
100
75
50
25
0
UT CA CO AZ TX MN MA CT NJ NM AK HI NE DC WY ID IL NV NY RI WA WI ND OR VA SD FL KS PA MD IA GA MI ME VT IN MT NH NC OH MO KY DE OK TN SC LA AR MS AL WV
type
Model−adjusted
Unadjusted

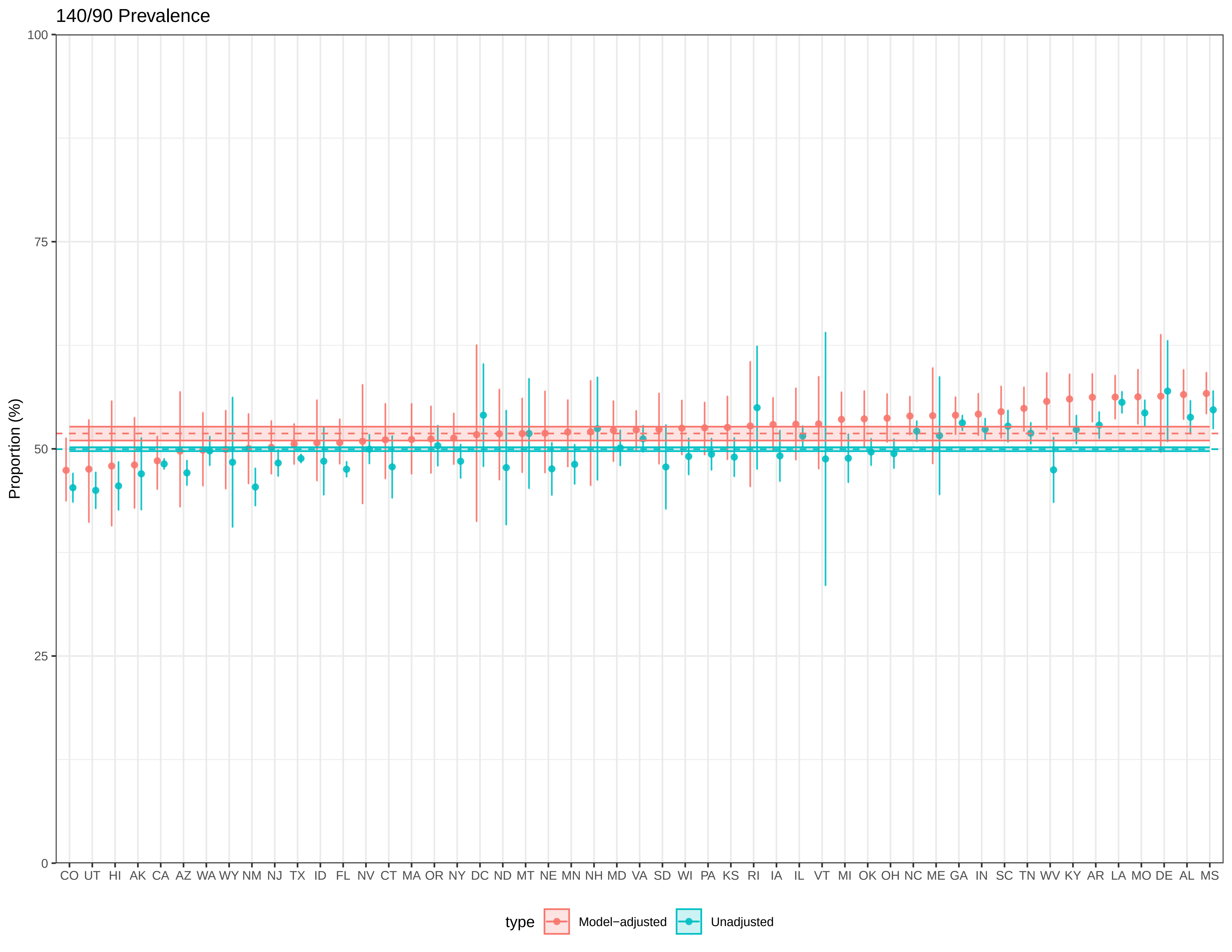
140/90 Prevalence
Proportion (%)
100
75
50
25
0
CO UT HI AK CA AZ WA WY NM NJ TX ID FL NV CT MA OR NY DC ND MT NE MN NH MD VA SD WI PA KS RI IA IL VT MI OK OH NC ME GA IN SC TN WV KY AR LA MO DE AL MS
type
Model−adjusted
Unadjusted

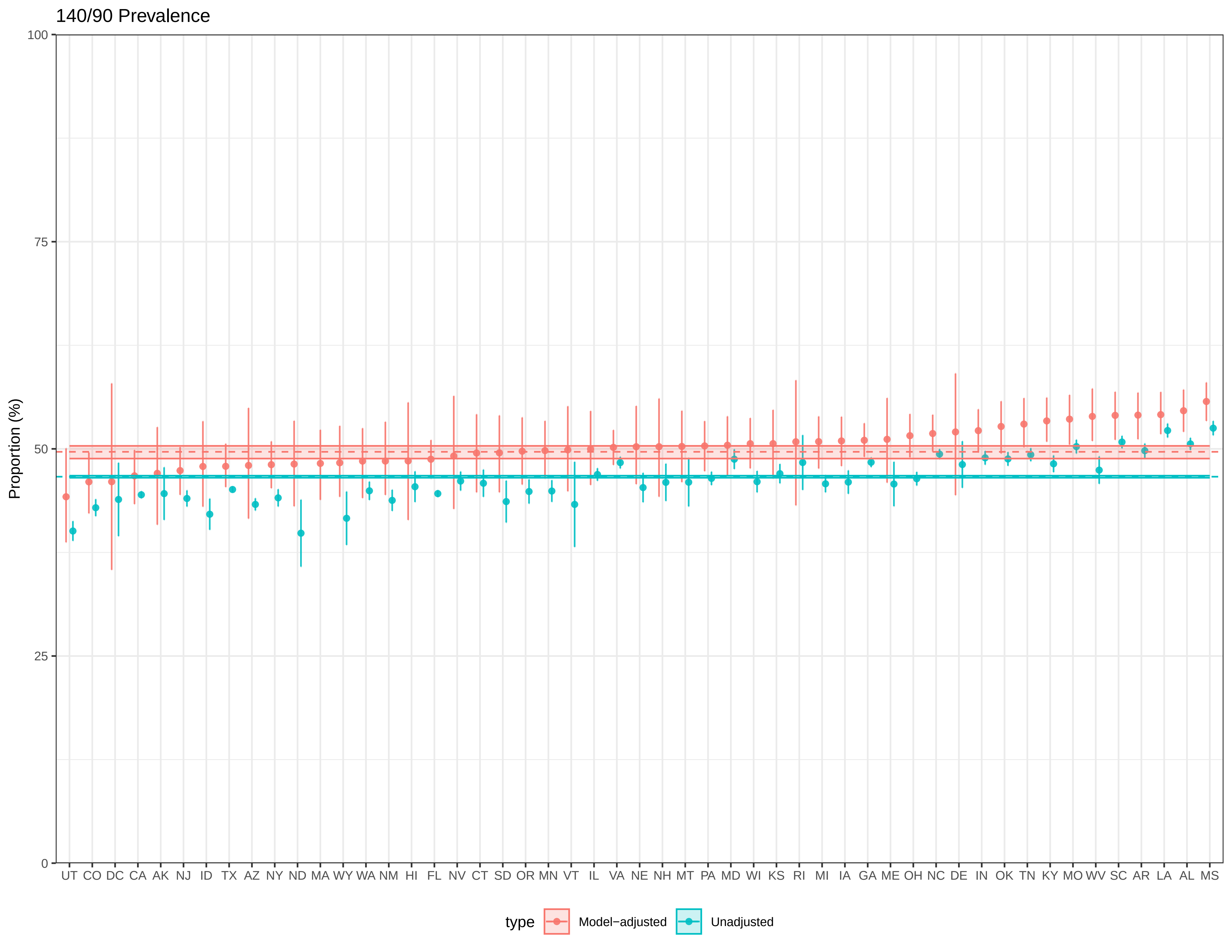

140/90 Prevalence
Proportion (%)
100
75
50
25
0
UT CO DC CA AK NJ ID TX AZ NY ND MA WY WA NM HI FL NV CT SD OR MN VT IL VA NE NH MT PA MD WI KS RI MI IA GA ME OH NC DE IN OK TN KY MO WV SC AR LA AL MS
type
Model−adjusted
Unadjusted

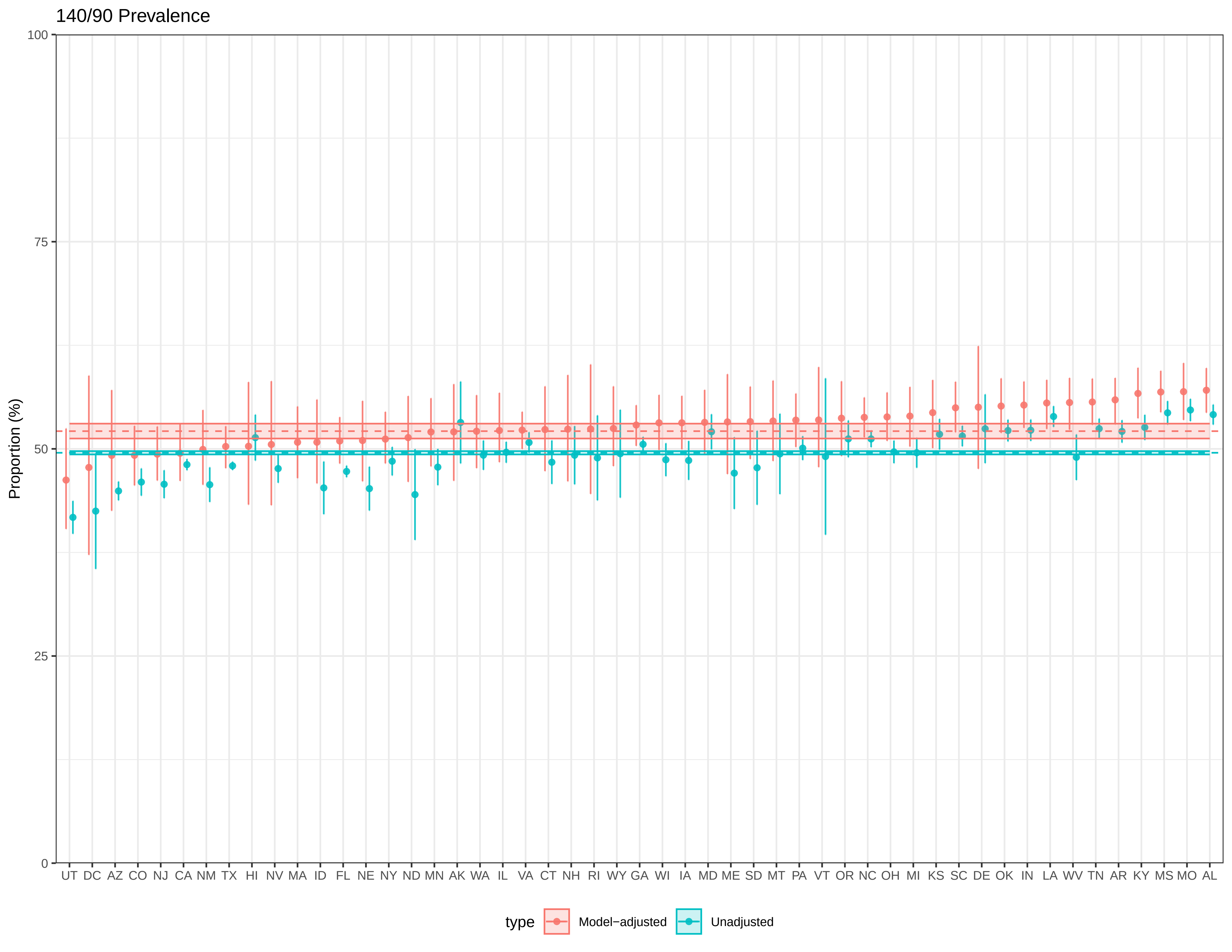

140/90 Prevalence
Proportion (%)
100
75
50
25
0
UT DC AZ CO NJ CA NM TX HI NV MA ID FL NE NY ND MN AK WA IL VA CT NH RI WY GA WI IA MD ME SD MT PA VT OR NC OH MI KS SC DE OK IN LA WV TN AR KY MS MO AL
type
Model−adjusted
Unadjusted

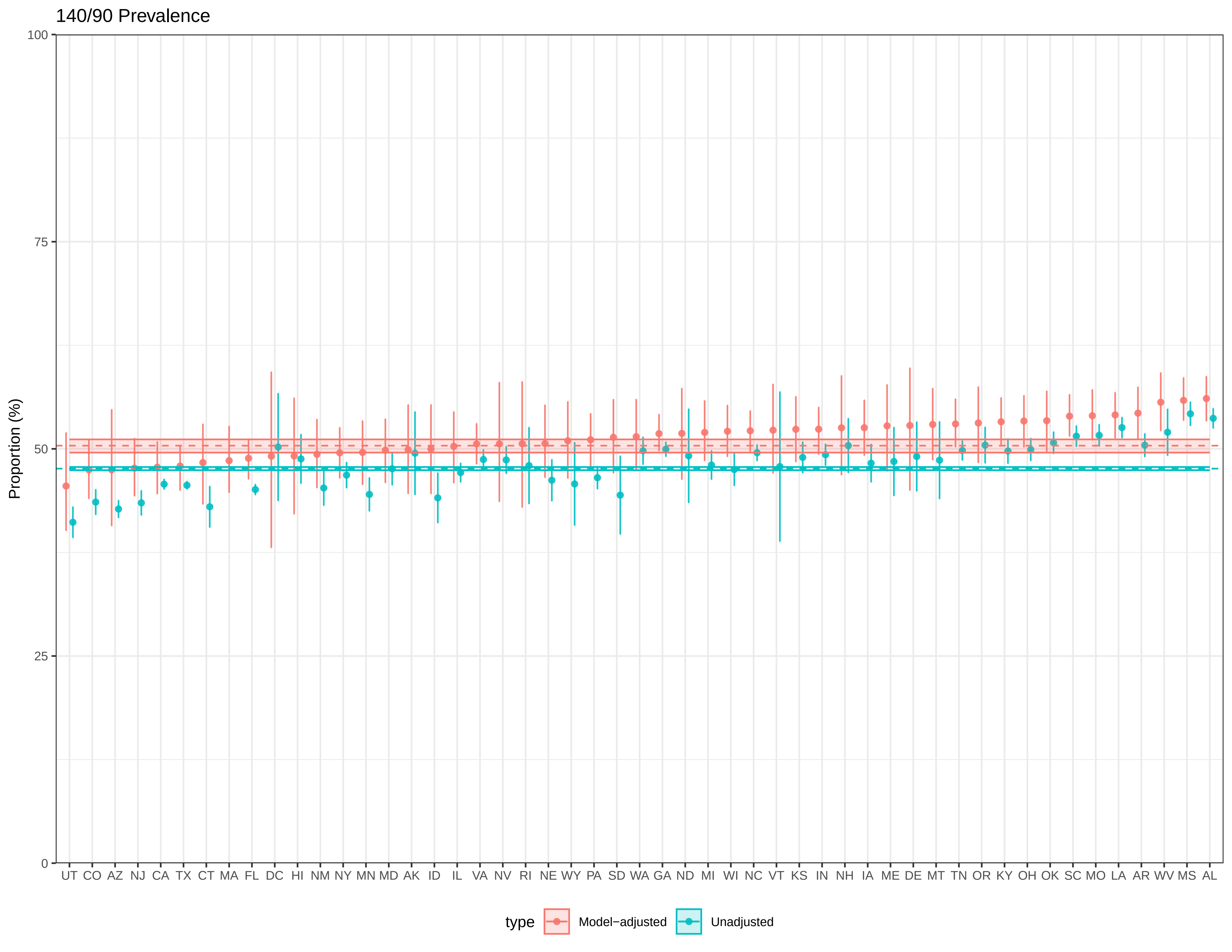

140/90 Prevalence
Proportion (%)
100
75
50
25
0
UT CO AZ NJ CA TX CT MA FL DC HI NM NY MN MD AK ID IL VA NV RI NE WY PA SD WA GA ND MI WI NC VT KS IN NH IA ME DE MT TN OR KY OH OK SC MO LA AR WV MS AL
type
Model−adjusted
Unadjusted

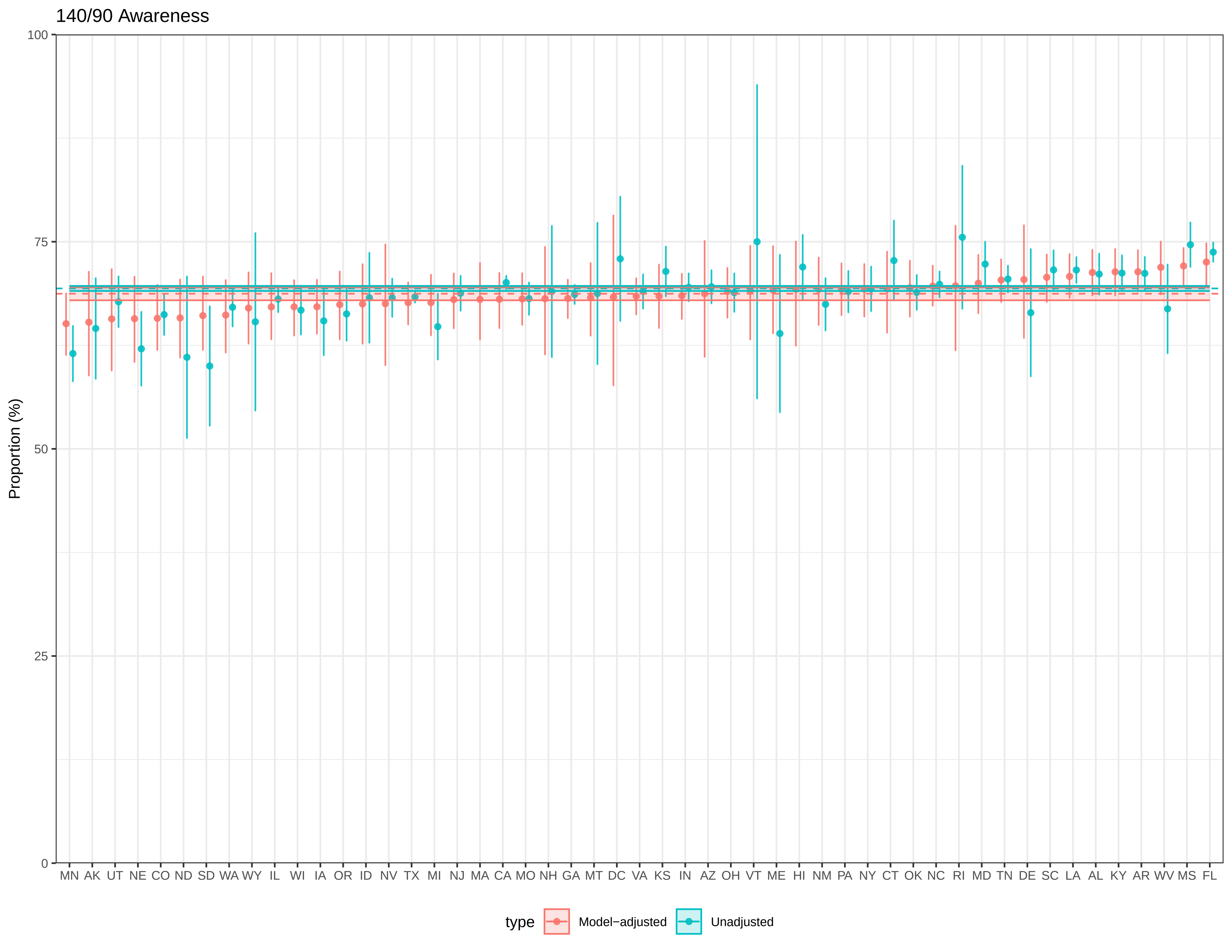

140/90 Awareness
Proportion (%)
100
75
50
25
0
MN AK UT NE CO ND SD WA WY IL WI IA OR ID NV TX MI NJ MA CA MO NH GA MT DC VA KS IN AZ OH VT ME HI NM PA NY CT OK NC RI MD TN DE SC LA AL KY AR WV MS FL
type
Model−adjusted
Unadjusted

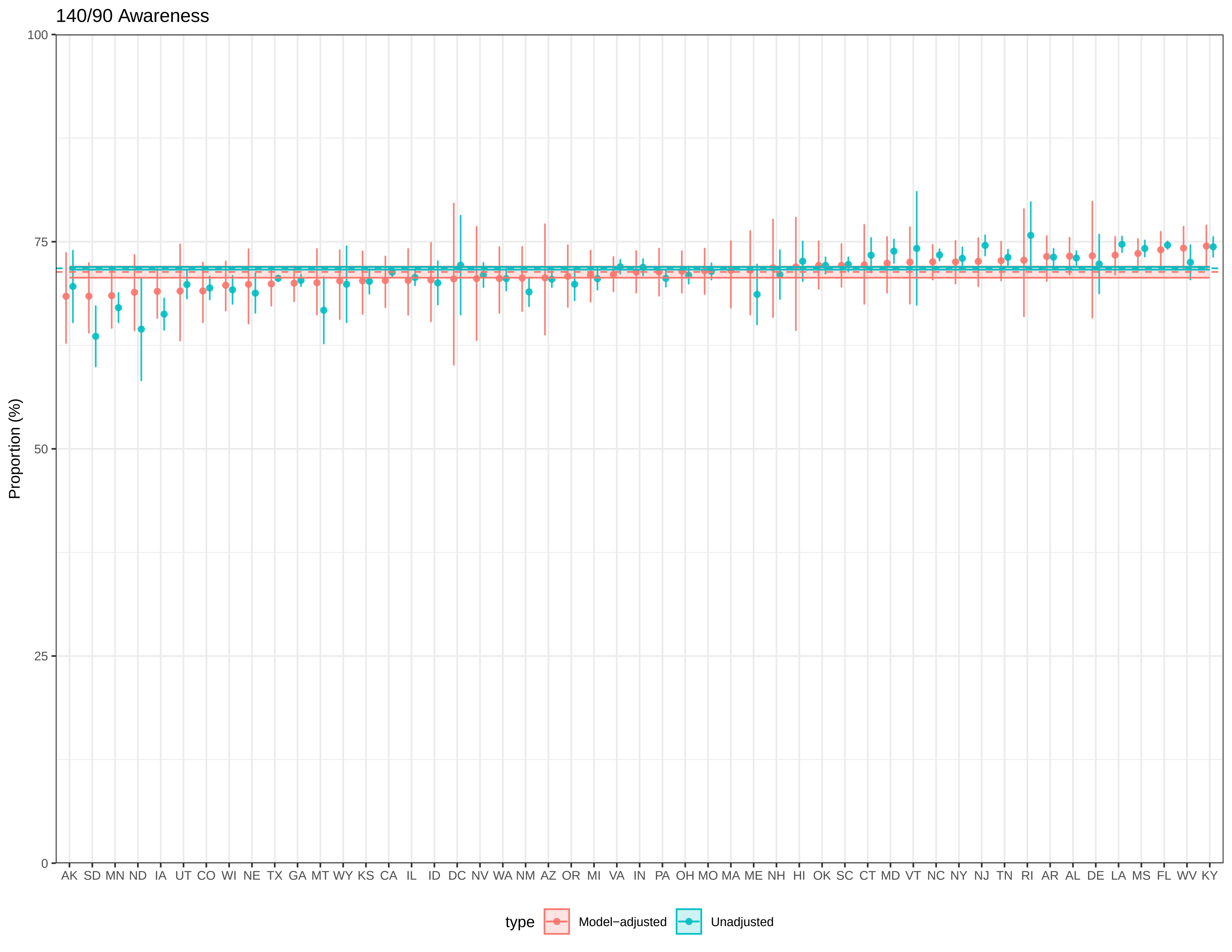

140/90 Awareness
Proportion (%)
100
75
50
25
0
AK SD MN ND IA UT CO WI NE TX GA MT WY KS CA IL ID DC NV WA NM AZ OR MI VA IN PA OH MO MA ME NH HI OK SC CT MD VT NC NY NJ TN RI AR AL DE LA MS FL WV KY
type
Model−adjusted
Unadjusted

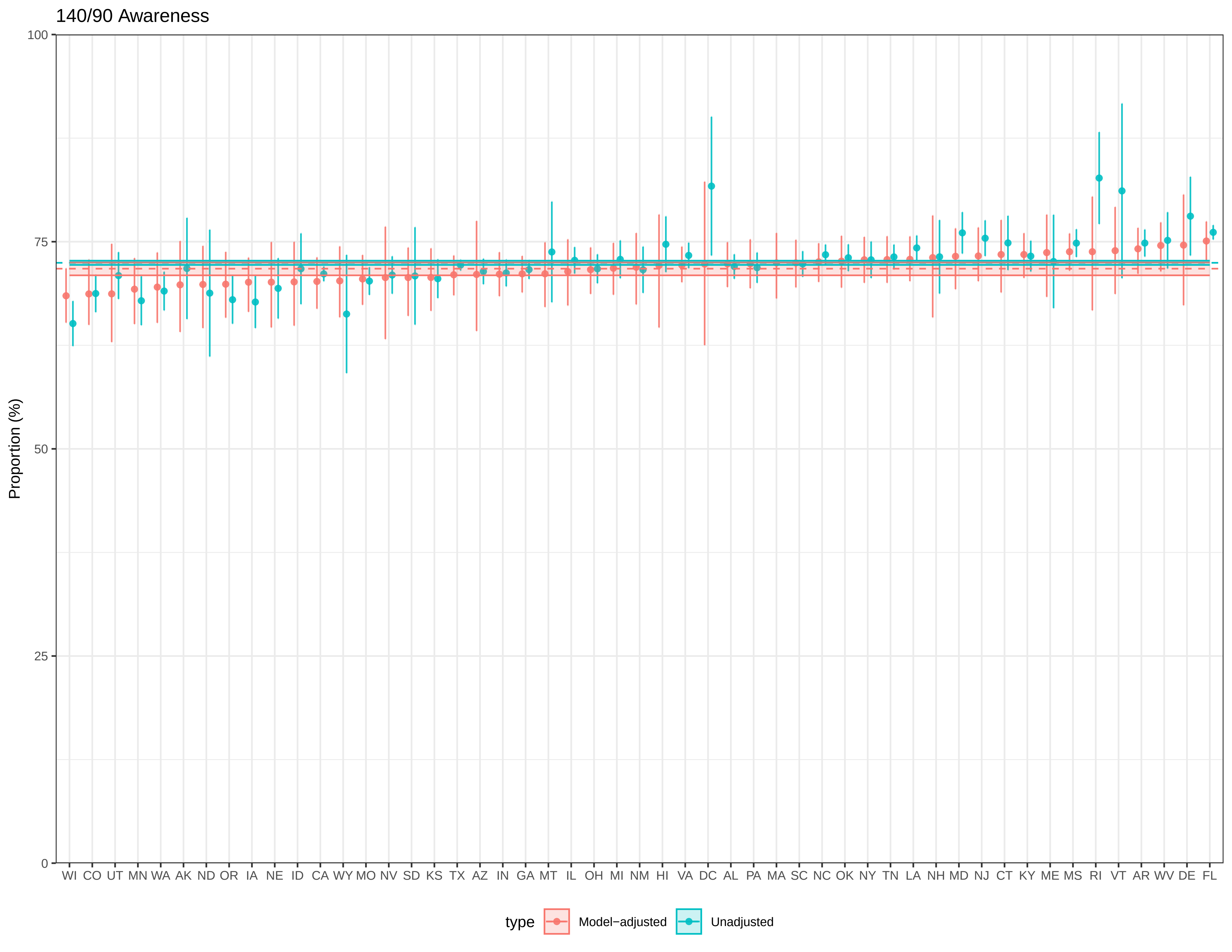

140/90 Awareness
Proportion (%)
100
75
50
25
0
WI CO UT MN WA AK ND OR IA NE ID CA WY MO NV SD KS TX AZ IN GA MT IL OH MI NM HI VA DC AL PA MA SC NC OK NY TN LA NH MD NJ CT KY ME MS RI VT AR WV DE FL
type
Model−adjusted
Unadjusted

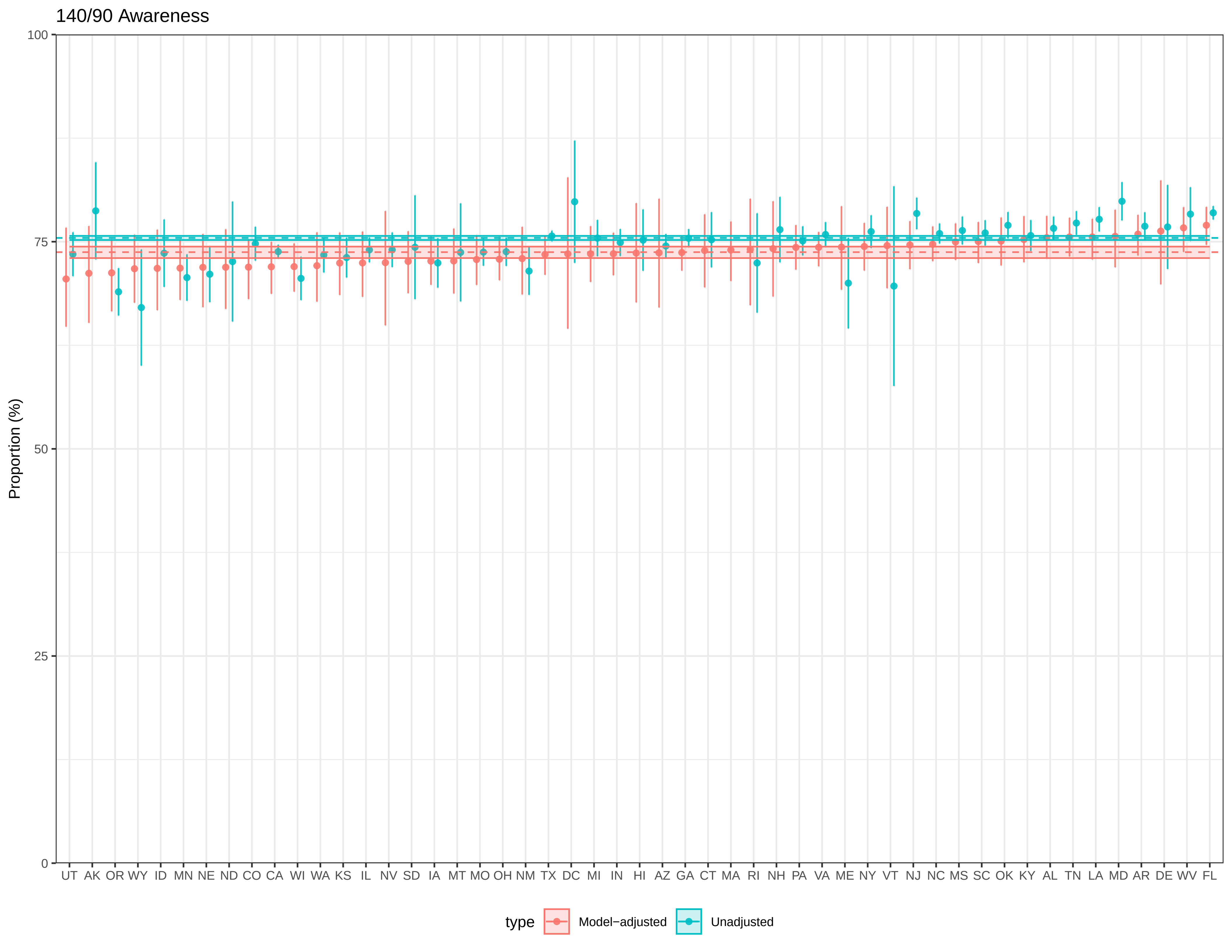

140/90 Awareness
Proportion (%)
100
75
50
25
0
UT AK OR WY ID MN NE ND CO CA WI WA KS IL NV SD IA MT MO OH NM TX DC MI IN HI AZ GA CT MA RI NH PA VA ME NY VT NJ NC MS SC OK KY AL TN LA MD AR DE WV FL
type
Model−adjusted
Unadjusted

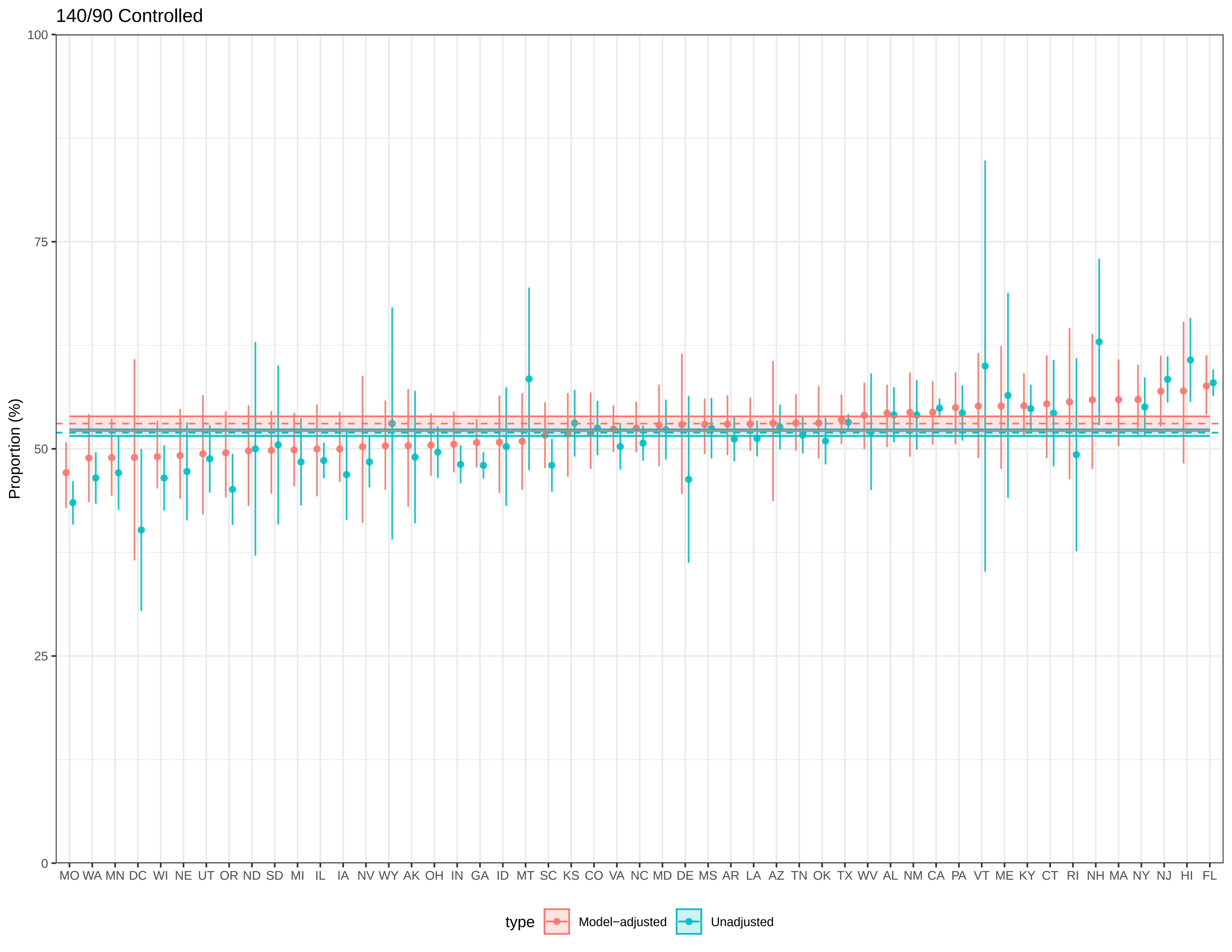
140/90 Controlled
Proportion (%)
100
75
50
25
0
MO WA MN DC WI NE UT OR ND SD MI IL IA NV WY AK OH IN GA ID MT SC KS CO VA NC MD DE MS AR LA AZ TN OK TX WV AL NM CA PA VT ME KY CT RI NH MA NY NJ HI FL
type
Model−adjusted
Unadjusted

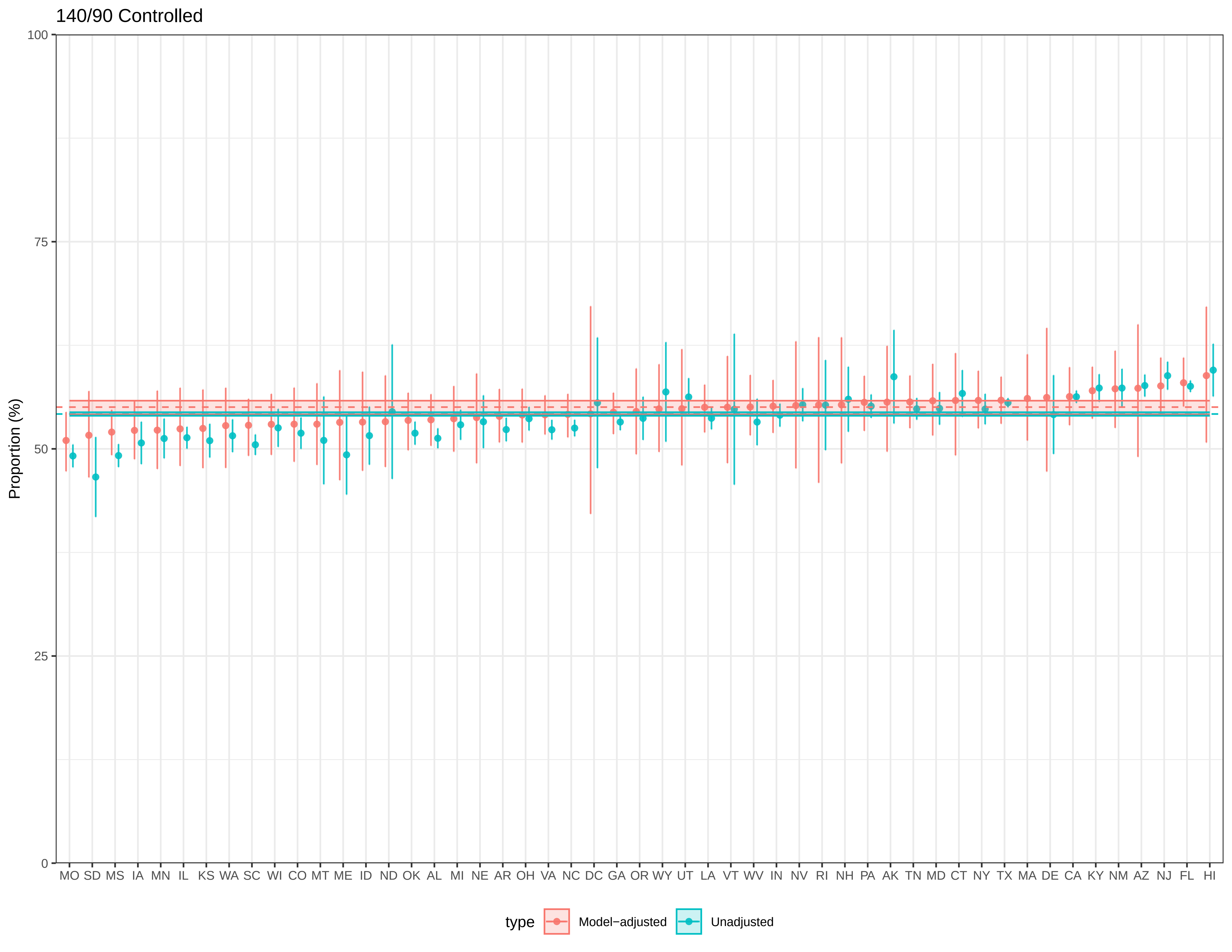

140/90 Controlled
Proportion (%)
100
75
50
25
0
MO SD MS IA MN IL KS WA SC WI CO MT ME ID ND OK AL MI NE AR OH VA NC DC GA OR WY UT LA VT WV IN NV RI NH PA AK TN MD CT NY TX MA DE CA KY NM AZ NJ FL HI
type
Model−adjusted
Unadjusted

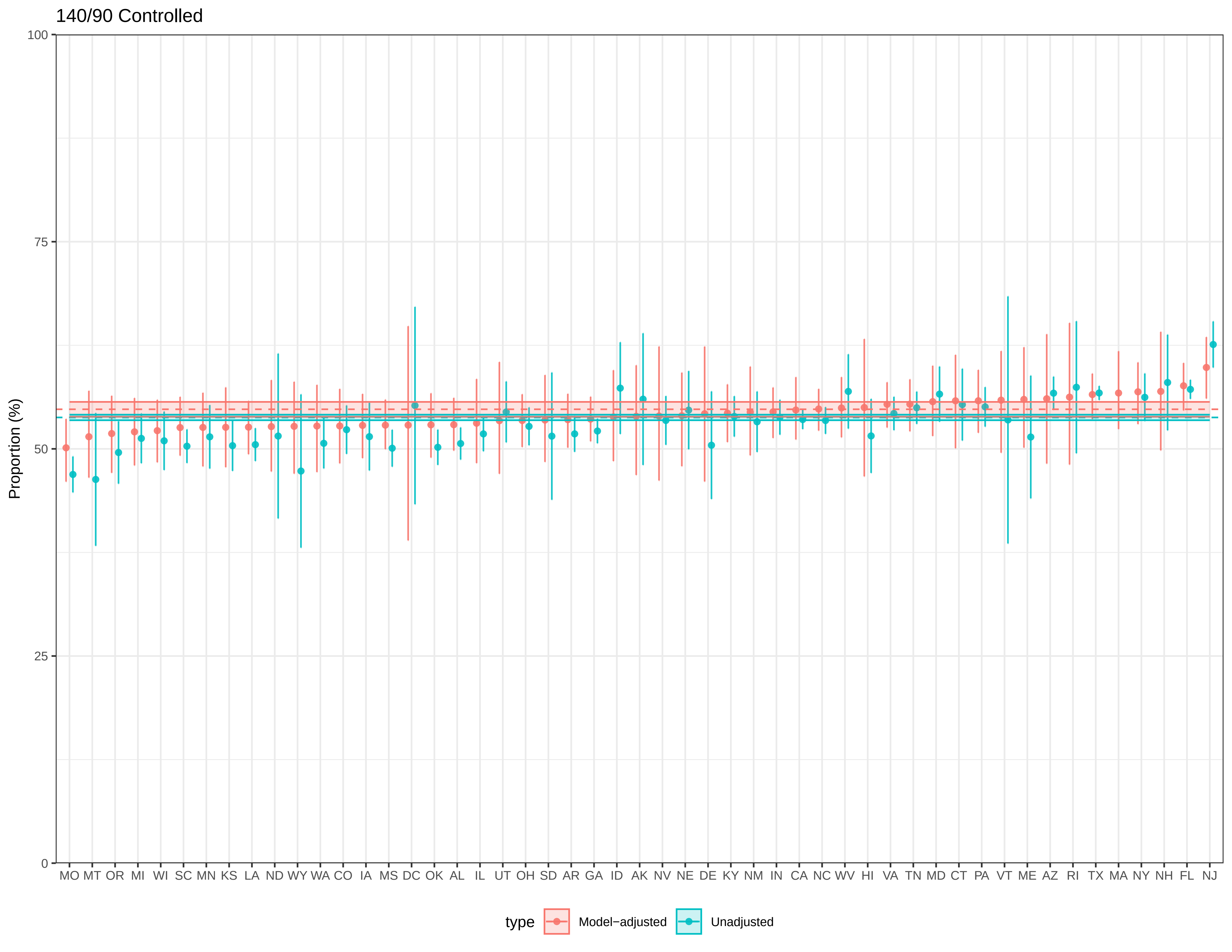

140/90 Controlled
Proportion (%)
100
75
50
25
0
MO MT OR MI WI SC MN KS LA ND WY WA CO IA MS DC OK AL IL UT OH SD AR GA ID AK NV NE DE KY NM IN CA NC WV HI VA TN MD CT PA VT ME AZ RI TX MA NY NH FL NJ
type
Model−adjusted
Unadjusted

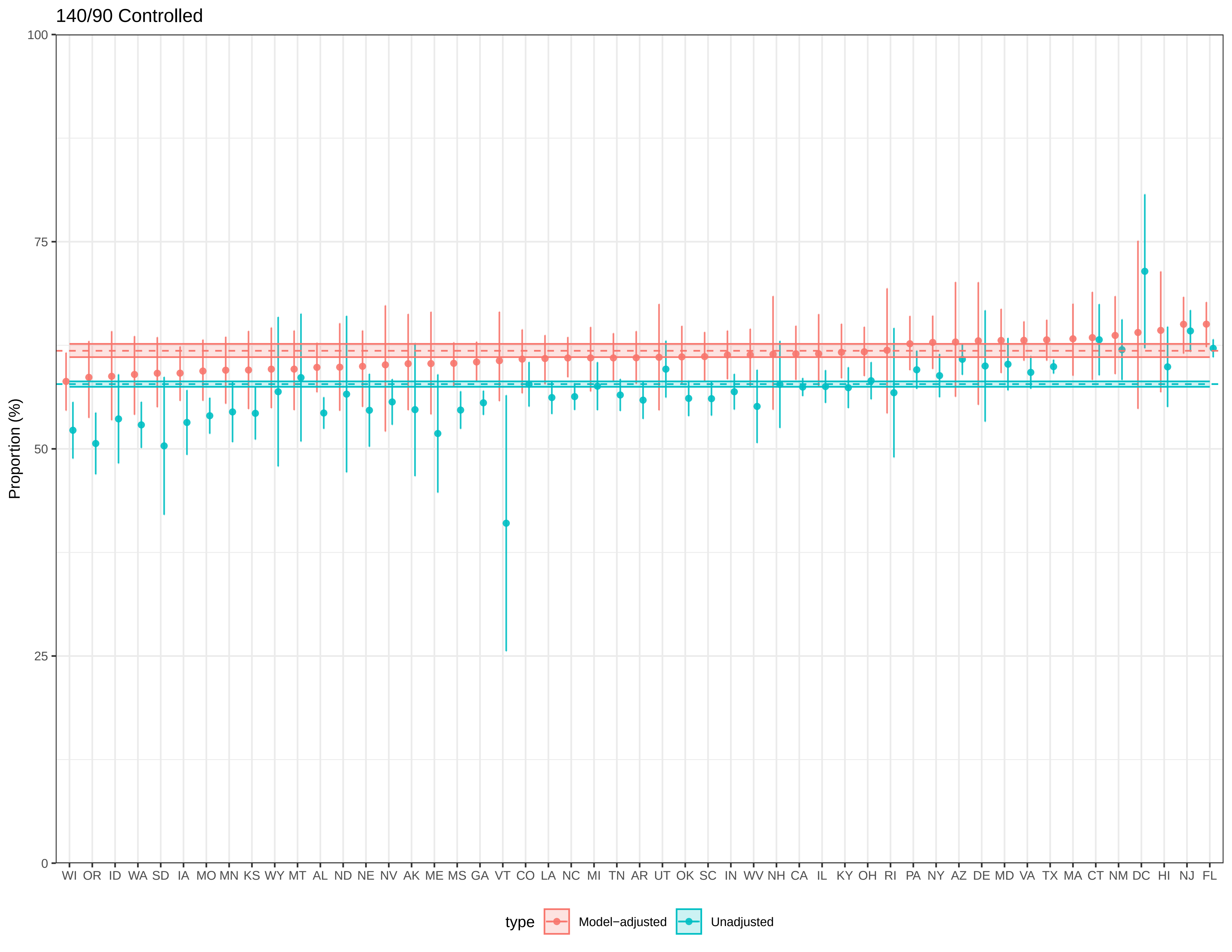
140/90 Controlled
Proportion (%)
100
75
50
25
0
WI OR ID WA SD IA MO MN KS WY MT AL ND NE NV AK ME MS GA VT CO LA NC MI TN AR UT OK SC IN WV NH CA IL KY OH RI PA NY AZ DE MD VA TX MA CT NM DC HI NJ FL
type
Model−adjusted
Unadjusted

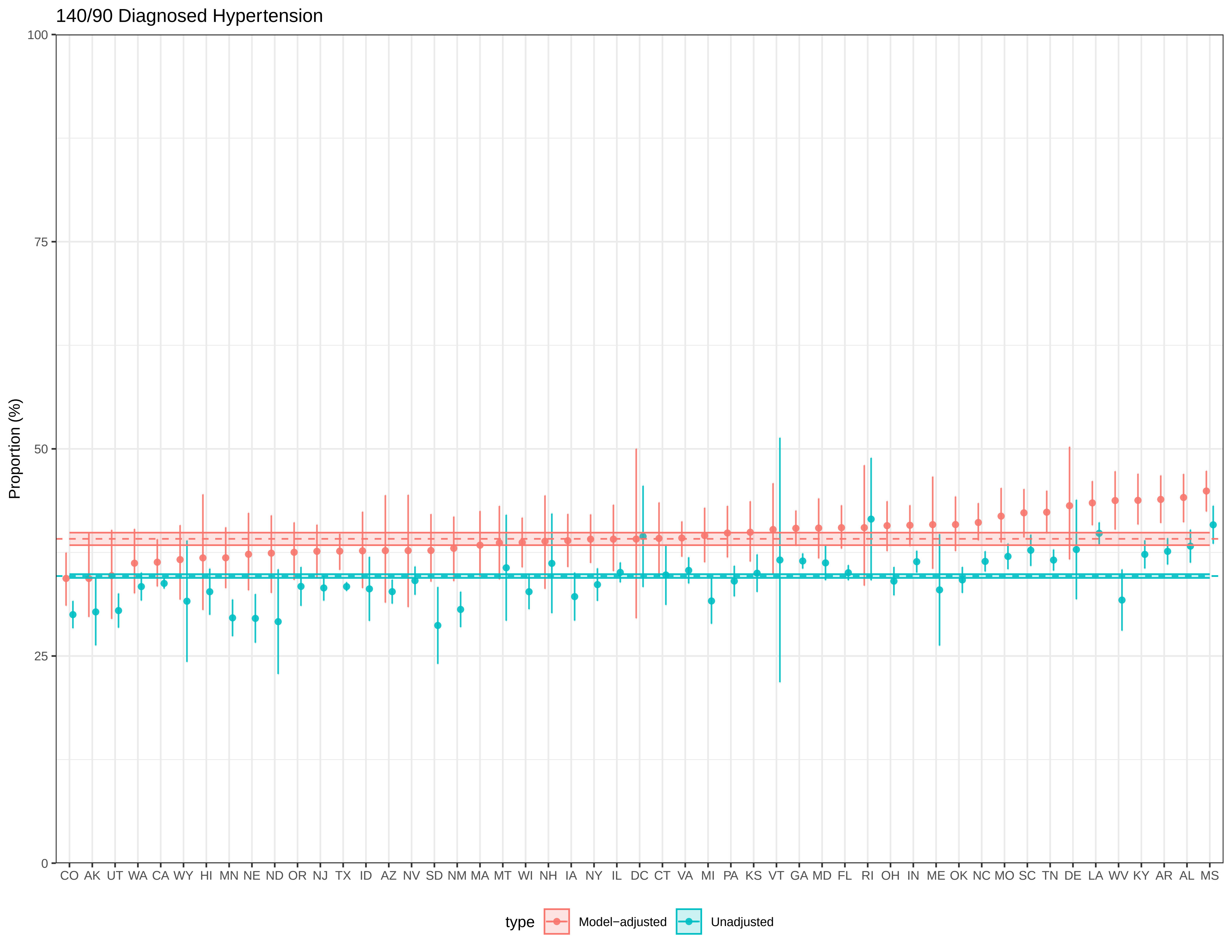
140/90 Diagnosed Hypertension
Proportion (%)
100
75
50
25
0
CO AK UT WA CA WY HI MN NE ND OR NJ TX ID AZ NV SD NM MA MT WI NH IA NY IL DC CT VA MI PA KS VT GA MD FL RI OH IN ME OK NC MO SC TN DE LA WV KY AR AL MS
type
Model−adjusted
Unadjusted

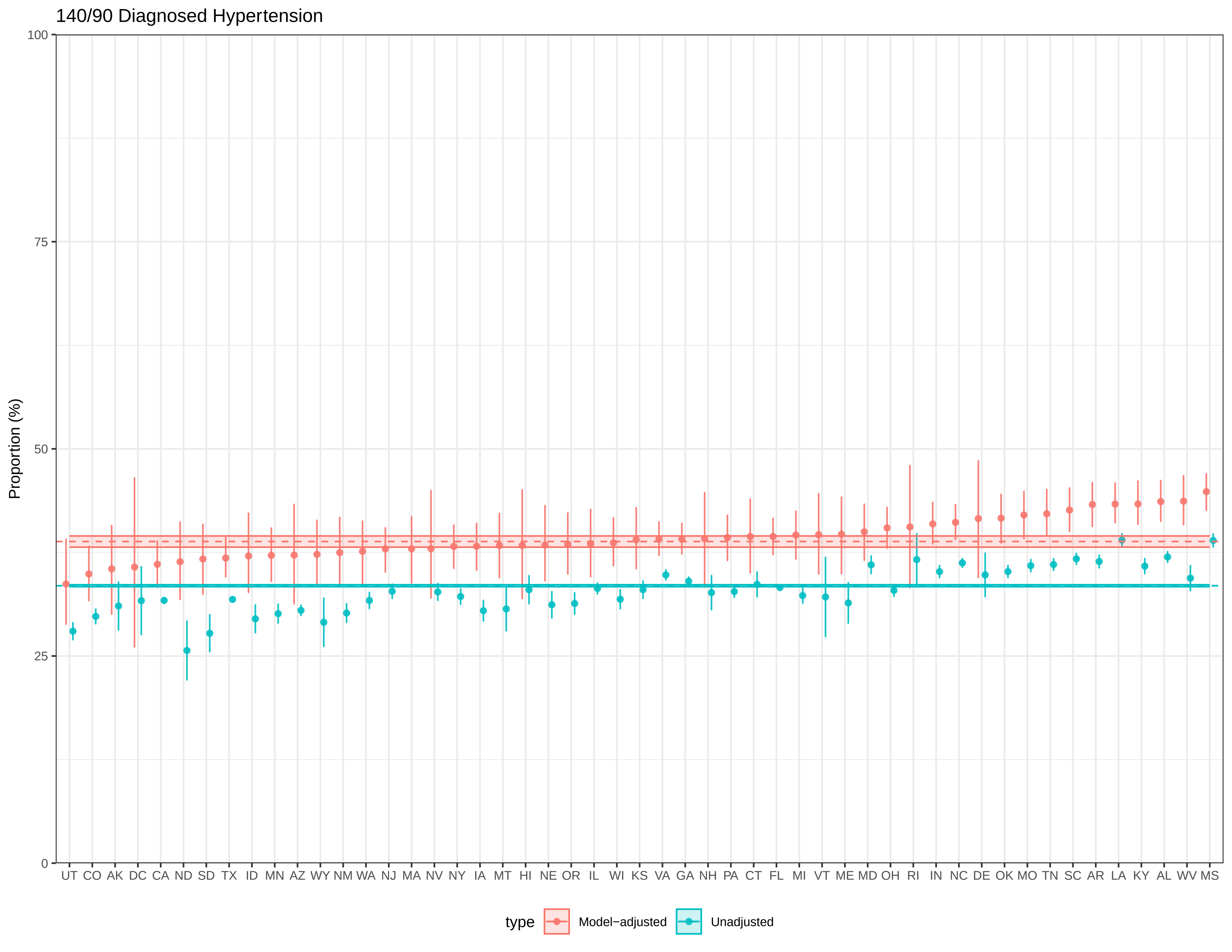
140/90 Diagnosed Hypertension
Proportion (%)
100
75
50
25
0
UT CO AK DC CA ND SD TX ID MN AZ WY NM WA NJ MA NV NY IA MT HI NE OR IL WI KS VA GA NH PA CT FL MI VT ME MD OH RI IN NC DE OK MO TN SC AR LA KY AL WV MS
type
Model−adjusted
Unadjusted

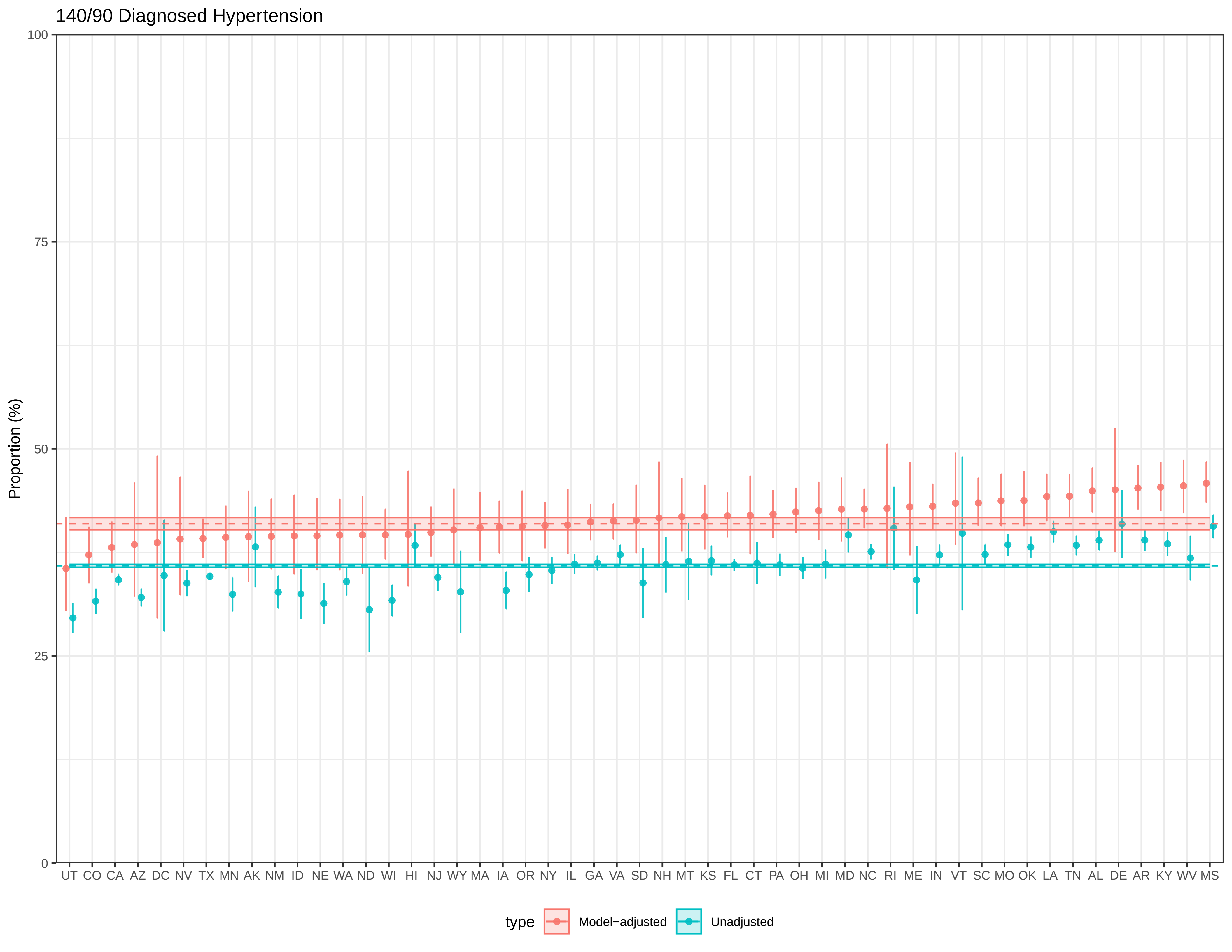
140/90 Diagnosed Hypertension
Proportion (%)
100
75
50
25
0
UT CO CA AZ DC NV TX MN AK NM ID NE WA ND WI HI NJ WY MA IA OR NY IL GA VA SD NH MT KS FL CT PA OH MI MD NC RI ME IN VT SC MO OK LA TN AL DE AR KY WV MS
type
Model−adjusted
Unadjusted

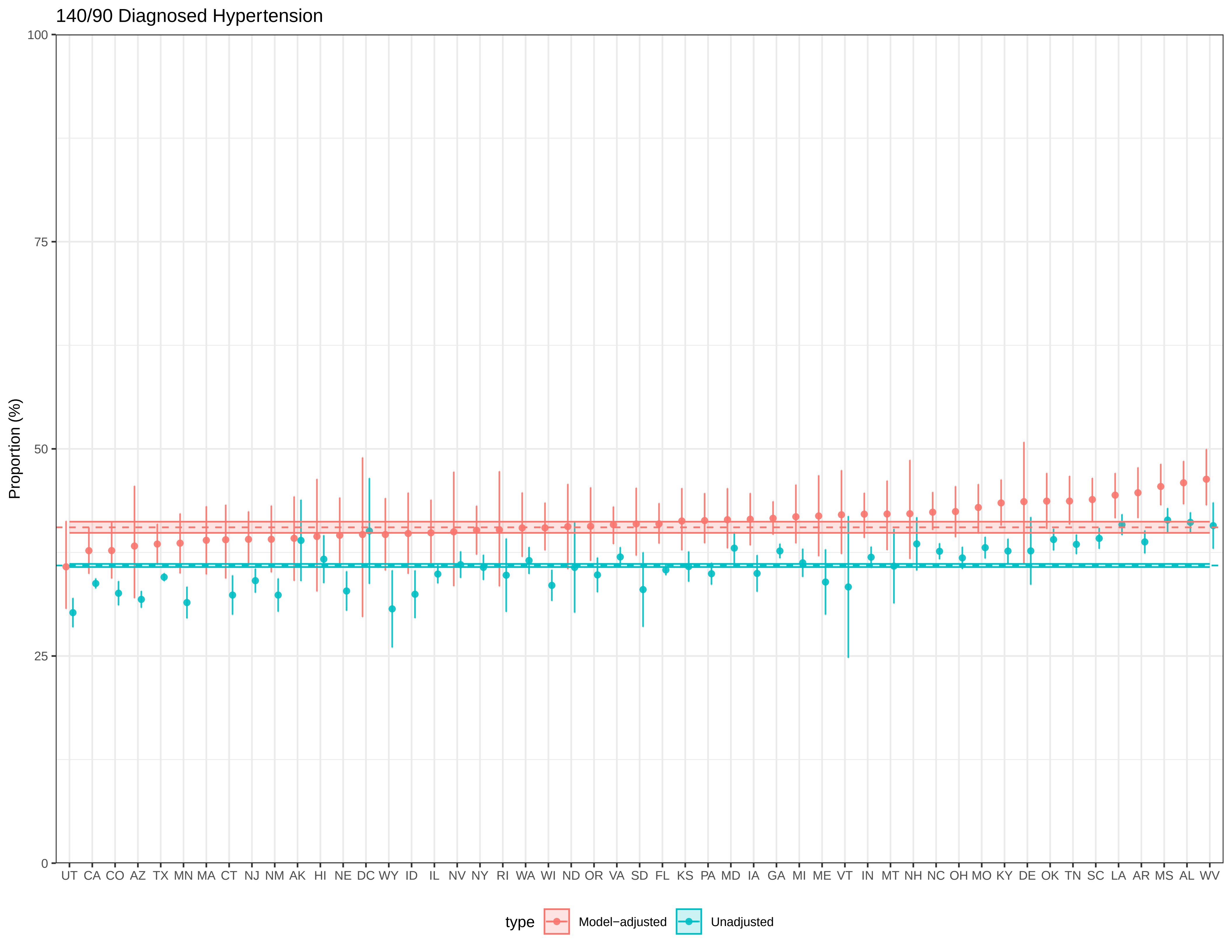

140/90 Diagnosed Hypertension
Proportion (%)
100
75
50
25
0
UT CA CO AZ TX MN MA CT NJ NM AK HI NE DC WY ID IL NV NY RI WA WI ND OR VA SD FL KS PA MD IA GA MI ME VT IN MT NH NC OH MO KY DE OK TN SC LA AR MS AL WV
type
Model−adjusted
Unadjusted
