## Supplementary File 5 for "County Prevalence and Control of High Blood Pressure from Health Kiosks, 2017-2024"

**Online Only Supplementary Material 1**

**Online Only Supplementary Material 2: Estimates of prevalence, awareness, control and diagnosed hypertension by state and county** (separate file)

**Online Only Supplementary Material 3: Precision of county-level estimates from Supplementary Material 2** (separate file)

**Online Only Supplementary Material 4: Pursuant national estimate barplots by age, gender, and race & ethnicity** (separate file)

**Online Only Supplementary Material 5: Comparison of direct- and model-based estimates for states** (separate file)

**Figure S1: Local Moran's I for BRFSS 2021 and Pursuant 2021-2022**

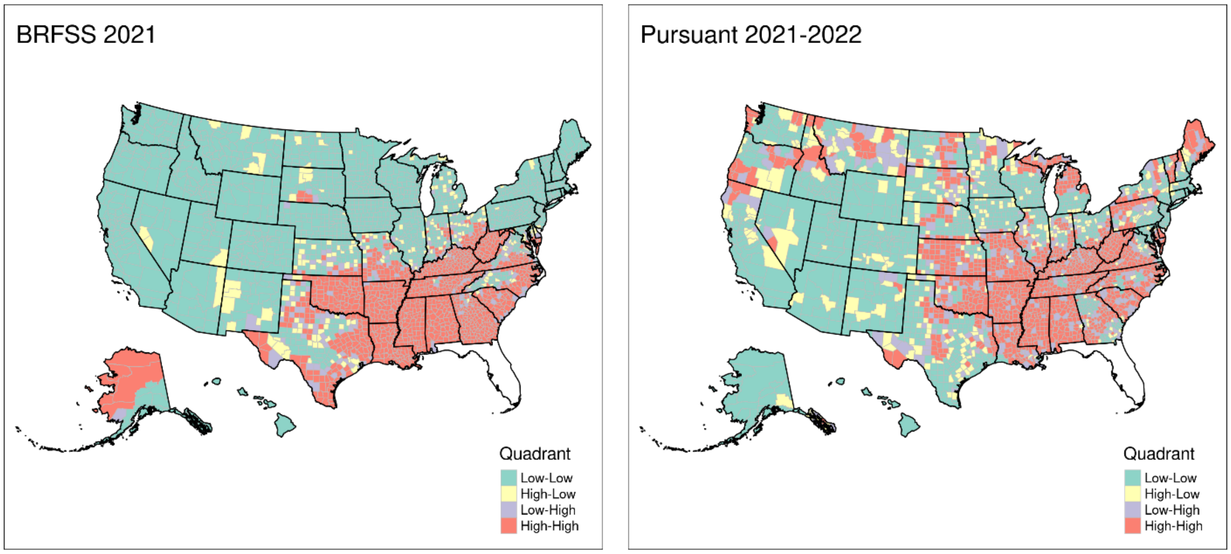

Local Moran's I classifications for the county-level mean estimates of diagnosed hypertension for BRFSS 2021 and Pursuant 2021-2022.

**Supplementary Methods Note**

**Statistical Analysis**

### Multilevel regression and poststratification (MRP)

The logistic MRP mixed effects model fit on the analytic sample for each outcome is as follows. Let $Y_i$ be a binary indicator for high blood pressure (BP) or other outcome in this study for subject $i$. Let $c[i]$ and $s[i]$ be the county and state indices marking the location for subject $i$, and $X_i$ be the covariate vector containing individual (age group, gender, race/ethnicity, gender and race/ethnicity interaction), county (% unemployed, % without HS education, median household income ($)), and state level fixed effects (region of US) for subject $i$. The mixed effects model was then specified as:

$$P(Y_i = 1) = invlogit\left(\alpha^{county}_{c[i],s[i]} + \alpha^{state}_{s[i]} + X_i^{\top}\beta\right),$$
$$\alpha^{county}_{c[i],s[i]} \sim N(0, \sigma_c^2),$$
$$\alpha^{state}_{s[i]} \sim N(0, \sigma_s^2).$$

Models were fit in `R 4.4.1` with the `lme4` package. Once the model was estimated, 500 predictions were made for each unique subpopulation to be used in the poststratification step using draws generated from bootstrap samples with the `merTools` package. The variables identifying each subpopulation were county, state, age group (18-19, 20-44, 45-64, 65+), sex (male, female), and race/ethnicity (Hispanic, NH White, NH Black, NH Asian, NH Other). The post-stratification estimate for high BP prevalence was computed as

$$\hat{\theta}_j = \sum_{k \in \{1,2,\ldots,K_j\}} \frac{N_k\, \hat{\theta}_k}{N_k},$$

where $\hat{\theta}_j$ is the estimated prevalence for aggregate group $j$, $k \in \{1,2,\ldots,K_j\}$ is the index for a given stratification group among the total number $K_j$ of stratification groups that make up the aggregate population $j$ of interest. The $N$'s refer to population sizes for each subgroup according to the ACS 2018-2022.

### Estimation of the care cascade

One complication in our analysis was estimation of the high BP care continuum, which organizes the high BP population into four decreasing levels:

1. Total hypertension (prevalence)
2. Aware
3. Treated
4. Controlled

Each level is nested within the previous level. In other words, we define individuals *aware* of their hypertension only among those who are in the *prevalent* population, whereas individuals who have *controlled* their high BP are defined only within the *treated* population. Conditional probabilities are a natural way to model the nested structure of the care cascade. The Pursuant data allows us to estimate a *partial* care cascade consisting of the following three levels:

1. Total hypertension (prevalence)
2. Aware
3. Controlled

Let $H$ denote high BP prevalence, $A$ denote awareness, and $C$ denote controlled. A key constraint required of any care cascade model is that the joint probabilities implied by the conditional probabilities cannot exceed any marginal probability. For example, an estimate for the proportion of controlled hypertension among the aware $P(C|A)$ should not imply a joint probability $P(C, A)$ that is higher than $P(A)$ or $P(C)$.

To enforce this constraint in estimation of the cascade, our approach was to fit each level of the cascade as a separate model, restricted to the subset of data corresponding to the denominator defined at that level. In other words, the model for hypertension control would only be fit on all subjects who are deemed "aware." The estimates from this model would then be conditional on hypertension awareness. Likewise, the model for hypertension awareness would be fit only on all subjects deemed "prevalent" and the model for hypertension prevalence would be fit on all subjects. Independent estimation of the conditional probabilities within each level of the care cascade would not directly violate any rules of probability.

While this approach works for generating estimates at the smallest subgroup level, i.e. county-state-age group-sex-race/ethnicity, difficulties arise when estimating at any higher aggregate level. Direct application of the poststratification weights using the ACS 2018-2022 would not apply to the estimates from the conditional awareness and controlled models because the poststratification weights reflect the national population, not the hypertension prevalent population or the hypertension aware population in which the conditional probabilities are defined over. Those weights only apply to marginal outcomes where the denominator is the national population as opposed to a subset like the high BP population or the aware population.

To solve this issue, for a given conditional probability to be estimated at an aggregate level with 95% CI, we made use of Bayes' Rule to reconcile our conditional model estimates with their poststratified estimates at any aggregate level. Suppose we are trying to estimate $P(A|H)$ for aggregate group $j$ which consists of mutually exclusive subgroups indexed by $k \in \{1,2, \ldots, n_j\}$, where $n_j$ is the total number of subgroups in $j$

and $N_k$ is the total population within subgroup $k$. Let $P_j^*(E)_i$ denote the poststratified probability estimate for event $E$ in aggregate group $j$ for draw $i$ as generated by the bootstrap samples in merMod. Let $P_k(E)_i$ denote the probability estimate for event $E$ in subgroup $k$ for draw $i$, directly output by the mixed effects model. Then the poststratified conditional estimate for $P(A|H)$ for the $i$th draw for group $j$ is

$$P_j^*(A|H)_i = \frac{P_j^*(A,H)_i}{P_j^*(H)_i},$$

where

$$P_j^*(H)_i = \sum_{k\in\{1,2,\ldots,n_j\}} \frac{N_k P_k(H)_i}{N_k},$$

are the high BP prevalence estimates and

$$P_j^*(A,H)_i = \sum_{k\in\{1,2,\ldots,n_j\}} \frac{N_k P_k(A,H)_i}{N_k} = \sum_{k\in\{1,2,\ldots,n_j\}} \frac{N_k P_k(A|H)_i P_k(H)_i}{N_k},$$

are the marginal high BP awareness estimates.

Poststratified estimates for controlled hypertension were computed with the same procedure (exchange $H$ for $A$ and $A$ for $C$). The mean and 95% CI's are calculated over 500 draws for each quantity. Future work will focus on how to incorporate correlations between geographic and demographic subunits in this procedure rather than assuming independence.
